# Disparities in adolescent mental health-related National Health Service presentations in England: secondary school cohort study

**DOI:** 10.1101/2025.11.03.25338372

**Authors:** Kate Lewis, Helen Dale, Dan Lewer, Johnny Downs, Ruth Blackburn

## Abstract

**Introduction:** Evidence on demographic and socioeconomic differences in adolescent mental health service use is vital for developing equitable health policy and service responses. We quantified inequalities in referrals to National Health Service (NHS)-funded mental health services and NHS-hospital use for mental health concerns by social strata for secondary school pupils in England.

**Methods:** We used linked records from state-funded schools and national health services (ECHILD) in England to create a secondary school cohort of pupils (age 11 years/Year 7 at entry) from 2012/13 to 2021/22. We measured rates of service use by gender, racial-ethnic group, region of residence, free school meal eligibility, and area-level deprivation defined by the index of multiple deprivation. Service contacts were captured by records in the Mental Health Services Data Set and Hospital Episode Statistics. Pupils were followed up until the first chronological event of: service contact, age 21 years, death or end of study (31st August 2022).

**Results:** Of 5,629,719 pupils, 980,648 (17.4%) were referred to NHS-funded mental health services; a rate of 45.2 per 1000 person-years (95% confidence interval 45.1-45.2). Within stratum, rates of referrals were higher in females (55.5, 55.4-55.7) than males (35.7, 35.6-35.8), highest in White Irish Traveller (70.8, 66.2-75.8) and lowest in Pakistani racial-ethnic groups (20.8, 20.5-21.1), highest in the North East (58.2, 57.7-58.7) and lowest in London (37.7, 37.5-37.9). Adolescents who were eligible for free school meals had particularly high referral rates (75.3, 75.0-75.6) and there was a gradient in rates by quintiles of area-level deprivation. There was consistency in the sociodemographic patterns across mental health-related referrals, emergency department attendances, and hospital admissions.

**Conclusions:** We find marked variation in mental health service use across social strata, but with consistency across different service types, indicating significant inequalities in the underlying prevalence of mental ill-health and access to services.

**Key Messages:** *What is already known on this topic:* summarise the state of scientific knowledge on this subject before you did your study and why this study needed to be done

- Improving access to mental health services for young people is a priority for the English National Health Service, but there is limited population-level information about rates of contacts across multiple health care services and with comprehensive coverage of England.

*What this study adds:* summarise what we now know as a result of this study that we did not know before

- Almost one fifth of pupils were referred to mental health services, with stark variation by gender, racial-ethnic group, free school meal eligibility, neighbourhood deprivation and region of residence
- Relative patterns of contacts by social strata were similar across emergency department attendances and hospital admissions

*How this study might affect research, practice or policy:* Summarise the implications of this study

- Clear differences in services contacts across social strata indicate significant inequalities, likely reflecting differential barriers to access, the acceptability of NHS services to specific population strata, as well as differences in the underlying prevalence of mental ill-health
- We find no evidence of disproportionate use of a single service type, suggesting that policies addressing inequalities should span across all services and must seek to address social determinants of health, including differential barriers to service use.

## Introduction

Adolescence is a key time for the emergence of mental health conditions. A meta-analysis in 2022 found that half of all mental disorders worldwide begin before age of 18 years.^1^ Whilst the benefits of interventions are thought to be greatest when implemented at the point of onset, considerable unmet need in young people is reported.^1,2^ In England, a wealthy country with a National Health Service (NHS) that is free for residents at point of use, it was estimated that around three-fifths of all under 18 year olds with a diagnosable mental health problem did not access mental health services in 2023/24.^3^ There are recognised inequities (*i.e.,* unjust and avoidable differences) in the prevalence of mental health disorders, driven by the unequal distribution of the social determinants of health and, subsequently, differential exposure to social, economic and environmental stressors.^4^ For example, in England in 2023, 55% of 11-16 year olds in families with self-reported financial worries had a probable mental disorder compared with 27% without these worries.^5^ Access to formal mental health services is also likely to be inequitable.^6^ Qualitative research has highlighted barriers in both the accessibility of services (including information provision, service capacity and culture/gender representation amongst professionals) and in the ability of populations to access services (including health beliefs, stigma or shame and practical difficulties, such as transport).^7–11^

In recognition of disparities in appropriate care, NHS England has set access for mental health services for 0-17 year olds for “certain ethnic groups, age, gender and deprivation” groups as one of five clinical areas requiring accelerated improvement for children and young people.^12^ The NHS 10 years plan emphasises early intervention and local approaches to narrowing mental health inequalities.^13^ Despite these policy priorities, detailed population-based information on mental health-related contacts across multiple healthcare settings and social strata, including region if residence, is lacking. Given the rising demand for services, indicated by surveys of mental health prevalence and rates of admissions to acute paediatric wards for mental health concerns,^5,14^ establishing national patterns of service contacts is a critical step to ensuring equitable service provision. Estimating disparities in contacts across services may highlight where inequities exist and for whom. For example, adolescents who face barriers to accessing out-patient mental health support, may disproportionately end up in emergency care. Using national linked education and health records in England, this study describes rates of mental health-related NHS presentations among adolescents, by key social strata (region, gender, racial-ethnic group, socioeconomic deprivation) and other high-risk populations (care experienced, chronic conditions, special educational needs and disability (SEND) provision, persistently absent from education). We examined rates of NHS mental health service referrals, emergency department (ED) attendances and emergency hospital admissions.

## Methods

### Public and patient involvement and engagement (PPIE)

We consulted with a young people’s and parent’s PPIE group during the planning and results stages of this study. These groups helped us to shape the research, interpret the results and identify limitations of this study. For example, sessions with parents led to the inclusion of the “refused” racial-ethnic category in our analysis (as opposed to searching for non-refused or missing racial-ethnic group across longitudinal records) and helped to contextualise our findings on non-linkage, particularly around opt outs among some minoritised ethnic groups. In the young people’s sessions, we gained insight into the pathways between social strata and referrals to mental health services. Critiques of the project centred around what was missing from our planned analyses, namely early life risk factors, some marginalised groups of young people, types of mental health service provision and characteristics of service providers. This led to additional groups being included in our analysis (including persistently absent pupils) and, where this was not possible, has provided direction for further research.

### Datasets and linkage

This study used the ‘Education and Child Health Insights from Linked Data’ (ECHILD) database, which contains individually linked and de-identified education, social care and health records for pupils in England.^15,16^ The data sources used in this study were the National Pupil Database, Hospital Episode Statistics, the Mental Health Services Data Set and Office for National Statistics (ONS) mortality records. The National Pupil Database contains information about state-funded schools in England, including school enrolments, pupil characteristics, recorded SEND provision, attainment, absences, and exclusions. It also contains two children’s social care modules; the “Children Looked After” return, relating to children in care, and the “Children In Need” census, relating to children referred to or needing additional support from social care services. Hospital Episode Statistics contains administrative, demographic and clinical information on inpatient admissions (admitted patient care records), outpatient appointments and ED attendance activity (the Emergency Care Data Set, ECDS). The Mental Health Services Data Set contains referrals to and contacts with wholly or partially NHS-funded mental health services. ONS mortality records provide information on the cause and date of deaths registered in England and Wales, including those that occur outside of hospital.^18^

These datasets were linked by NHS England using a two-stage deterministic linkage algorithm using natural identifiers (forename, surname, sex, date of birth and postcode), with reported linkage rates of 98% between the National Pupil Database and Hospital Episode Statistics for children born in 2004/05.^15^ To be eligible to be linked to health datasets, pupils were required to have had any NHS contact (*e.g.,* registration with a general practice) and to not have opted out of sharing their data for research through the national data opt-out programme.^19^ Data in the ECHILD database has been de-identified, meaning that direct identifiers such as names, addresses, postcodes or dates of birth have been removed. Table S1 gives further details of the ECHILD datasets used in this study.

### Study Design

Secondary school cohort study (see Figure S1).

### Population

Our population was defined as adolescents enrolled in Year 7 of state-funded secondary school in England between 2012/13 and 2021/22, corresponding to age 11 years at entry (Figure S1). Enrolment was defined as a record in the January School Census, alternative provision or pupil referral unit census. For pupils not following the national curriculum (such as some pupils receiving SEND provision), we assigned a school Year assigned to them based on their age at entry (i.e., age 11 years for Year 7). We excluded pupils; i) who did not link to the health datasets, including those with national data opt outs applied to their health records,^20^ ii) aged <10 or >12 years old at entry into Year 7, iii) who had missing data on residential address or had a Middle Layer Super Output Area indicating Welsh/Scottish residence.

### Follow-up

Start of follow-up differs by outcome (see Figure S1). For hospital admissions, follow-up begins from entry into Year 7. For mental health services, follow-up begins on 1 September 2016 or entry into Year 7 thereafter, to coincide with the availability of Mental Health Services Data Set. For ED attendances, follow-up begins on 1 September 2020 (or entry into Year 7 thereafter) to coincide with the availability of the ECDS. For all outcomes, follow-up ends at the first chronological event of: outcome event, death, age 21 years or end of study (31^st^ August 2022). We capture only the first of each event during secondary school, rather than all events, to prevent rates from being skewed by repeated contacts from the same individuals.

### Social strata

Social strata were selected to align (where possible) with NHS England’s target populations for reducing health inequalities (the Core20PLUS5).^12^ We use the term social strata to mean the different positions occupied by individuals in society based on their socially constructed identities and socioeconomic circumstances.^21^ The theoretical framework guiding the proposed pathways between social strata and healthcare contacts is shown in **Figure S2.** We included: parent-reported gender (female or male, only two options available); region of residence (nine groups mapped to middle layer super output area of the child’s residential address); index of multiple deprivation (IMD) quintiles based on residential address; free school meal (FSM) eligibility (yes or no); and racial-ethnic group (derived from the parent-reported 20-category minor variable). We purposefully use the term “racial-ethnic” to signify that this variable contains both race-based (e.g. White, Black) and ethnicity-based (e.g. British, Caribbean) identifiers. Social strata were defined at January Census of Year 7 (or nearest prior non-missing Spring Census).

### High-risk populations

We included a secondary group of circumstances that are associated with, or indicative of, poorer mental health in adolescence to provide currently lacking information on groups of adolescents at high needs of mental health support: care experienced (yes or no), defined as a child who has been in the care of their local authority for more than 24 hours in the 5 years prior to study entry (between school Years 1-6, ages 5/6-10/11 years); chronic conditions, defined by the presence of diagnosis codes from the Hardelid code list in any hospital admission in the 5 years prior to study entry (between school Years 1-6, ages 5/6-10/11 years; see Supplementary Appendix 1);^22^ recorded SEND provision (including SEND support and education health and care plans) in National Pupil Database at entry into the cohort; and persistent absence, defined as 10% or more missed sessions in Year 6 (ages 10/11 years old) or Year 4 and Year 5 for the 2021/22 and 2020/21 cohorts, respectively (as absence data is not available for 2019/20-2020/21 due to the COVID-19 pandemic).

### Covariates

We stratified results by age at referral (11-15 years, 16-17 years, 18-20 years) due to differences in mental health presentations, referral pathways and thresholds for hospital admissions by age.^23,24^ Age at referral was calculated as date of the outcome event minus date of birth. Date of birth was derived from month and year of birth, setting the day to the 15^th^ for all children as full date of birth is not available in ECHILD.

### Outcomes

We used three sources of mental health-related outcomes in this study: referrals to mental health services; emergency hospital admissions; and ED attendances.

#### Referrals to NHS-funded mental health services

We captured rates of referrals to NHS-funded mental health services, including children and adolescent mental health services (CAMHS, also known as CYPMHS) per 1000 person-years using the Mental Health Services Data Set. We excluded referrals that were likely to be only related to neurodevelopmental concerns/neurodiversity-related difficulties using reason for referral and service type codes (Table S2). We described the proportion of all referrals and, separately, rates (per 1000 person-years) of records that indicated a “crisis” referral, by social strata and high-risk groups (Table S2).

#### Mental health-related ED attendances

We defined mental health-related ED attendances as any ECDS record during the follow-up period with a primary diagnosis/chief complaint code appearing in either of two NHS England Systematized Nomenclature of Medicine Clinical Terms (SNOMED CT) code lists (Table S3). The first list is the ECDS psychology/toxicology/drug and alcohol group, including common mental health disorders, harmful effects from drugs, medicaments and other substances.^25^ Attendances were included in this definition if the code appeared in the primary diagnosis position, identified as the lowest sequence diagnosis number. The second list is the psychosocial/behaviour change reference set, which encompasses attendances with a chief complaint of self-injurious behaviour, depressed mood, suicidal thoughts, anxiety, bizarre behaviour, physical aggression, feeling agitated, hallucinations or delusions.^25^

#### Mental health-related emergency hospital admissions

We defined mental health-related emergency hospital admissions as a hospital admission during the follow-up period with: an emergency admission method in the first episode of care; and International Classification of Diseases 10th Revision (ICD-10) code(s) recorded in any diagnosis field indicating mental disorder or adversity-related injury (Table S4).^26^ Mental disorders include mental health diagnoses as indicated by an F chapter (mental, behavioural, and neurodevelopmental disorders) ICD-10 code in any diagnostic position. Adversity-related injuries were indicated by any S or T (injuries, poisoning, and other consequences of external causes) chapter ICD-10 code alongside an ICD-10 code indicating that the harm was either self-inflicted or inflicted by another person (as opposed to accidental) in any diagnostic position.^26^ In sensitivity analyses, we also included potentially psychosomatic symptoms in our definition of emergency hospital admissions, as indicated by ICD-10 codes related to pain, cardiovascular/respiratory, digestive, skin, sleep and fatigue.^24^ These are used to indicate potential manifestations of stress as the main reason for admission (when not accompanied by a medical or surgical operation/diagnostic code in the same admission; Table S5).^24^

### Statistical analyses

First, we described the characteristics of our study population, including missing data, in numbers and percentages. Observed incidence rates of referrals to mental health services, hospital admissions and ED attendances for each social strata and high-risk population group were calculated by dividing the number of events by person-time at risk per 1000 person-years. We then fit separate Poisson regression models for each outcome, stratified by social strata and age band, including the logarithm of person-time as the offset. Stratification by age band was necessary to account for known differences in mental health needs and presentations by age.^14,24,27^ We adjusted for follow-up year to account for likely period effects in the data, particularly during the COVID-19 pandemic.^14,28^ We estimated year-adjusted incidence rates (per 1000 person-years) for each social strata and age group. We further estimated incidence rate ratios (IRRs) comparing outcomes within social strata and age group, using the observation weighted grand mean predicted rate as the reference group for ratios to avoid delineating non-reference groups as “other”.^29^ In additional analyses, we further stratified models by gender to explore intersectional experiences of social identities^14^. Statistical analyses were conducted using Stata v18.0 in the ONS Secure Research Service. The code and algorithms used in this study are available from: https://github.com/UCL-CHIG/Mental_health_contacts. We made some minor deviations from our study protocol based on data availability at the time of analysis, including reducing the study end date by a year and not reporting mental health service waiting times.^30^

## Results

### Cohort derivation and characteristics

There were 5,920,062 pupils enrolled in Year 7 of state-funded secondary school in England between 2012/13 and 2021/22 (Figure S3). We excluded 28,599 (0.5%) pupils with a missing or non-English residential address or who were aged <10 or >12 years at entry into Year 7. We further excluded 266,716 (4.5%) pupils who did not have a linked health record. Non-linkage rates were particularly high amongst pupils from Black Caribbean (11,451/72,558; 15.8%), Black (other) (4,136/42,063; 9.8%) and “refused” (3,867/43,337; 8.9%) racial-ethnic groups, as well as those with a residential address in London (61,990/879,271; 7.1%), Table S6. Finally, we excluded 13 pupils who had a death recorded before estimated entry into Year 7. The final cohort included 5,629,719 pupils, of whom 51.3% (2,886,323) were male, 69.6% (3,917,140) were White British and 30.4% (1,712,435) had a residential address in London or the South East (Table 1).

**Table 1.**
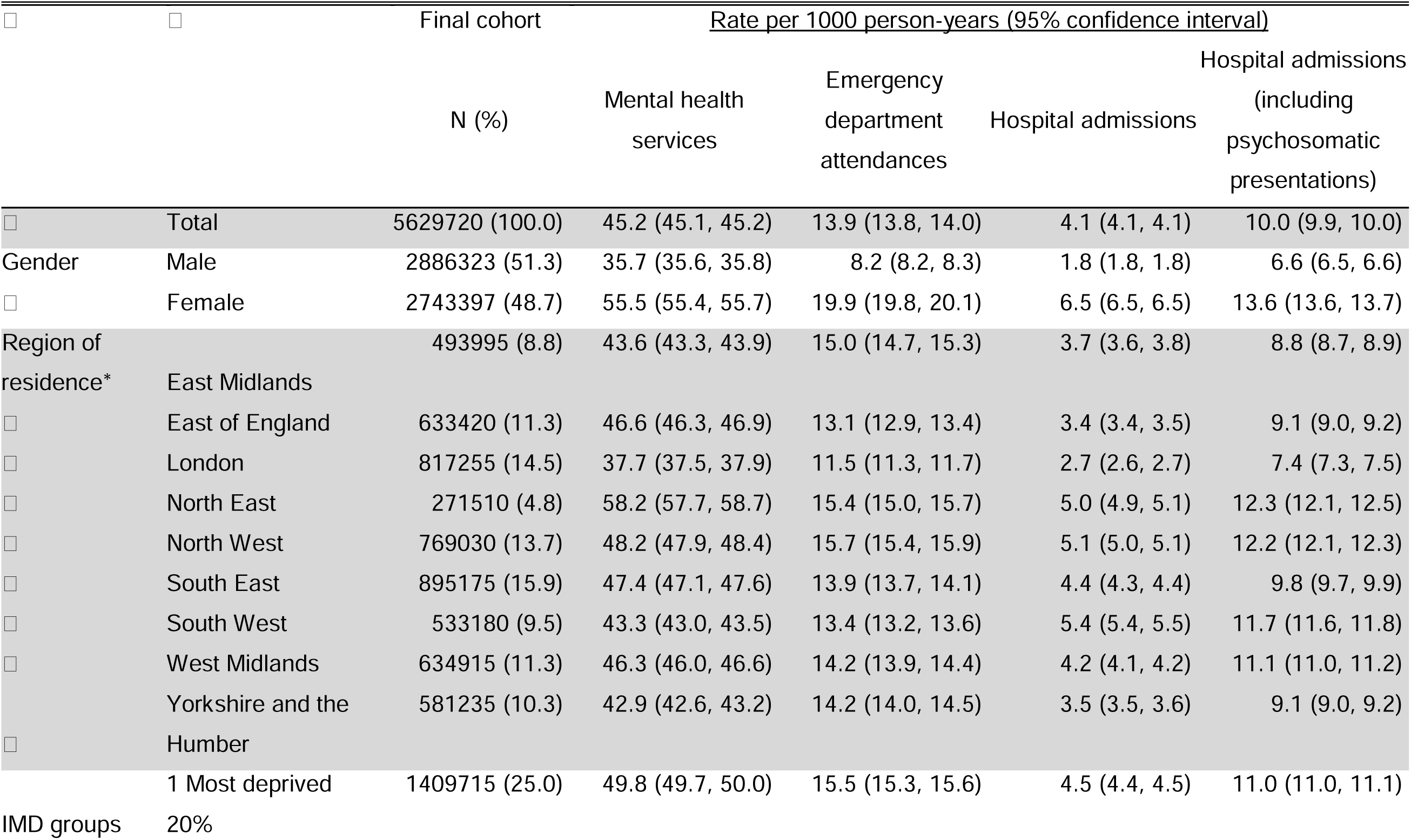
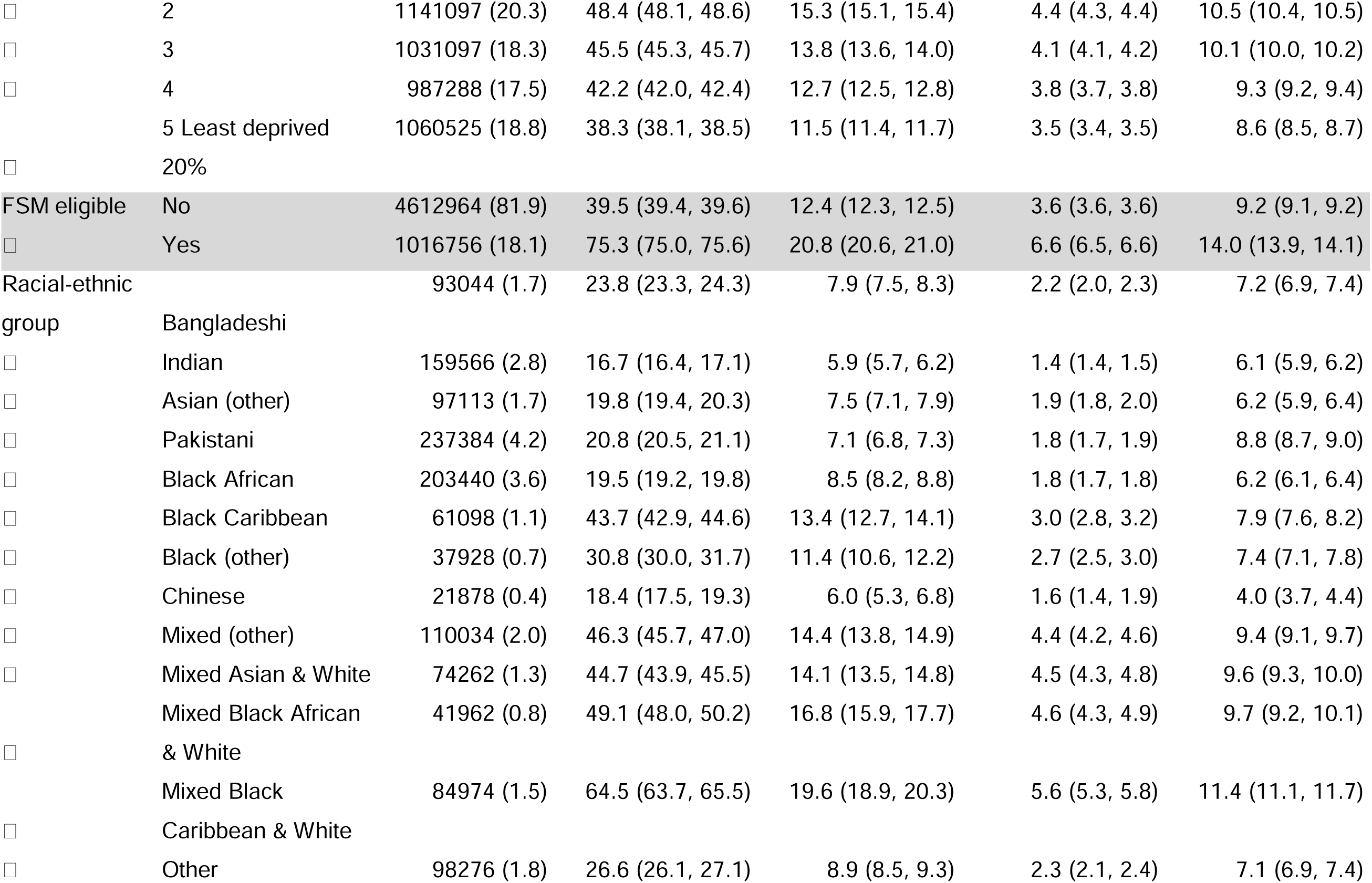
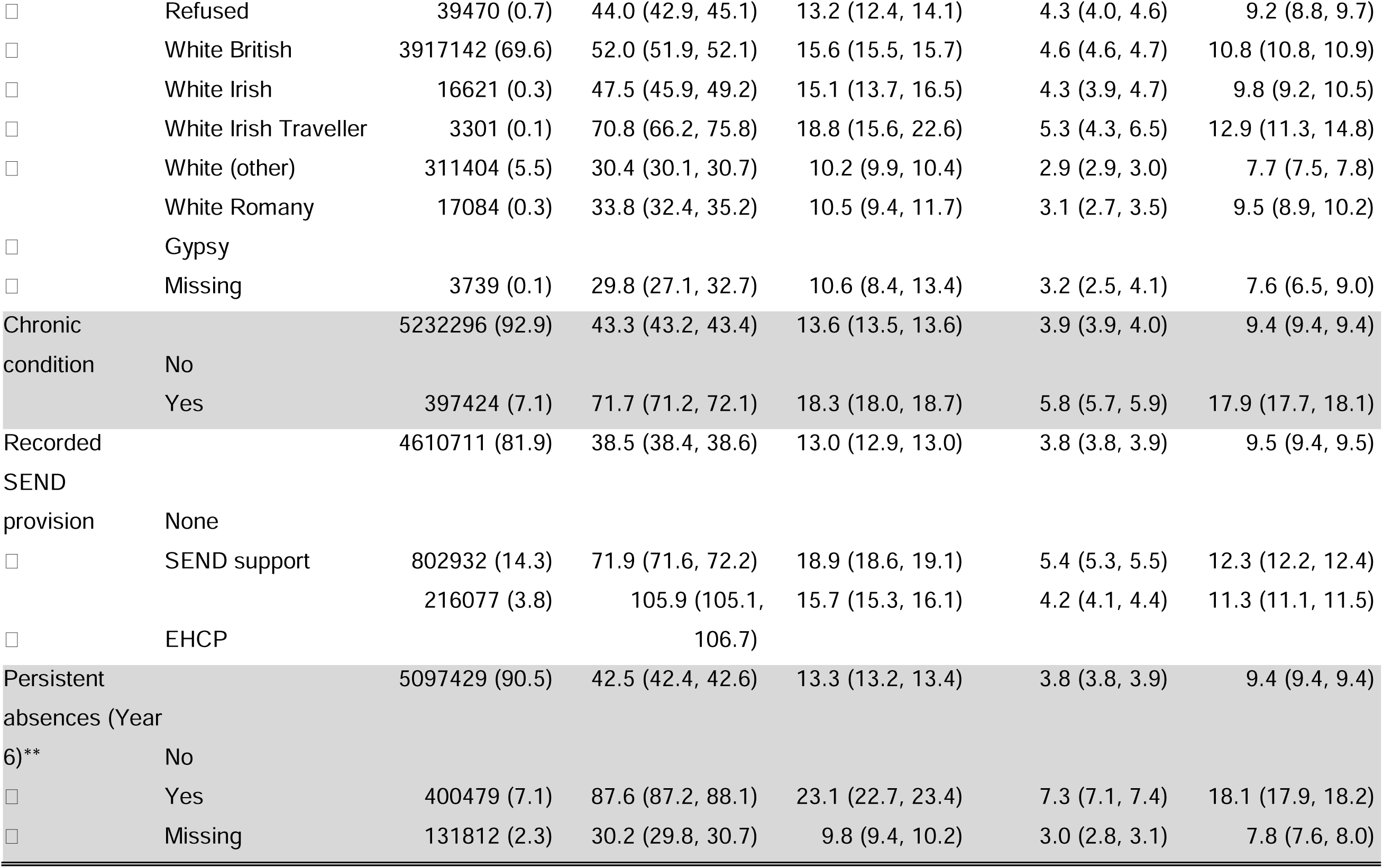
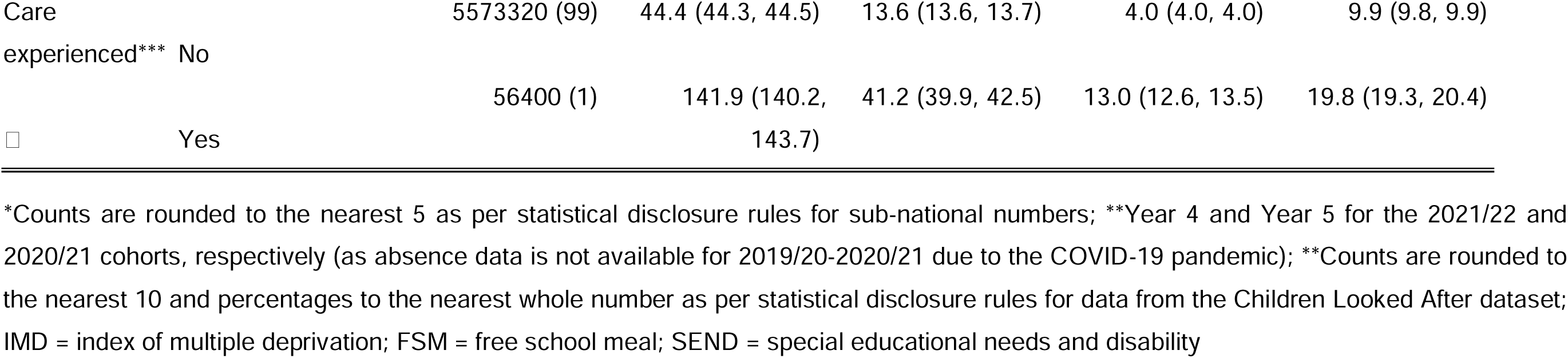
Final cohort characteristics and unadjusted rates of mental health-related contacts across service types.

### Referrals to mental health services

Across all ages during follow-up (mean = 3.86 years, standard deviation = 1.95), 17.4% (980,648) of pupils had a recorded referral to mental health services, a rate of 45.2 per 1000 person-years (95% CI 45.1, 45.2; Table 1). Overall, 1.8% (17,486) of these referrals met our definition for crisis referral, Table S7. There was marked variation in unadjusted rates of referrals by social strata (Table 1). Females (55.5, 95% CI 55.4, 55.7) had higher observed rates of referrals than males (35.7, 95% CI 35.6, 35.8). Within region, observed rates were highest in the North East (58.2, 95% CI 57.7, 58.7) and lowest in London (37.7, 95% CI 37.5, 37.9). There was a gradient in referral rates by IMD quintiles, from 38.3 (95% CI 38.1, 38.5) among pupils living in the least deprived to 49.8 (95% CI 49.7, 50.0) in the most deprived areas. Pupils who were FSM eligible had the highest observed rates of referrals of any social strata studied (75.3, 95% CI 75.0, 75.6). Of the racial-ethnic groups, the lowest rates of referrals were observed among Indian and Black African groups and highest among the mixed Black Caribbean-White and White Irish Traveller groups. All high-risk populations had higher observed rates of referrals than average, including adolescents with care experience (141.9, 95% CI 140.2, 143.7), those with a recorded EHCP (105.9, 95% CI 105.1, 106.7), with persistent absence in Year 6 (87.6, 95% CI 87.2, 88.1) and hospital record defined chronic conditions (71.7, 95% CI 71.2, 72.1). Unadjusted rates of first crisis referral followed similar stratum-specific patterns to all referrals, asides from region, where the South West, West Midlands and South East had lower rates than London (Table S7).

When grouped by age, stratum-specific rates by gender, region, IMD and FSM eligibility showed consistent patterning, with higher rates among the youngest age groups (11-<16 years; Figure 1; Table S8). In contrast, several racial-ethnic groups had similar rates among 11-<16 and 16-<18-year-olds; Bangladeshi, Indian, Asian (other), Pakistani, Black African, Black (other), Chinese, Other, “refused”, White Irish Traveller, White Romany Gypsy and those with missing data on racial-ethnic group. Relative differences (IRRs) in rates, shown in Figure S4 and Table S9, highlights that relative differences between racial-ethnic groups tended to reduce as age group increased. Further stratification by gender for the 11-<16-year-old group, indicated in Figures S5 (rates) and S6 (IRRs), highlights the overlap in rates for females and males from White Irish Traveller and White Romany Gypsy racial-ethnic groups.

**Figure 1.**
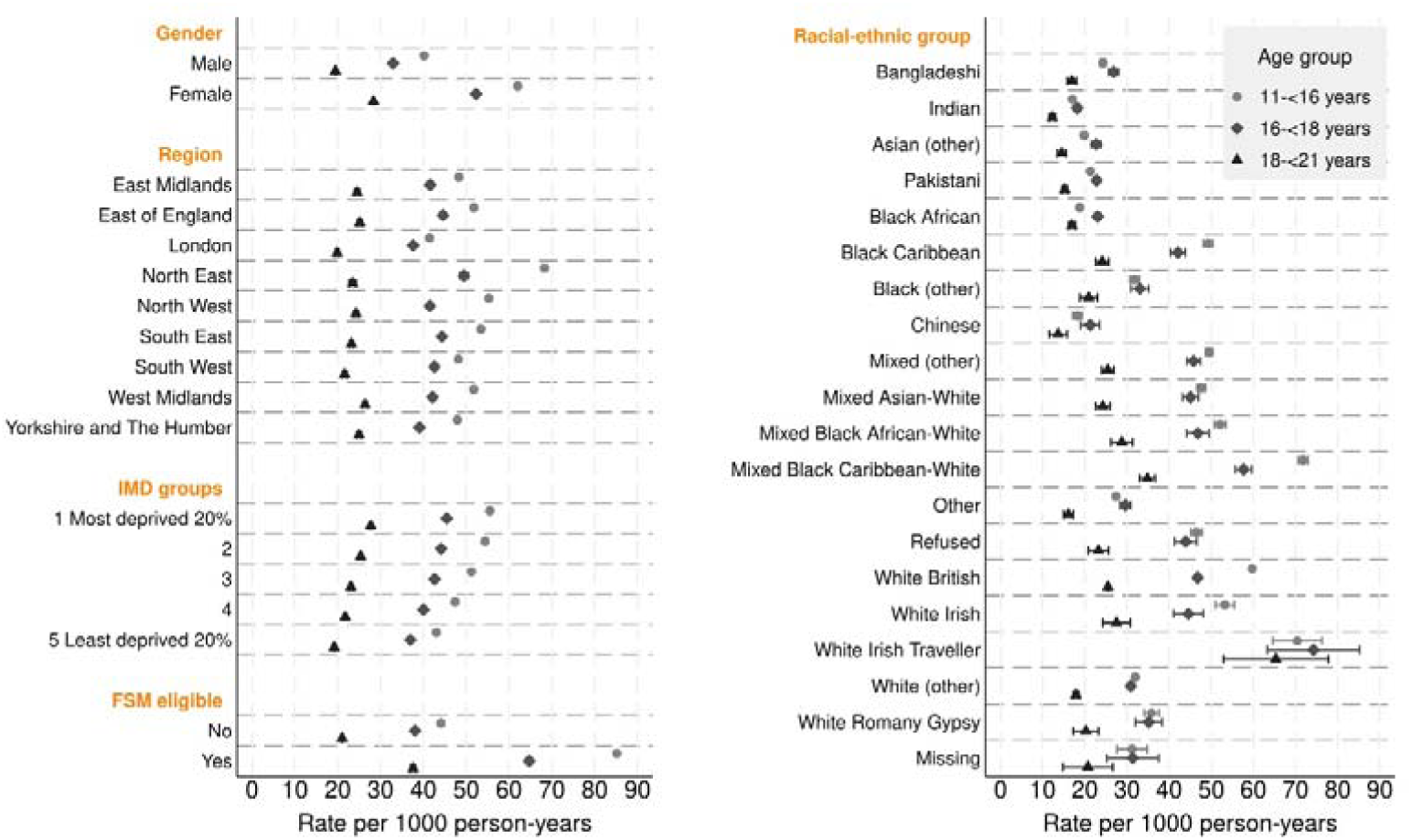
Year-adjusted rates of first referral to NHS-funded mental health services (per 1000 person-years) with 95% confidence intervals: derived from age group-specific Poisson regression models for each group of social strata. FSM = free school meals; IMD = index of multiple deprivation.

### Mental health-related ED attendances

Across all ages during follow-up (mean = 1.82 years, standard deviation = 0.39), 2.5% (142,118) of pupils had a recorded mental health-related ED attendance, a rate of 13.9 per 1000 person-years (95% CI 13.8, 14.0; Table 1). Variation in in unadjusted rates of referrals by gender was marked (Table 1), with females (19.9, 95% CI 19.8, 20.1) having more than double the observed rates of males (8.2, 95% CI 8.2, 8.3). Within region of residence, rates were highest in the North West (15.7, 95% CI 15.4, 15.9) and lowest in London (11.5, 95% CI 11.3, 11.7). A gradient in rates was present across IMD groups. Pupils who were FSM eligible had the highest observed rates of referrals of any social strata studied (20.8, 95% CI 20.6, 21.0). Of the included racial-ethnic groups, the lowest rates of referrals were observed among Indian and Chinese groups and highest among the mixed White-Black Caribbean and White Irish Traveller groups. All high-risk populations had higher observed rates of referrals than average, particularly adolescents with care experience (41.2, 95% CI 39.9, 42.5), with persistent absence in Year 6 (23.1, 95% CI 22.7, 23.4).

When grouped by age, stratum-specific rates broadly increased with older age, with some 95% CI overlaps for the two older age groups (Figure 2; Table S10). Relative differences (IRRs) in rates were particularly high among females age 11-<16 years compared to the other age groups (Figure S7; Table S11). Further stratification by gender for the 11-<16 year old group in Figures S8 (rates) and Figure S9 (IRRs) indicated particularly low rates for females from the Chinese racial-ethnic group.

**Figure 2.**
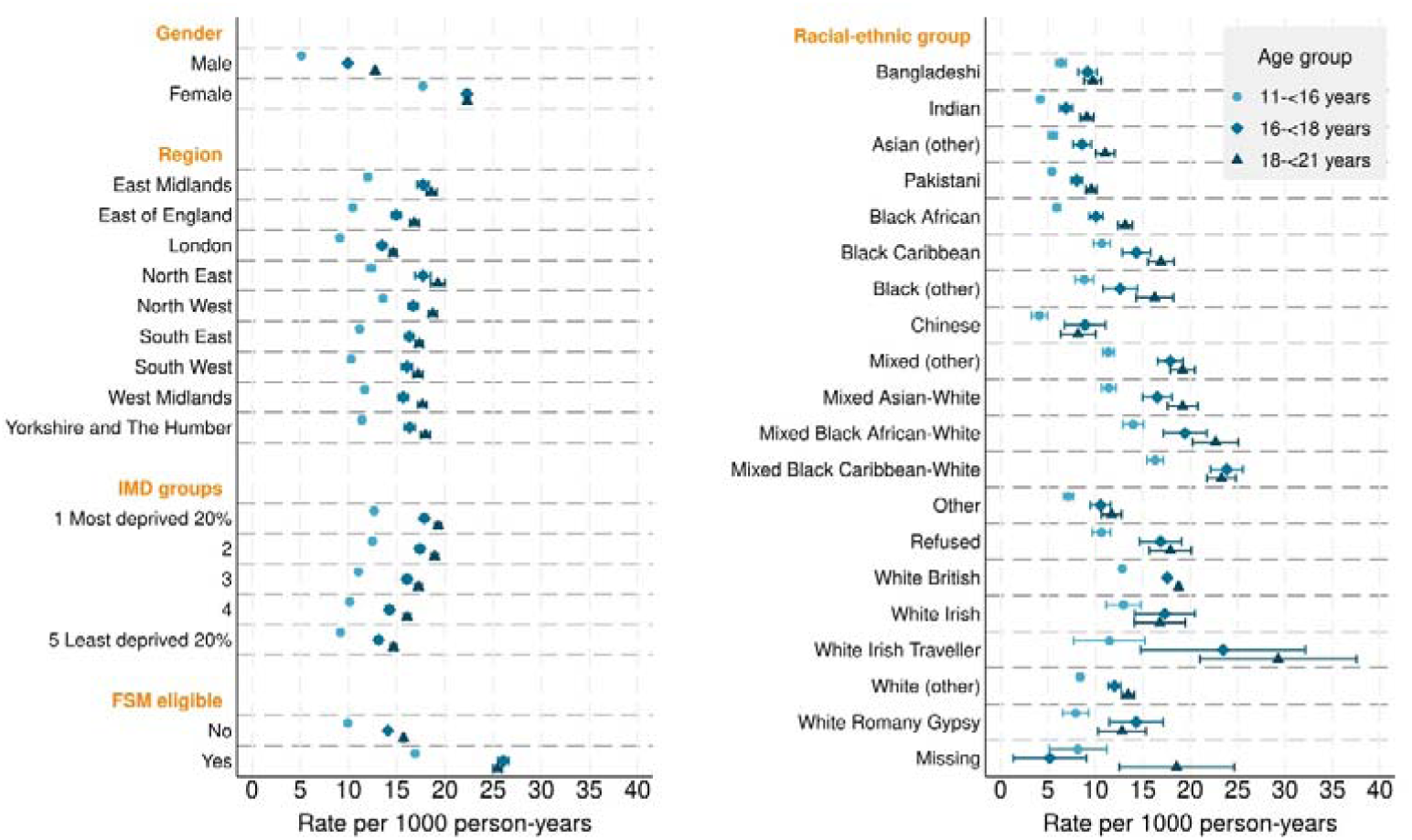
Year-adjusted rates of first mental health-related emergency department attendance (per 1000 person-years) with 95% confidence intervals: derived from age group-specific Poisson regression models for each group of social strata. FSM = free school meals; IMD = index of multiple deprivation.

### Mental health-related emergency hospital admissions

Across all ages during follow-up (mean = 5.18 years, standard deviation = 2.78), 2.1% (118,680) of pupils had a recorded mental health-related emergency hospital admission, a rate of 4.1 per 1000 person-years (95% CI 4.1, 4.1; Table 1). Females (6.5, 95% CI 6.5, 6.5) had more than three times the observed rates of males (1.8, 95% CI 1.8, 1.8). Within region, rates were highest in the South West (5.4, 95% CI 5.4, 5.5) and lowest in London (2.7, 95% CI 2.6, 2.7). A gradual social gradient was estimated across IMD groups. Pupils who were FSM eligible had the highest observed rates of referrals of any social strata studied (6.6, 95% CI 6.5, 6.6). Of the included racial-ethnic groups, the lowest rates of referrals were observed among Indian and Chinese adolescents and highest among mixed White-Black Caribbean and White Irish Traveller groups. All high-risk populations had higher observed rates of referrals than average, including adolescents with care experience (13.0, 95% CI 12.6, 13.5) and persistent absence in Year 6 (7.3, 95% CI 7.1, 7.4).

When grouped by age, stratum-specific admission rates were not consistently patterned (Figure 3; Table S12): rates increased with age for males but were lowest in the 18-<21-year-old group for females. Regional rates were similar across age-groups aside from the South East and South West, where the 16-<18-year-old group had the highest rates. Rates were similar across age groups for racial-ethnic groups, apart from the Asian (other), Pakistani and Black African groups, whom had lower rates in the youngest age group. Relative differences (IRRs) in rates are indicated in Figure S10 and Table S13, with further stratification by gender for the 11-<16 year old group in Figures S11 (rates) and S12 (IRRs). These indicate that relative rates within-gender were similar across all strata asides from Bangladeshi (higher in males) and White (other) (higher in females) adolescents.

**Figure 3.**
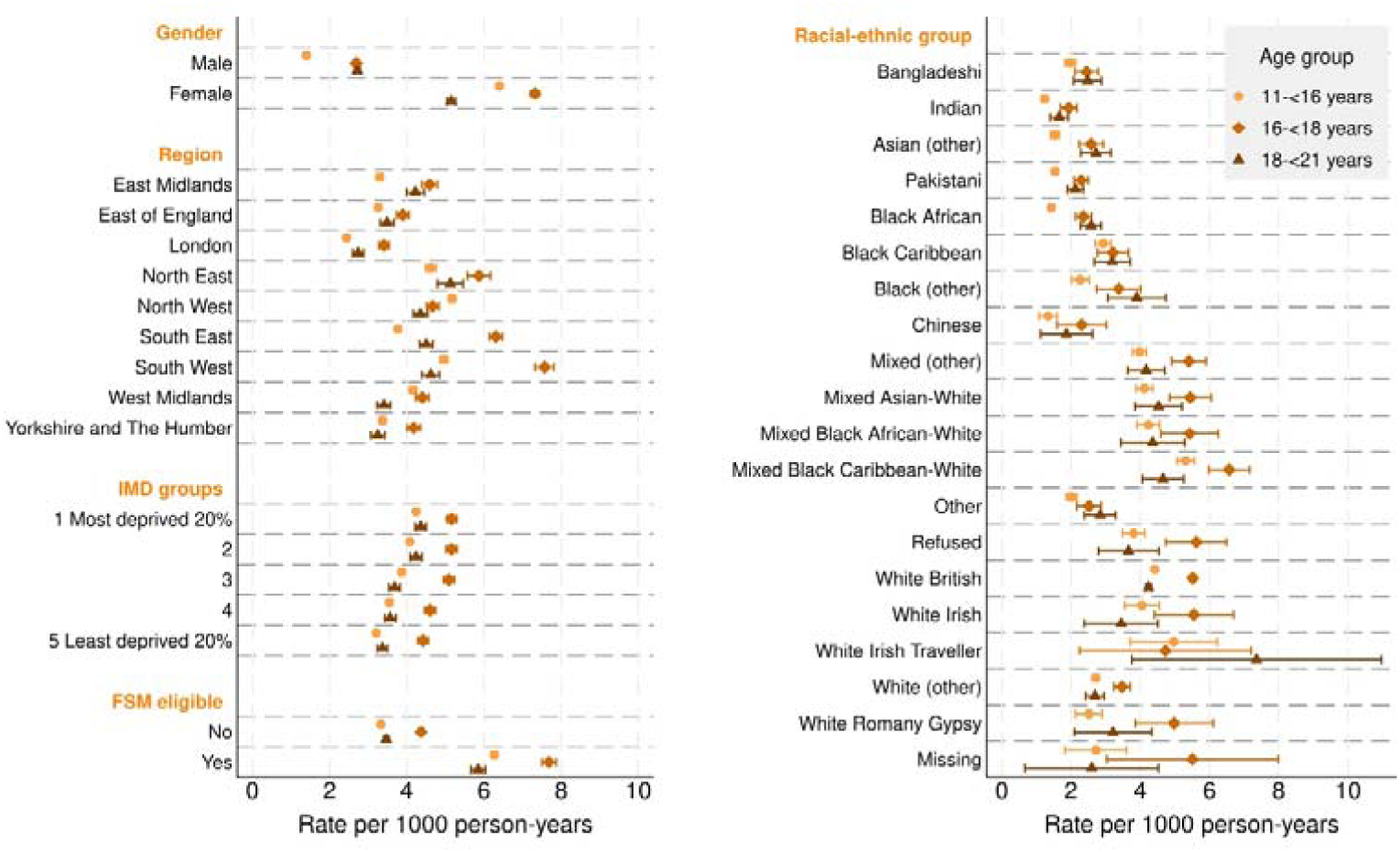
Year-adjusted rates of first mental health-related emergency hospital admissions (per 1000 person-years) with 95% confidence intervals: derived from Poisson regression models with separate models for each social strata and age group. FSM = free school meals; IMD = index of multiple deprivation.

Adding potentially psychosomatic symptoms into the definition of mental health-related emergency hospital admissions increased the overall rate of admissions to 10.0 per 1000 person-years (95% CI 9.9, 10.0), Table 1, but a generally similar patterning by strata remained (Figure S13; Table S14). Relative measures (by age group; Figure S14; Table S15) indicated that the broader case definition for admissions typically resulted in shift in stratum specific IRRs that were closer to the mean. This shift towards the mean was particularly pronounced for Pakistani adolescents aged 11-<16 years (from an IRR of 0.43 (95% 0.41, 0.45) to 0.92 (95% CI 0.90, 0.94).

### Combined relative results (11-<16 year old age group only)

Figure 4 displays the relative results across all three service contact points for the youngest age group. This highlights qualitatively similar relative differences within social strata. For example, IRRs of less than one for most Asian groups (Bangladeshi, Indian, Asian (other), Pakistani, Chinese), and IRRs at or above one for Mixed groups (Mixed (other), Mixed Asian-White, Mixed Black African, Mixed Black Caribbean-White), White British, White Irish and White Irish Traveller groups. The one exception is region; adolescents living in the East Midlands had higher relative rates of ED attendances than average and those living in the South West had higher relative rates of hospital admissions (but lower relative rates for other services).

**Figure 4.**
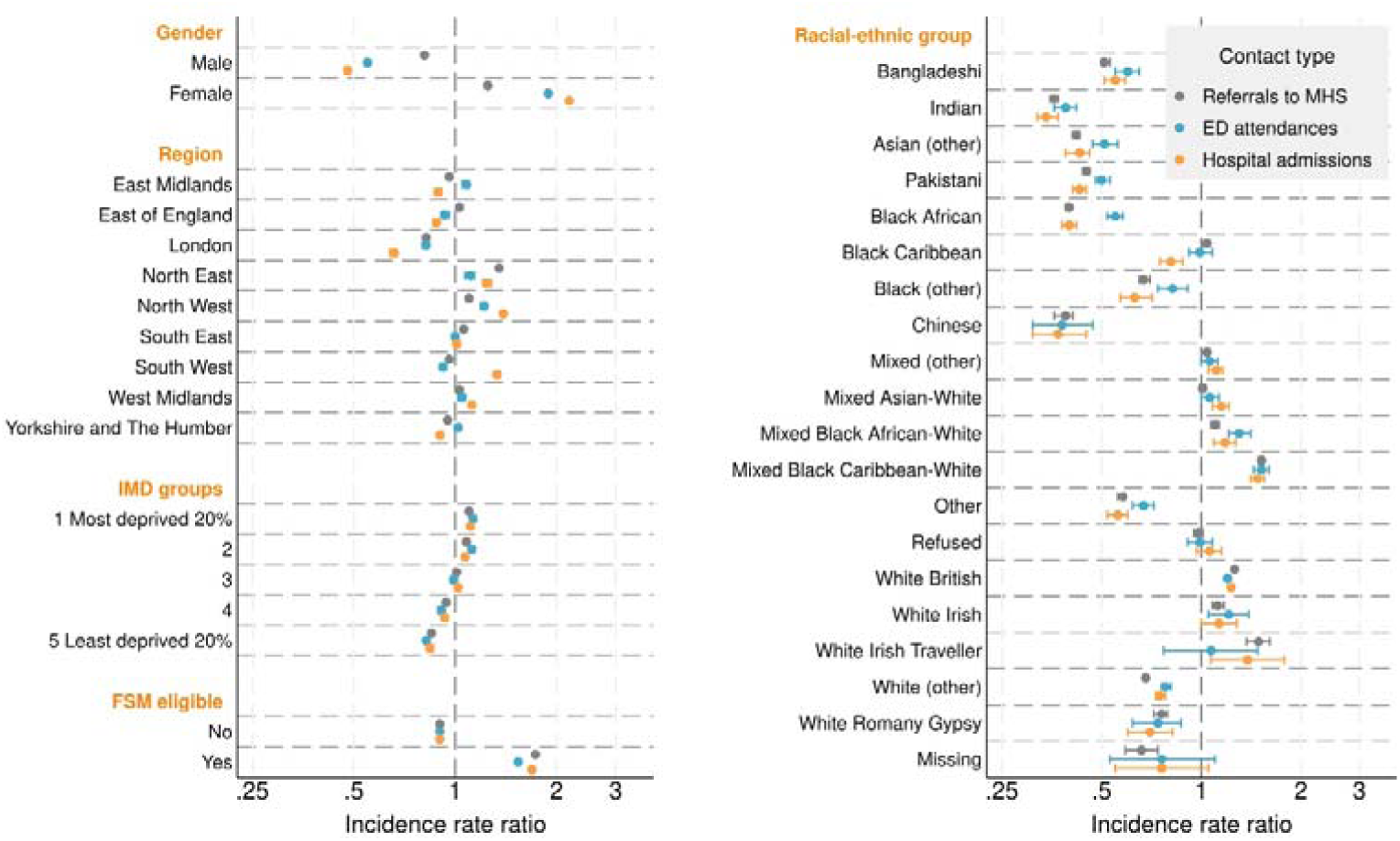
Year-adjusted incidence rate ratios (compared to the social strata- and service-specific mean rate) of first referrals to mental health services, mental health-related emergency department attendances and emergency hospital admissions: derived from service-specific Poisson regression models with separate models for each social strata (11-<16 years old group only). ED = emergency department; FSM = free school meals; IMD = index of multiple deprivation; MHS = mental health services.

## Discussion

In this national cohort study of secondary school pupils in England, we find clear inequalities in rates of mental health-related service contacts by social strata. Notably higher rates of referrals to mental health services, ED attendances and hospital admissions were shown for females, adolescents who were FSM eligible, in the most deprived IMD groups, living in the North, White Irish Travellers and Mixed Black Caribbean-White. Consistently lower rates were estimated for males, adolescents living in London and several racial-ethnic groups, including Bangladeshi, Indian, Pakistani and Black African. All high-risk groups had higher than average rates, particularly care experienced adolescents who had three times the average rate for referrals to mental health services and ED attendances. There was consistency in relative rates across services, particularly for socioeconomic circumstances (IMD and FSM eligibility) and racial-ethnic groups, indicative of systematic inequalities underpinned by social determinants.

### Strengths and limitations

To our knowledge, this the first national study using a secondary school cohort to look at patterns of mental health contacts across multiple NHS services, which is a critical gap given evidence of rising rates of urgent and emergency healthcare for adolescent mental health.^14^ The large population allowed for granular analysis of social strata and other high-risk groups. Assessment of contacts across three health contact points provides us with a richer understanding of where mental health support is being sought and whether inequalities are most evident at the level of the social strata and/or by service. However, the extent to which unequal distribution of service contacts exemplifies inequities at the point of trying to reach services cannot be quantified using service contacts alone. In follow-up work, findings from this study will be triangulated with evidence on young people’s mental health service use in Bradford, which includes indicators of underlying mental health needs;^30^ a key facet of mental health access that is not captured in our administrative datasets. Due to data quality concerns, we focussed on referrals to mental health services (a mandatory field in the Mental Health Services Data Set), which gives a joint indication of perceived mental health needs paired with healthcare seeking and reaching, but does not indicate whether treatment was received. The ECDS was only available for two years of follow-up in our study (2020/21 and 2021/22), which overlaps with some of the COVID-19 lockdown period,^31^ when ED attendance rates were lower. During this time there was a limited list of SNOMED codes available to report clinical information in EDs, which may have led to underreporting of mental health-related events. However, a strength of our study is that we applied standardised code lists that have been created to improve accuracy and standardisation in clinical recording of ECDS submissions.^32^

Only pupils who enrolled in Year 7 of a state-funded school during the study period are included in our cohort. Alongside home schooling and exclusion, the most common reason for not attending a state-funded school is attendance at a fee-paying school (6-7% of the population),^33^ meaning our results likely underrepresent pupils from advantaged backgrounds. In total, 4.5% of pupils were not linked to the health datasets and were also excluded from this study. Linkage was non-random and varied substantially by racial-ethnic group (higher in all minoritised ethnicities, particularly among Black Caribbean pupils). Non-links can occur for several reasons, including no contact with the NHS, missed links due to inconsistently recorded personal information, and data opt outs (accounting for 87% of non-links across ECHILD).^34^ We also excluded a smaller number of adolescents with missing residential information, which may include populations who are at greater risk of mental ill-health, such as those experiencing housing instability.^35^ Other relevant social strata were unmeasured in our study, such as sexual and gender minorities, whom have particularly elevated rates of mental distress and face additional barriers to care compared to peers.^8^ Only two genders, based on reported gender (replaced with legal sex in 2023/24), were available for analysis. Whilst self-declared gender identity can also be recorded by educational establishments since 2023/24, this information is not available for analysis in the National Pupil Database.^36^

### Interpretation and implications

Our main finding is that there are substantial disparities in rates of health service contacts related to mental health among adolescents in England. Similar associations have been observed elsewhere, including: higher rates of primary or secondary healthcare service use for emotional problems in female compared to male secondary school pupils in the Midlands and South West England;^37^ higher reported contact with any professional services among White young people with a diagnosed mental health condition compared with young people from other racial groups in a UK-based survey;^23^ and a social gradient between area-level deprivation and referrals to specialist mental health services in Scotland.^38^ We find that rates of contacts for pupils that are FSM eligible that area up to twice that of pupils living in most deprived area, indicating the importance of individual-level measures for capturing the structural conditions that lead to higher levels of mental health needs.

We present rates for more granular racial-ethnic groups than typically reported, highlighting the unique experiences of people from culturally diverse ethnic groups. This is particularly stark within marginalised populations usually grouped under racialised categories “White” (*e.g.,* White Irish Travellers with higher and White Romany Gypsy with lower than average rates across all service types) and “Black” (*e.g.,* Black African with near average and Black Caribbean with lower than average rates across all service types). Further, by presenting results also stratified by gender we show that the gendered social risk factors are experienced differently across racial-ethnic groups. We also show that region of residence, which is unexplored in similar studies, is an important source of variation in mental health-related contacts. Differences in models of service delivery across England, as well as density of services and populations, may place some populations at further risk of inequalities in provision. Future research could use methods grounded in intersectional theory, such as the intersectional multilevel analysis of individual heterogeneity and discriminatory accuracy approach, to describe mental health-related contacts across multiple interlocking social strata.^21^

Our results also show that relative rates across services are broadly consistent over social strata, with little evidence that specific population groups disproportionately use specific service types (e.g. emergency care). This is an important finding because it indicates systematic differences in NHS service use, likely underpinned by social determinants and requiring an upstream approach that addresses both underlying health needs and barriers to access. Consideration of how mental health presentations are defined and recognised is an important component of this. Applying a wider case definition for mental health-related hospital admissions substantially decreased the relative difference between groups, particularly for young people from Pakistani backgrounds. Other studies have shown that children from Pakistani and Bangladeshi backgrounds have a higher risk of internalising problems than their White peers.^39,40^ The perception and expression of symptoms therefore differ for individuals, which is important for policy makers to consider when evaluating evidence and, furthermore, for clinicians to be mindful of when making clinical judgements.^40^ However, the sensitivity and specificity of these undifferentiated stress presentations is unknown and needs further validation. Other research points to systemic differences in referral method to mental health services (indicative of punitive criminal justice responses to mental health problems) and length of stay in hospital (indicative of poorer care),^40,41^ highlighting the importance of considering multiple aspects of the journey to and through services. Future research using ECHILD could consider study designs that examine transitions across services to further examine pathways to care.

## Data Availability

The ECHILD database is made available for free for approved research based in the UK, via the ONS Secure Research Service. Enquiries to access the ECHILD database can be made by emailing. Researchers will need to be approved and submit a successful application to the ECHILD Data Access Committee and ONS Research Accreditation Panel to access the data, with strict statistical disclosure controls of all outputs of analyses

## Contributorship statement

RB and KL designed the study with input from HD, DL and JD. KL carried out the analysis and wrote the manuscript with supervision from RB. All authors contributed to revisions of the manuscript. KL and RB are guarantors of the paper.

## Data access statement

KL and RB had access to the raw data during this study. The ECHILD database is made available for free for approved research based in the UK, via the Office for National Statistics (ONS) Secure Research Service. Enquiries to access the ECHILD database can be made by emailing. Researchers will need to be approved and submit a successful application to the ECHILD Data Access Committee and ONS Research Accreditation Panel to access the data, with strict statistical disclosure controls of all outputs of analyses.

## Ethics and consent

Permissions to use linked, de-identified data from Hospital Episode Statistics and the National Pupil Database were granted by the Department for Education (DR200604.02B) and NHS Digital (DARS-NIC-381972). Ethical approval for the ECHILD project was granted by the National Research Ethics Service (17/LO/1494), NHS Health Research Authority Research Ethics Committee (20/EE/0180 and 21/SW/0159) and is overseen by the UCL Great Ormond Street Institute of Child Health’s Joint Research and Development Office (20PE16). Patient consent was not required to use the deidentified data in this study.

## Acknowledgements and disclosures

We gratefully acknowledge all children and families whose de-identified data are used in this research. We thank the CPRU Parent and Young Person Panels and the Bradford Institute for Health Research Mental Health Collaborative PPIE group for their contributions to this research. We also thank Constance Hobbs for her input into a draft version of this manuscript. We thank all the children, young people, parents and carers who contributed to the ECHILD project. We would like to acknowledge the contribution of the wider ECHILD Database support and programme management.

The ECHILD Database uses data from the Department for Education (DfE). The DfE does not accept responsibility for any inferences or conclusions derived by the authors. This work uses data provided by patients and collected by the National Health Service as part of their care and support. We are grateful to the ONS for providing the trusted research environment for the ECHILD Database. ONS agrees that the figures and descriptions of results in the attached document may be published. This does not imply ONS’ acceptance of the validity of the methods used to obtain these figures, or of any analysis of the results. The DfE, NHS England and ONS do not accept responsibility for any inferences or conclusions derived by the authors. The views in this publication do not necessarily reflect the views of UCL.

