## Supplementary Appendix for "Disparities in adolescent mental health-related National Health Service presentations in England: secondary school cohort study"

**Table S1.** Datasets from ECHILD used in this study

| Dataset | Common acronym | Data provider | Description | Key variables* | Years |
|---|---|---|---|---|---|
| **National pupil database school census** | NPD school census | Department for Education | Termly census with information for pupils in state-maintained educational settings in England, including special schools, maintained pupil referral units, alternative provision academies and free schools | Age at start of the academic year, ethnic group, gender, English as an additional language, special educational needs, MSOA | 2012/13-2022/23 |
| **NPD alternative provision census** | NPD AP census | Department for Education | Annual census with information for pupils in maintained pupil referral units, alternative provision academies and alternative provision free schools | Age at start of the academic year, ethnic group, gender, English as an additional language, special educational needs, MSOA | 2012/13-2022/23 |
| **NPD absences** | NPD absences | Department for Education | Data on the number of possible sessions and absences. Compiled from the NPD school censuses | Sessions possible, authorised absences, unauthorised absences | 2011/12-2021/22 |
| **NPD Children Looked After Return** | NPD CLA | Department for Education | National individual-level dataset, which contains information on all looked-after children and recent care leavers in England | Care episode start date, unaccompanied asylum seeker flag | 2012/13-2022/23 |
| **Get information about schools database** | GIAS | Department for Education | Opensource information about schools and colleges in England, including names, type, establishment group and governance (formerly Edubase). Linked to ECHILD using school URN. | Type of establishment, unique reference number | Cross-sectional data (with historical information included) as at 2023 |
| **National Statistics Postcode Lookup** | NSPL | ONS | Opensource geographical information mapping Census Output Areas to a range of higher statistical geographies | Region, MSOA | Cross-sectional data from 2012 to 2023 |

| | | | | | |
|---|---|---|---|---|---|
| **Hospital episode statistics admitted patient care** | HES APC | NHS England | Episode level data on inpatient and day case discharges from English NHS hospitals and English NHS commissioned activity in the independent sector | Diagnoses, episode start data, episode end date | 2016/17-2022/23 |
| **Emergency care data set** | ECDS | NHS England | Attendances at emergency departments and urgent care in English NHS hospitals and English NHS commissioned activity in the independent sector | Attendance, admission date, discharge dates, locations, presenting complaint, diagnoses | 2020/21-2022/23 |
| **Mental health services dataset** | MHSDS | NHS England | Holds patient-level data from the health records of children, young people and adults who are in contact in-patient, out-patient, and community mental health services | Referral rejection reason, Referral request received date, Service or team type referred to | 2016/17-2022/23 |
| **Mortality data** | | ONS | Information on the cause and date of deaths registered in England and Wales (including those that occur outside of hospital). | Date of death | 2011/12-2022/23 |

*Variables as described in the source dataset; ECHILD = Education and Child Health Insights from Linked Data; MSOA = middle super output area; NHS = National Health Service; ONS = Office for National Statistics; URN = unique reference number

**Follow-up year (1 September to 31 August)**

| Secondary school start year (1 September to 31 August) | 2012/13 | 2013/14 | 2014/15 | 2015/16 | 2016/17 | 2017/18 | 2018/19 | 2019/20 | 2020/21 | 2021/22 |
|---|---|---|---|---|---|---|---|---|---|---|
| **2012/13** | 11 (Y7) | 12 | 13 | 14 | 15 | 16 | 17 | 18 | 19 | 20 |
| **2013/14** | | 11 (Y7) | 12 | 13 | 14 | 15 | 16 | 17 | 18 | 19 |
| **2014/15** | | | 11 (Y7) | 12 | 13 | 14 | 15 | 16 | 17 | 18 |
| **2015/16** | | | | 11 (Y7) | 12 | 13 | 14 | 15 | 16 | 17 |
| **2016/17** | | | | | 11 (Y7) | 12 | 13 | 14 | 15 | 16 |
| **2017/18** | | | | | | 11 (Y7) | 12 | 13 | 14 | 15 |
| **2018/19** | | | | | | | 11 (Y7) | 12 | 13 | 14 |
| **2019/20** | | | | | | | | 11 (Y7) | 12 | 13 |
| **2020/21** | | | | | | | | | 11 (Y7) | 12 |
| **2021/22** | | | | | | | | | | 11 (Y7) |

HES APC (2012/13 to 2021/22)

MHSDS (2016/17 to 2021/22)

ECDS (2020/21 to 2021/22)

**Key**

Numbers in cells indicate expected age at 1 September

Brackets indicate school Year (Y)

Newly enrolled into cohort (light grey)

Follow up (dark grey)

**Figure S1.** Expected age of pupils at entry into each follow-up year: entry into study and dates of outcome datasets highlighted. ECDS = emergency care data set; HES APC = hospital episode statistics admitted patient care; MHSDS = mental health services dataset

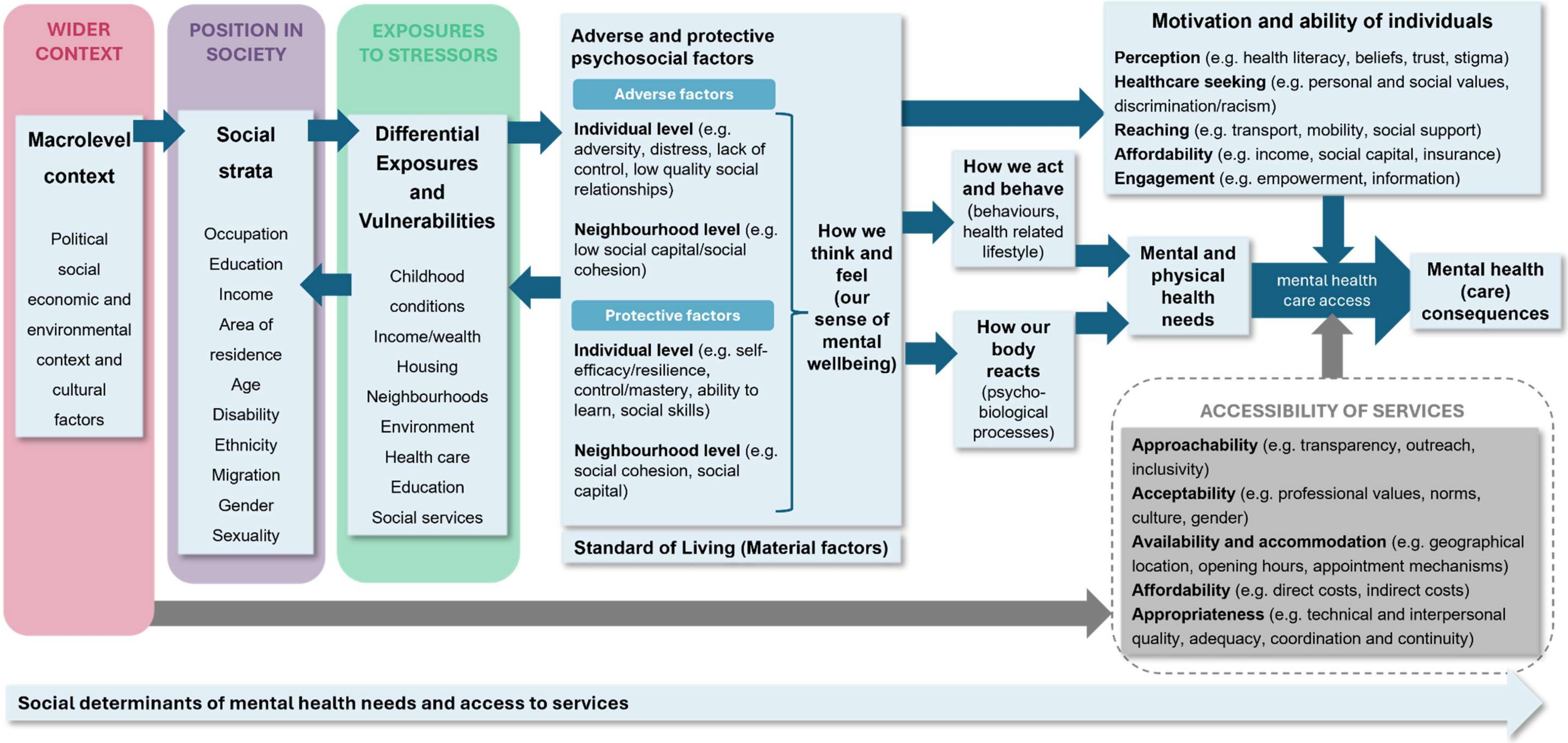


**Figure S2.** Conceptual diagram of the social determinants of mental health needs and service access. Adapted from: Public Health England & the UCL Institute of Health Equity, 2017; Levesque, Harris & Russell, 2013

**Table S2.** Mental health services dataset (MHSDS) variables and values used to define NHS-funded mental health service outcomes

| Outcome | Data dictionary name [variable name] / MHSDS table | Description | Rules applied [values] |
|---|---|---|---|
| **Referral - any** | Referral request received date [ReferralRequestReceivedDate] / MHS101 | This is the date the referral request was received by the Health Care Provider | Include referral requests received date is within specified follow-up time |
| | Service or team type referred to [ServTeamTypeRefToMH] / MHS102 | The type of service or team within a Mental Health Service that a patient was referred to | Include any |
| **Referral - neurodevelopmental concerns/ neurodiversity-related difficulties* ***** | Referral request received date [ReferralRequestReceivedDate] / MHS101 | This is the date the referral request was received by the Health Care Provider | Include referral requests received date is within specified follow-up time |
| | Primary reason for referral [PrimReasonReferralMH] / MHS101 | This is the primary presenting condition or symptom for which the patient was referred to a Mental Health Service | Must only include any of:<br>• Neurodevelopmental conditions, excluding autism [24]<br>• Suspected autism [25]<br>• Diagnosed autism [26]<br>• Behaviours that challenge due to a learning disability [30] |
| | Service or team type referred to [ServTeamTypeRefToMH] / MHS102 | The type of service or team within a Mental Health | Must only include any of:<br>• Autism services [C01]<br>• Neurodevelopmental team [C04]<br>• Enhanced/Intensive Support Service [E04] |
| **Referral - crisis** ***** | Primary reason for referral [PrimReasonReferralMH] / MHS101 | This is the primary presenting condition or symptom for which the patient was referred to a Mental Health Service | Include: In crisis [18] |
| | Source of referral [SourceOfReferralMH] / MHS101 | The source of referral to a Mental Health Service | Include: Acute Secondary Care: Emergency Care Department [H1] |

| | | | |
|---|---|---|---|
| | Referral request received date [ReferralRequestReceivedDate] / MHS101 | This is the date the referral request was received by the Health Care Provider | Include referral requests received date is within specified follow-up time |
| | Service or team type referred to [ServTeamTypeRefToMH] / MHS102 | The type of service or team within a Mental Health Service that a patient was referred to | Include any of:<br>• Crisis Resolution Team/Home Treatment Service [A02]<br>• Psychiatric Liaison Service [A11]<br>• 24/7 Crisis Response Line [A19]<br>• Health Based Place Of Safety Service [A20]<br>• Crisis Café/ SafeHaven/ Sanctuary Service [A21]<br>• Walk-in Crisis Assessment Unit Service [A22]<br>• Crisis House Service [A25]<br>• Paediatric Liaison Service [C05] |

*included if matches criteria for primary reason for referral or, if reason for referral is missing, criteria for service or team type referred to; **must meet all specified criteria to be included within definition; ***linked to MHS101 (referrals table) using “uniqservreqid_deid” and “person_id_deid”; NHS = National Health Service

**Table S3.** SNOMED CT codes used to define mental health-related emergency department attendances in the Emergency Care Data Set (ECDS), by NHS England definition group

| Definition group | | SNOMED CT Code | Description |
|---|---|---|---|
| **ECDS group 1: Psych / tox / D+A*** | Mental health | 33449004 | Personality disorder |
| | | 72366004 | Eating disorder |
| | | 191736004 | Obsessive-compulsive disorder |
| | | 371631005 | Panic disorder |
| | | 197480006 | Anxiety disorder |
| | | 35489007 | Depression |
| | | 13746004 | Bipolar disorder |
| | | 58214004 | Schizophrenia |
| | | 69322001 | Psychotic disorder |
| | | 44376007 | Dissociative disorder |
| | | 397923000 | Somatisation disorder |
| | | 30077003 | Somatoform pain disorder |
| | | 17226007 | Adjustment disorder |
| | | 50705009 | Factitious disorder |
| | Toxicology | 295124009 | Paracetamol overdose |
| | | 295217003 | Non-steroidal anti-inflammatory overdose |
| | | 295830007 | Overdose of antidepressant drug |
| | | 296015009 | Sedative overdose |
| | | 242253008 | Overdose of opiate |
| | | 296335002 | Overdose of beta-adrenergic blocking drug |
| | | 296355001 | Overdose of calcium-channel blockers |
| | | 297062008 | Digoxin overdose |
| | | 24354007 | Poisoning by colchicine |
| | | 111769007 | Poisoning by antihypertensive agent |
| | | 419347003 | Poisoning by antibacterial drug |
| | | 57005003 | Poisoning by caffeine |
| | | 1382631000000101 | Poisoning caused by household product |
| | | 275390005 | Toxic effect of plant |
| | | 44400004 | Toxic effect of venom |
| | | 43302000 | Anticoagulant overdosage |
| | | 1148733008 | Iron and/or iron compound overdose |
| | | 17383000 | Toxic effect of carbon monoxide |
| | | 66207005 | Toxic effect of cyanide |
| | | 38959009 | Methaemoglobinaemia |

| | | | |
|---|---|---|---|
| | | 67426006 | Toxic effect of alcohol |
| | | 371089000 | Serotonin syndrome |
| | | 1148734002 | Acetylcholine receptor antagonist poisoning |
| | | 61356009 | Poisoning by parasympathomimetic drug |
| | | 371043007 | Toxic inhalation injury |
| | | 426936004 | Smoke inhalation injury |
| | | 75478009 | Poisoning |
| | Drug / alcohol | 25702006 | Alcohol intoxication |
| | | 66590003 | Alcohol dependence |
| | | 191480000 | Alcohol withdrawal syndrome |
| | | 308742005 | Alcohol withdrawal-induced convulsion |
| | | 1098111000000101 | Drug-induced seizure |
| | | 307052004 | Illicit drug use |
| | | 268640002 | Hypnotic or anxiolytic dependence |
| | | 75544000 | Opioid dependence |
| **Psychosocial/ behaviour change reference set**** | | 248062006 | Self-harm |
| | | 366979004 | Depressive feelings |
| | | 6471006 | Feeling suicidal |
| | | 48694002 | Feeling anxious |
| | | 248020004 | Behaviour: unusual |
| | | 248004009 | Behaviour: violent |
| | | 24199005 | Behaviour: agitation |
| | | 7011001 | Hallucinations |
| | | 2073000 | Delusions |

*Code must be in the diagnosis field with the lowest sequence number, indicating primary diagnosis; **code in the chief complaint field; NHS = National Health Service; SNOMED CT = Systematized Nomenclature of Medicine Clinical Terms

**Table S4**. ICD-10 codes used to identify emergency hospital admissions* for mental health disorders and adversity-related injuries in hospital episode statistics admitted patient care

| Category | Sub-category | ICD-10 codes | ICD-10 Description |
|---|---|---|---|
| **Mental health disorders**** | | | |
| Internalising | Mood Disorders | F320 | Mild depressive episode |
| | | F321 | Moderate depressive episode |
| | | F322 | Severe depression without psychotic symptoms |
| | | F328 | Other depressive episodes |
| | | F329 | Depressive episode, unspecified |
| | | F330 | Recurrent depressive disorder, current episode mild |
| | | F331 | Recurrent depressive disorder, current episode moderate |
| | | F332 | Recurrent depressive disorder, current episode severe without psychotic symptoms |
| | | F333 | Recurrent depressive disorder, current episode severe with psychotic symptoms |
| | | F338 | Other recurrent depressive disorders |
| | | F339 | Recurrent depressive disorder, unspecified |
| | | F341 | Dysthymia |
| | Anxiety or fear-related, Obsessive-compulsive, & Dissociative disorders | F40 | Phobic anxiety disorders |
| | | F41 | Other anxiety disorders |
| | | F94.0 | Elective mutism |
| | | F42 | Obsessive-compulsive disorder |
| | | F44 | Dissociative [conversation] disorders |
| | | F45 | Somatoform disorders |
| | | F48 | Other neurotic disorders |
| | | F54 | Psychological and behavioural factors associated with disorders or diseases classified elsewhere |
| | Disorders specifically associated with stress | F43 | Reaction to severe stress, and adjustment disorders |
| | | F62 | Enduring personality changes, not attributable to brain damage and disease |
| | | F94.1 | Reactive attachment disorder of childhood |
| | | F94.2 | Disinhibited attachment disorder of childhood |
| | Eating disorders | F50 | Eating disorders |
| | Emotional disorders with onset usually occurring in childhood and adolescence | F93 | Emotional disorders with onset specific to childhood |
| | | F98 | Other behavioural and emotional disorders with onset usually occurring in childhood and adolescence |
| Externalising | Impulse control, | F63 | Habit and impulse disorders |
| | | F90 | Disturbance of activity and attention |

| | | | |
|---|---|---|---|
| | Disruptive behaviour or dissocial disorders | F91 | Conduct disorders |
| | | F92 | Mixed disorders of conduct and emotions |
| Thought Problems & Personality Disorders | Mood Disorders with mania or psychotic symptoms | F30 | Manic episode |
| | | F31 | Bipolar affective disorder |
| | | F323 | Severe depression with psychotic symptoms |
| | Schizophrenia or other primary psychotic disorders | F20 | Schizophrenia |
| | | F21 | Schizotypal disorder |
| | | F22 | Persistent delusional disorders |
| | | F23 | Acute and transient psychotic disorders |
| | | F24 | Induced delusional disorder |
| | | F25 | Schizoaffective disorders |
| | | F28 | Other nonorganic psychotic disorders |
| | | F29 | Unspecified nonorganic psychosis |
| | Disorders of adult personality and behaviour | F60 | Specific Personality Disorders |
| | | F68 | Other disorders of adult personality and behaviour |
| | | F69 | Unspecified disorder of adult personality and behaviour |
| **Adversity-related injuries*** ** | | | |
| Drug and/or alcohol misuse | | F11-16, F18-19 | Mental and behavioural disorders due to psychoactive substance use |
| | | R78.1-R78.5 | Findings of drugs and other substances, not normally found in blood |
| | | T36-T50 (excl T50.6) | Poisoning by drugs, medicaments and biological substances |
| | | Y10-Y14 | Poisoning, undetermined intent |
| | | Z50.3 | Drug rehabilitation |
| | | Z71.5 | Drug abuse counselling and surveillance |
| | | Z72.2 | Problems related to lifestyle, drug use |
| | | F18 | Mental and behavioural disorders due to use of volatile solvents |
| | | X40-X44 X46-X49 | Accidental poisoning by and exposure to noxious substances |
| | | X69 | Intentional self-poisoning by and exposure to other and unspecified chemicals and noxious substances |
| | | Y16-Y19 | Poisoning, undetermined intent |
| | | G40.5 | Special epileptic syndromes |
| | | Z04.0 | Examination for Blood-alcohol and blood-drug test |
| | | E24.4 | Alcohol-induced pseudo-Cushing syndrome |
| | | F10 | Mental and behavioural disorders due to use of alcohol |

| | | |
|---|---|---|
| | G31.2 | Degeneration of nervous system due to alcohol |
| | G62.1 | Alcoholic polyneuropathy |
| | G72.1 | Alcoholic polyneuropathy |
| | I42.6 | Alcoholic cardiomyopathy |
| | K29.2 | Alcoholic gastritis |
| | K70 | Alcoholic liver disease |
| | K85.2 | Alcohol-induced acute pancreatitis |
| | K86.0 | Alcohol-induced chronic pancreatitis |
| | O35.4 | Maternal care for (suspected) damage to foetus from alcohol |
| | R78.0 | Finding of alcohol in blood |
| | T51 | Toxic effect of alcohol |
| | X45 | Accidental poisoning by and exposure to alcohol |
| | Y15 | Poisoning by and exposure to alcohol, undetermined intent |
| | Y90 | Evidence of alcohol involvement determined by blood alcohol level |
| | Y91 | Evidence of alcohol involvement determined by level of intoxication |
| | Z50.2 | Alcohol rehabilitation |
| | Z71.4 | Alcohol abuse counselling and surveillance |
| | Z72.1 | Problems related to lifestyle, alcohol use |
| Violence | T74 | Maltreatment syndrome |
| | T73 | Effects of other deprivation (extreme neglect) |
| | Y06, Y07 | Perpetrator of neglect and other maltreatment syndromes |
| | Y04, Y05 | Assault by bodily force and sexual assault |
| | X85-Y03, Y08, Y09 | other type of assault |
| | Y20-Y34 | Events of undetermined intent |
| | Z04.5 | Examination and observation following other inflicted injury |
| | Z04.8 | Examination and observation for other reasons: request for expert evidence |
| Self Harm | X60-69 | Intentional self-poisoning |
| | X70-X84 | Intentional self-harm |
| | Z91.5 | Personal history of self-harm |

*Hospital admissions were created by linking finished consultant episodes into a continuous inpatient stay (including hospital transfers); **Diagnosis (in any position) accompanied by episode start date [epistart] within follow-up time and an emergency admission method [admimeth = 21-24, 28] in the first episode of the admission; ***In any diagnostic position accompanied by a primary diagnosis of injury (any ICD-10 code beginning with “S” or “T”, injuries, poisoning, and other consequences of external causes) and episode start date [epistart] within follow-up time and an emergency admission method [admimeth = 21-24, 28] in the first episode of the admission; ICD-10 = International Classification of Diseases 10th Revision

**Table S5.** ICD-10 codes used to identify admissions with potentially psychosomatic symptoms in hospital episode statistics admitted patient care (including medical and surgical exclusion codes)

| | ICD-10 codes | ICD-10 Description |
|---|---|---|
| **Potentially psychosomatic symptoms* **** | | |
| Abdominal/pelvic pain | R10 | Abdominal and pelvic pain |
| Headache | R51 | Headache |
| | G442 | Tension-type headache |
| Other pain | M54 | Panniculitis affecting regions of neck and back |
| | M626 | Muscle strain |
| | M796 | Pain in limb |
| | R52 | Acute pain |
| Circulatory/respiratory signs | R00 | Abnormalities of heart beat |
| | R03 | Abnormal blood pressure reading, without diagnosis |
| | R05 | Cough |
| | R06 | Abnormalities of breathing |
| | R07 | Pain in throat and chest |
| Digestive symptoms | R11-14 | Nausea and vomiting, Heartburn, Dysphagia, Flatulence and related conditions |
| | R194 | Change in bowel habit |
| Skin symptoms | R20-21 | Disturbances of skin sensation, rash and other nonspecific skin eruption |
| | R231 | Pallor |
| | R234 | Changes in skin texture |
| | R238 | Other and unspecified skin changes |
| Nervous/musculoskeletal symptoms | R25 | Abnormal involuntary movements |
| | R26 | Abnormalities of gait and mobility |
| | R27 | Other lack of co-ordination |
| | R29.2-29.4 | Abnormal reflex, Abnormal posture, Clicking hip |
| | R298 | Other and unspecified signs and symptoms involving nervous and musculoskeletal systems |
| Cognitive symptoms | R40.0-40.1 | Somnolence, Stupor |
| | R41-42 | Other symptoms and signs involving cognitive functions and awareness, Dizziness and giddiness |
| Malaise/Fatigue/Syncope | R53 | Malaise and fatigue |
| | R55 | Syncope and collapse |
| Other/general symptoms | R44 | Other symptoms and signs involving general sensations and perceptions |
| | R45 | Symptoms and signs involving emotional state |
| | R46 | Symptoms and signs involving appearance and behaviour |

| | | |
|---|---|---|
| | R47 | Dysphasia and aphasia |
| | R49 | Voice disturbances |
| | Z56.3-56.4 | Stressful work schedule, Discord with boss and workmates |
| | Z71.1 | Person with feared complaint in whom no diagnosis is made |
| | Z73.3 | Stress, not elsewhere classified |
| Sleep disorders | F51 | Nonorganic sleep disorders |
| | G47 | Disorders of initiating and maintaining sleep [insomnias] |
| **Medical and surgical exclusion codes** | | |
| Medical | A00-09 | Intestinal infectious diseases |
| | K52.0 | Gastroenteritis and colitis due to radiation |
| | K52.1 | Toxic gastroenteritis and colitis |
| | K52.9 | Noninfective gastroenteritis and colitis unspecified |
| | J09-10, J13-18 | Influenza and pneumonia |
| | N81, N83, N85-90 | Non-inflammatory disease of female genital tract<br>Female genital prolapse |
| | E28.2 | Polycystic ovarian syndrome |
| | N31 | Neuromuscular dysfunction of bladder not elsewhere classified |
| | N39.0 | Urinary tract infection site not specified |
| | K55 | Vascular disorders of intestine |
| | K56 | Paralytic ileus and intestinal obstruction without hernia |
| | G40 | Epilepsy |
| | G41 | Status epilepticus |
| | G45 | Transient cerebral ischaemic attacks and related syndromes |
| | G46 | Vascular syndromes of brain in cerebrovascular diseases |
| | I60-69 | Cerebrovascular disease |
| | C15-26 | Malignant neoplasms of digestive organs |
| | C30-39 | Malignant neoplasms of respiratory and intrathoracic organs |
| | C40-41 | Malignant neoplasms of bone and articular cartilage |
| | C45-49 | Malignant neoplasms of mesothelial and soft tissue |
| | C51-58 | Malignant neoplasms of female genital organs |
| | C64-68 | Malignant neoplasms of urinary tract |
| | C69-72 | Malignant neoplasms of eye brain and other parts of central nervous system |
| | C73-75 | Malignant neoplasms of thyroid and other endocrine glands |
| | K35-38 | Diseases of appendix |
| Surgical | Y75.2 | Laparoscopic approach to the abdominal cavity not elsewhere classified |

| | | |
|---|---|---|
| | H01.1 | Emergency excision of abnormal appendix and drainage HFQ |
| | H01.2 | Emergency excision of normal appendix not elsewhere classified |
| | H01.3 | Emergency excision of normal appendix |
| | H01.9 | Unspecified emergency excision of appendix |
| | H02.1 | Interval appendectomy |
| | H02.3 | Prophylactic appendectomy NEC |
| | H02.4 | Incidental appendectomy |
| | H02.8 | Other specified excision of appendix |
| | H02.9 | Unspecified excision of appendix |

*Diagnosis (in any diagnostic position) accompanied by episode start date [epistart] within follow-up time and an emergency admission method [admimeth = 21-24, 28] in the first episode of the admission; **Admissions were not categorised as potentially psychosomatic if a medical or surgical cause was indicated by an operation or subsidiary diagnostic code for the same admission; ICD-10 = International Classification of Diseases 10th Revision

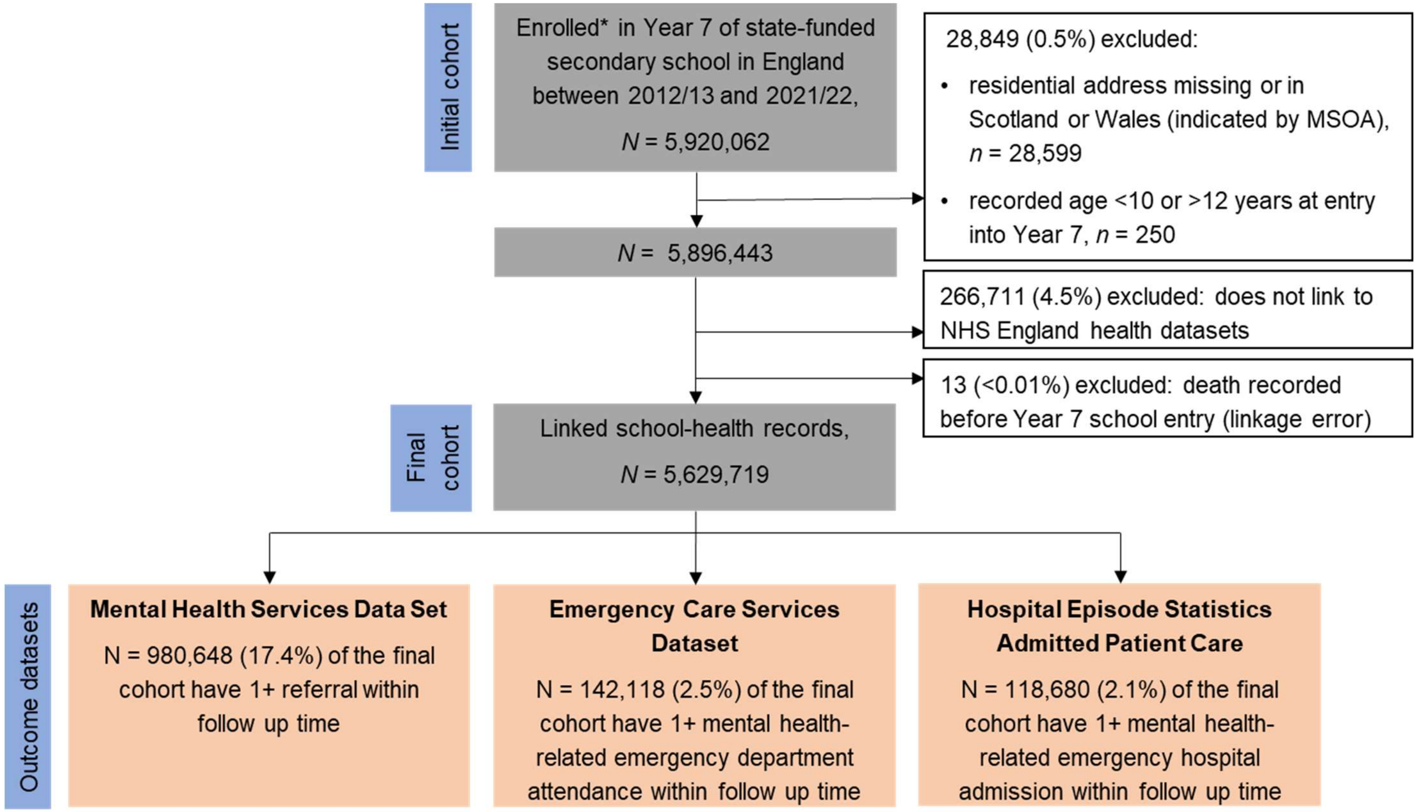


**Figure S3.** Flow diagram of cohort derivation and cohort appearance in outcome datasets; *Enrolment defined by appearance in January census of the School Census or the alternative provision census or pupil referral unit census; MSOA = middle layer super output area

**Table S6.** Linkage between the national pupil database and NHS-England datasets, by cohort characteristics

| | | **Total** N | **Linked** N (%) | **Not linked** N (%) |
|---|---|---|---|---|
| | Total | 5896443 | 5629732 (95.5) | 266711 (4.5) |
| Cohort (Year 7 entry) | 2012/13 | 534016 | 503359 (94.3) | 30657 (5.7) |
| | 2013/14 | 530854 | 503410 (94.8) | 27444 (5.2) |
| | 2014/15 | 552023 | 526665 (95.4) | 25358 (4.6) |
| | 2015/16 | 570233 | 542045 (95.1) | 28188 (4.9) |
| | 2016/17 | 582917 | 554167 (95.1) | 28750 (4.9) |
| | 2017/18 | 594928 | 566595 (95.2) | 28333 (4.8) |
| | 2018/19 | 614782 | 586178 (95.4) | 28604 (4.7) |
| | 2019/20 | 638689 | 612429 (95.9) | 26260 (4.1) |
| | 2020/21 | 633154 | 612140 (96.7) | 21014 (3.3) |
| | 2021/22 | 644847 | 622744 (96.6) | 22103 (3.4) |
| Gender | Male | 3017539 | 2886328 (95.7) | 131211 (4.4) |
| | Female | 2878904 | 2743404 (95.3) | 135500 (4.7) |
| Region of residence* | East Midlands | 513310 | 494000 (96.2) | 19310 (3.8) |
| | East of England | 664309 | 633420 (95.4) | 30885 (4.7) |
| | London | 879251 | 817260 (93.0) | 61990 (7.1) |
| | North East | 279511 | 271515 (97.1) | 7995 (2.9) |
| | North West | 808461 | 769030 (95.1) | 39435 (4.9) |
| | South East | 933727 | 895180 (95.9) | 38550 (4.1) |
| | South West | 553318 | 533180 (96.4) | 20135 (3.6) |
| | West Midlands | 663059 | 634915 (95.8) | 28145 (4.2) |
| | Yorkshire and the Humber | 601497 | 581230 (96.6) | 20265 (3.4) |
| IMD groups | 1 Most deprived 20% | 1480504 | 1409706 (95.2) | 70798 (4.8) |
| | 2 | 1196718 | 1141204 (95.4) | 55514 (4.6) |
| | 3 | 1079446 | 1031046 (95.5) | 48400 (4.5) |
| | 4 | 1031517 | 987215 (95.7) | 44302 (4.3) |
| | 5 Least deprived 20% | 1108258 | 1060561 (95.7) | 47697 (4.3) |
| FSM eligible | No | 4830203 | 4612972 (95.5) | 217231 (4.5) |
| | Yes | 1066240 | 1016760 (95.4) | 49480 (4.6) |
| Racial-ethnic group (minor) | Bangladeshi | 99809 | 93045 (93.2) | 6764 (6.8) |
| | Indian | 168835 | 159566 (94.5) | 9269 (5.5) |
| | Asian (other) | 101602 | 97120 (95.6) | 4482 (4.4) |
| | Pakistani | 252193 | 237382 (94.1) | 14811 (5.9) |
| | Black African | 218696 | 203439 (93.0) | 15257 (7.0) |
| | Black Caribbean | 72555 | 61104 (84.2) | 11451 (15.8) |
| | Black (other) | 42063 | 37927 (90.2) | 4136 (9.8) |
| | Chinese | 23060 | 21877 (94.9) | 1183 (5.1) |
| | Mixed (other) | 118202 | 110031 (93.1) | 8171 (6.9) |
| | Mixed Asian & White | 78253 | 74257 (94.9) | 3996 (5.1) |
| | Mixed Black African & White | 44676 | 41964 (93.9) | 2712 (6.1) |

| | | | | |
|---|---|---|---|---|
| | Mixed Black Caribbean & White | 91105 | 84974 (93.3) | 6131 (6.7) |
| | Other | 103977 | 98279 (94.5) | 5698 (5.5) |
| | Refused | 43336 | 39469 (91.1) | 3867 (8.9) |
| | White British | 4064670 | 3917148 (96.4) | 147522 (3.6) |
| | White Irish | 17594 | 16621 (94.5) | 973 (5.5) |
| | White Irish Traveller | 3484 | 3301 (94.8) | 183 (5.3) |
| | White (other) | 330378 | 311401 (94.3) | 18977 (5.7) |
| | White Romany Gypsy | 17847 | 17083 (95.7) | 764 (4.3) |
| | Missing | 4108 | 3744 (91.1) | 364 (8.9) |
| Recorded SEND provision | None | 4831458 | 4610725 (95.4) | 220735 (4.6) |
| | SEND support | 838764 | 802935 (95.7) | 35829 (4.3) |
| | EHCP | 226221 | 216074 (95.5) | 10147 (4.5) |
| Persistent absences | No | 5332305 | 5097450 (95.6) | 234855 (4.4) |
| | Yes | 420312 | 400470 (95.3) | 19842 (4.7) |
| | Missing | 143826 | 131812 (91.7) | 12014 (8.4) |
| Care experienced** | No | 5837050 | 5573340 (96) | 263710 (5) |
| | Yes | 59400 | 56400 (95) | 3000 (5) |

*Counts rounded to the nearest 5 as per statistical disclosure rules for sub-national numbers; **Counts are rounded to the nearest 10 and percentages to the nearest whole number as per statistical disclosure rules for data from the Children Looked After dataset; IMD = index of multiple deprivation; FSM = free school meal; SEND = special educational needs and disability

**Table S7.** First referral to mental health services (overall and crisis referrals), by social strata

| | | Referrals (all) | | Crisis referrals* | |
|---|---|---|---|---|---|
| | | N | N crisis (% of all referrals) | N | Rate per 1000 person-years (95% CI) |
| | Total | 980648 | 17486 (1.8) | 57204 | 2.36 (2.34, 2.37) |
| Gender | Male | 405483 | 5832 (1.4) | 16592 | 1.33 (1.31, 1.35) |
| | Female | 575165 | 11654 (2.0) | 40612 | 3.44 (3.41, 3.47) |
| Region of residence** | East Midlands | 83745 | 1865 (2.2) | 5970 | 2.81 (2.74, 2.88) |
| | East of England | 113545 | 1905 (1.7) | 7385 | 2.70 (2.64, 2.77) |
| | London | 120665 | 3375 (2.8) | 9365 | 2.66 (2.61, 2.72) |
| | North East | 58975 | 1115 (1.9) | 3800 | 3.24 (3.14, 3.34) |
| | North West | 141510 | 2915 (2.1) | 8715 | 2.63 (2.57, 2.68) |
| | South East | 162470 | 2190 (1.3) | 8210 | 2.13 (2.08, 2.17) |
| | South West | 89675 | 510 (0.6) | 1845 | 0.80 (0.76, 0.83) |
| | West Midlands | 113140 | 1395 (1.2) | 5105 | 1.86 (1.81, 1.91) |
| | Yorkshire and the Humber | 96930 | 2210 (2.3) | 6810 | 2.72 (2.65, 2.78) |
| IMD groups | 1 Most deprived 20% | 265864 | 5816 (2.2) | 17672 | 2.93 (2.89, 2.97) |
| | 2 | 210512 | 3936 (1.9) | 12793 | 2.61 (2.56, 2.65) |
| | 3 | 180981 | 2795 (1.5) | 9548 | 2.14 (2.10, 2.19) |
| | 4 | 162635 | 2452 (1.5) | 8595 | 2.01 (1.97, 2.05) |
| | 5 Least deprived 20% | 160656 | 2487 (1.5) | 8596 | 1.86 (1.82, 1.90) |
| FSM eligible | No | 723741 | 12766 (1.8) | 40956 | 2.03 (2.01, 2.05) |
| | Yes | 256907 | 4720 (1.8) | 16248 | 3.97 (3.91, 4.03) |
| Racial-ethnic group | Bangladeshi | 9060 | 345 (3.8) | 822 | 2.04 (1.91, 2.19) |
| | Indian | 10738 | 262 (2.4) | 655 | 0.98 (0.91, 1.06) |
| | Asian (other) | 7674 | 256 (3.3) | 600 | 1.48 (1.37, 1.60) |
| | Pakistani | 20098 | 486 (2.4) | 1245 | 1.23 (1.16, 1.30) |
| | Black African | 15900 | 720 (4.5) | 1457 | 1.71 (1.62, 1.80) |
| | Black Caribbean | 10690 | 309 (2.9) | 816 | 3.00 (2.80, 3.21) |
| | Black (other) | 4556 | 159 (3.5) | 362 | 2.28 (2.05, 2.52) |
| | Chinese | 1547 | 45 (2.9) | 92 | 1.05 (0.85, 1.28) |
| | Mixed (other) | 18333 | 416 (2.3) | 1312 | 2.98 (2.82, 3.14) |
| | Mixed Asian & White | 12190 | 221 (1.8) | 752 | 2.48 (2.31, 2.67) |
| | Mixed Black African & White | 7347 | 174 (2.4) | 536 | 3.20 (2.94, 3.49) |
| | Mixed Black Caribbean & White | 19836 | 422 (2.1) | 1398 | 3.90 (3.70, 4.11) |
| | Other | 10059 | 259 (2.6) | 623 | 1.55 (1.43, 1.67) |
| | Refused | 6219 | 125 (2.0) | 407 | 2.60 (2.36, 2.87) |
| | White British | 784222 | 12349 (1.6) | 43589 | 2.54 (2.52, 2.56) |
| | White Irish | 3111 | 53 (1.7) | 178 | 2.41 (2.08, 2.80) |
| | White Irish Traveller | 848 | 12 (1.4) | 43 | 3.04 (2.25, 4.09) |
| | White (other) | 35515 | 818 (2.3) | 2176 | 1.73 (1.66, 1.81) |
| | White Romany Gypsy | 2268 | 47 (2.1) | 120 | 1.64 (1.37, 1.96) |
| | Missing | 437 | 8 (1.8) | 21 | 1.33 (0.86, 2.03) |
| Chronic condition | No | 878951 | 16115 (1.8) | 51691 | 2.29 (2.27, 2.31) |
| | Yes | 101697 | 1371 (1.3) | 5513 | 3.25 (3.16, 3.33) |
| Recorded SEND provision | None | 697496 | 13643 (2.0) | 42332 | 2.13 (2.11, 2.15) |
| | SEN support | 211379 | 3219 (1.5) | 12039 | 3.40 (3.34, 3.46) |
| | EHCP | 71773 | 624 (0.9) | 2833 | 3.22 (3.10, 3.34) |
| Persistent absences | No | 842904 | 15410 (1.8) | 49260 | 2.24 (2.22, 2.26) |
| | Yes | 122795 | 1769 (1.4) | 6999 | 4.00 (3.90, 4.09) |
| | Missing | 14949 | 307 (2.1) | 945 | 1.77 (1.66, 1.89) |

| | | | | | |
|---|---|---|---|---|---|
| Care experienced*** | | | | 55300 | 2.30 (2.28, 2.32) |
| | No | 956270 | 17060 (2) | | |
| | Yes | 24370 | 430 (2) | 1910 | 7.88 (7.53, 8.24) |

*First crisis referral during follow up included in this definition, whereas column four ("N crisis (% of all referrals)") counts of the number of first referrals that are considered crisis (see Table S2); **Counts rounded to the nearest 5 as per statistical disclosure rules for sub-national numbers; ***Counts are rounded to the nearest 10 and percentages to the nearest whole number as per statistical disclosure rules for data from the Children Looked After dataset; IMD = index of multiple deprivation; FSM = free school meal; SEND = special educational needs and disability

**Table S8.** Year-adjusted rates of first referral to mental health services per 1000 person-years (95% confidence intervals): derived from age group-specific Poisson regression models with separate models for each group of social strata

| | | Age group | | |
|---|---|---|---|---|
| | | **11-<16** | **16-<18** | **18-<21** |
| | *N* people | 5,628,357 | 2,769,042 | 1,710,950 |
| | Overall | 50.7 (50.6, 50.8) | 42.0 (41.8, 42.2) | 23.6 (23.4, 23.8) |
| Gender | Male | 40.1 (40.0, 40.3) | 32.9 (32.7, 33.2) | 19.4 (19.2, 19.7) |
| | Female | 62.0 (61.8, 62.2) | 52.3 (52.0, 52.6) | 28.3 (28.1, 28.6) |
| Region | East Midlands | 48.3 (47.9, 48.7) | 41.6 (40.9, 42.2) | 24.5 (23.9, 25.1) |
| | East of England | 51.8 (51.4, 52.1) | 44.6 (44.0, 45.1) | 25.1 (24.6, 25.7) |
| | London | 41.4 (41.1, 41.7) | 37.6 (37.1, 38.1) | 19.8 (19.4, 20.3) |
| | North East | 68.3 (67.6, 68.9) | 49.4 (48.5, 50.4) | 23.5 (22.7, 24.2) |
| | North West | 55.3 (55.0, 55.6) | 41.5 (41.0, 42.0) | 24.3 (23.8, 24.7) |
| | South East | 53.4 (53.1, 53.7) | 44.3 (43.8, 44.8) | 23.2 (22.8, 23.6) |
| | South West | 48.1 (47.8, 48.5) | 42.6 (41.9, 43.2) | 21.6 (21.1, 22.1) |
| | West Midlands | 51.8 (51.4, 52.1) | 42.1 (41.5, 42.7) | 26.4 (25.8, 26.9) |
| | Yorkshire and The Humber | 47.9 (47.6, 48.3) | 39.1 (38.5, 39.7) | 25.0 (24.4, 25.5) |
| IMD groups | 1.Most deprived | 55.5 (55.3, 55.8) | 45.4 (45.0, 45.8) | 27.7 (27.3, 28.1) |
| | 2 | 54.4 (54.1, 54.7) | 44.1 (43.7, 44.6) | 25.3 (24.9, 25.7) |
| | 3 | 51.2 (50.9, 51.5) | 42.6 (42.1, 43.0) | 23.1 (22.7, 23.4) |
| | 4 | 47.4 (47.1, 47.6) | 40.0 (39.5, 40.4) | 21.7 (21.3, 22.1) |
| | 5.Least deprived | 43.0 (42.8, 43.3) | 37.0 (36.6, 37.4) | 19.2 (18.8, 19.5) |
| FSM eligible | No | 44.1 (44.0, 44.2) | 38.0 (37.8, 38.2) | 21.0 (20.8, 21.2) |
| | Yes | 85.2 (84.8, 85.6) | 64.7 (64.1, 65.3) | 37.6 (37.0, 38.1) |
| Racial-ethnic group (minor) | Bangladeshi | 24.3 (23.7, 24.9) | 26.8 (25.7, 28.0) | 17.0 (15.9, 18.1) |
| | Indian | 17.1 (16.7, 17.5) | 18.2 (17.5, 19.0) | 12.3 (11.6, 13.0) |
| | Asian (other) | 19.8 (19.3, 20.4) | 22.7 (21.6, 23.7) | 14.5 (13.5, 15.6) |
| | Pakistani | 21.3 (20.9, 21.7) | 22.8 (22.1, 23.4) | 15.3 (14.6, 15.9) |
| | Black African | 18.8 (18.4, 19.2) | 23.0 (22.3, 23.8) | 17.0 (16.2, 17.8) |

| | | | | |
|---|---|---|---|---|
| | Black Caribbean | 49.2 (48.1, 50.3) | 42.1 (40.4, 43.9) | 24.1 (22.6, 25.7) |
| | Black (other) | 31.8 (30.7, 32.9) | 33.1 (31.0, 35.2) | 20.9 (18.8, 23.0) |
| | Chinese | 18.3 (17.1, 19.4) | 21.3 (19.1, 23.5) | 13.7 (11.6, 15.8) |
| | Mixed (other) | 49.5 (48.6, 50.3) | 45.8 (44.3, 47.4) | 25.4 (24.0, 26.9) |
| | Mixed Asian & White | 47.6 (46.7, 48.6) | 45.1 (43.2, 46.9) | 24.3 (22.6, 26.0) |
| | Mixed Black African & White | 52.1 (50.7, 53.4) | 46.8 (44.2, 49.4) | 28.8 (26.2, 31.4) |
| | Mixed Black Caribbean & White | 71.8 (70.7, 72.9) | 57.7 (55.7, 59.6) | 34.8 (33.0, 36.7) |
| | Other | 27.4 (26.8, 28.0) | 29.6 (28.4, 30.9) | 16.1 (15.0, 17.3) |
| | Refused | 46.5 (45.2, 47.8) | 43.9 (41.3, 46.6) | 23.2 (20.8, 25.6) |
| | White British | 59.7 (59.6, 59.9) | 46.8 (46.5, 47.0) | 25.5 (25.3, 25.7) |
| | White Irish | 53.2 (51.0, 55.4) | 44.6 (41.1, 48.1) | 27.5 (24.2, 30.8) |
| | White Irish Traveller | 70.4 (64.6, 76.2) | 74.2 (63.3, 85.2) | 65.3 (52.9, 77.8) |
| | White (other) | 32.0 (31.6, 32.4) | 30.9 (30.2, 31.7) | 17.9 (17.2, 18.6) |
| | White Romany Gypsy | 35.8 (34.1, 37.6) | 35.2 (32.0, 38.4) | 20.2 (17.3, 23.2) |
| | Missing | 31.2 (27.6, 34.8) | 31.4 (25.2, 37.6) | 20.7 (14.8, 26.6) |

FSM = free school meals; IMD = index of multiple deprivation

**Table S9.** Year-adjusted incidence rate ratios of first referral to mental health services (95% confidence intervals), compared to the age- and social strata-specific mean rate: derived from age group-specific Poisson regression models with separate models for each group of social strata

| | | Age group | | |
|---|---|---|---|---|
| | | **11-<16** | **16-<18** | **18-<21** |
| | *N* people | 5,628,357 | 2,769,042 | 1,710,950 |
| Gender | Male | 0.81 (0.81, 0.81) | 0.80 (0.80, 0.81) | 0.84 (0.83, 0.84) |
| | Female | 1.25 (1.25, 1.25) | 1.28 (1.27, 1.28) | 1.22 (1.21, 1.23) |
| Region | East Midlands | 0.96 (0.95, 0.97) | 0.99 (0.98, 1.01) | 1.04 (1.02, 1.07) |
| | East of England | 1.03 (1.02, 1.03) | 1.06 (1.05, 1.08) | 1.07 (1.05, 1.09) |
| | London | 0.82 (0.82, 0.83) | 0.90 (0.89, 0.91) | 0.84 (0.83, 0.86) |
| | North East | 1.35 (1.34, 1.37) | 1.18 (1.16, 1.20) | 1.00 (0.97, 1.03) |
| | North West | 1.10 (1.09, 1.10) | 0.99 (0.98, 1.00) | 1.03 (1.01, 1.05) |
| | South East | 1.06 (1.05, 1.07) | 1.06 (1.05, 1.07) | 0.99 (0.97, 1.00) |
| | South West | 0.96 (0.95, 0.96) | 1.02 (1.00, 1.03) | 0.92 (0.90, 0.94) |
| | West Midlands | 1.03 (1.02, 1.03) | 1.00 (0.99, 1.02) | 1.12 (1.10, 1.14) |
| | Yorkshire and The Humber | 0.95 (0.94, 0.96) | 0.93 (0.92, 0.95) | 1.06 (1.04, 1.08) |
| IMD groups | 1 Most deprived | 1.10 (1.10, 1.10) | 1.08 (1.08, 1.09) | 1.19 (1.17, 1.20) |
| | 2 | 1.08 (1.07, 1.08) | 1.05 (1.04, 1.06) | 1.08 (1.07, 1.10) |
| | 3 | 1.01 (1.01, 1.02) | 1.02 (1.01, 1.03) | 0.99 (0.97, 1.00) |
| | 4 | 0.94 (0.93, 0.94) | 0.95 (0.95, 0.96) | 0.93 (0.91, 0.94) |
| | 5 Least deprived | 0.85 (0.85, 0.86) | 0.88 (0.87, 0.89) | 0.82 (0.81, 0.83) |
| FSM eligible | No | 0.90 (0.89, 0.90) | 0.92 (0.92, 0.93) | 0.91 (0.91, 0.92) |
| | Yes | 1.73 (1.72, 1.74) | 1.57 (1.56, 1.59) | 1.64 (1.61, 1.66) |
| Racial-ethnic group (minor) | Bangladeshi | 0.51 (0.50, 0.53) | 0.66 (0.63, 0.69) | 0.73 (0.69, 0.78) |
| | Indian | 0.36 (0.35, 0.37) | 0.45 (0.43, 0.47) | 0.53 (0.50, 0.56) |
| | Asian (other) | 0.42 (0.41, 0.43) | 0.56 (0.53, 0.58) | 0.63 (0.59, 0.67) |
| | Pakistani | 0.45 (0.44, 0.46) | 0.56 (0.54, 0.58) | 0.66 (0.63, 0.69) |
| | Black African | 0.40 (0.39, 0.40) | 0.57 (0.55, 0.58) | 0.73 (0.70, 0.77) |
| | Black Caribbean | 1.04 (1.02, 1.06) | 1.04 (0.99, 1.08) | 1.04 (0.98, 1.11) |

| | | | | |
|---|---|---|---|---|
| | Black (other) | 0.67 (0.65, 0.70) | 0.81 (0.76, 0.87) | 0.90 (0.82, 1.00) |
| | Chinese | 0.39 (0.36, 0.41) | 0.52 (0.47, 0.58) | 0.59 (0.51, 0.69) |
| | Mixed (other) | 1.04 (1.03, 1.06) | 1.13 (1.09, 1.17) | 1.10 (1.04, 1.16) |
| | Mixed Asian & White | 1.01 (0.99, 1.03) | 1.11 (1.06, 1.16) | 1.05 (0.98, 1.13) |
| | Mixed Black African & White | 1.10 (1.07, 1.13) | 1.15 (1.09, 1.22) | 1.24 (1.14, 1.36) |
| | Mixed Black Caribbean & White | 1.52 (1.49, 1.54) | 1.42 (1.37, 1.47) | 1.51 (1.43, 1.59) |
| | Other | 0.58 (0.56, 0.59) | 0.73 (0.70, 0.76) | 0.70 (0.65, 0.75) |
| | Refused | 0.98 (0.95, 1.01) | 1.08 (1.02, 1.15) | 1.00 (0.91, 1.11) |
| | White British | 1.26 (1.26, 1.26) | 1.15 (1.15, 1.15) | 1.10 (1.10, 1.11) |
| | White Irish | 1.12 (1.08, 1.17) | 1.10 (1.01, 1.19) | 1.19 (1.06, 1.34) |
| | White Irish Traveller | 1.49 (1.37, 1.61) | 1.83 (1.58, 2.12) | 2.83 (2.34, 3.42) |
| | White (other) | 0.68 (0.67, 0.68) | 0.76 (0.74, 0.78) | 0.78 (0.75, 0.81) |
| | White Romany Gypsy | 0.76 (0.72, 0.79) | 0.87 (0.79, 0.95) | 0.88 (0.76, 1.01) |
| | Missing | 0.66 (0.59, 0.74) | 0.77 (0.63, 0.94) | 0.90 (0.67, 1.19) |

FSM = free school meals; IMD = index of multiple deprivation

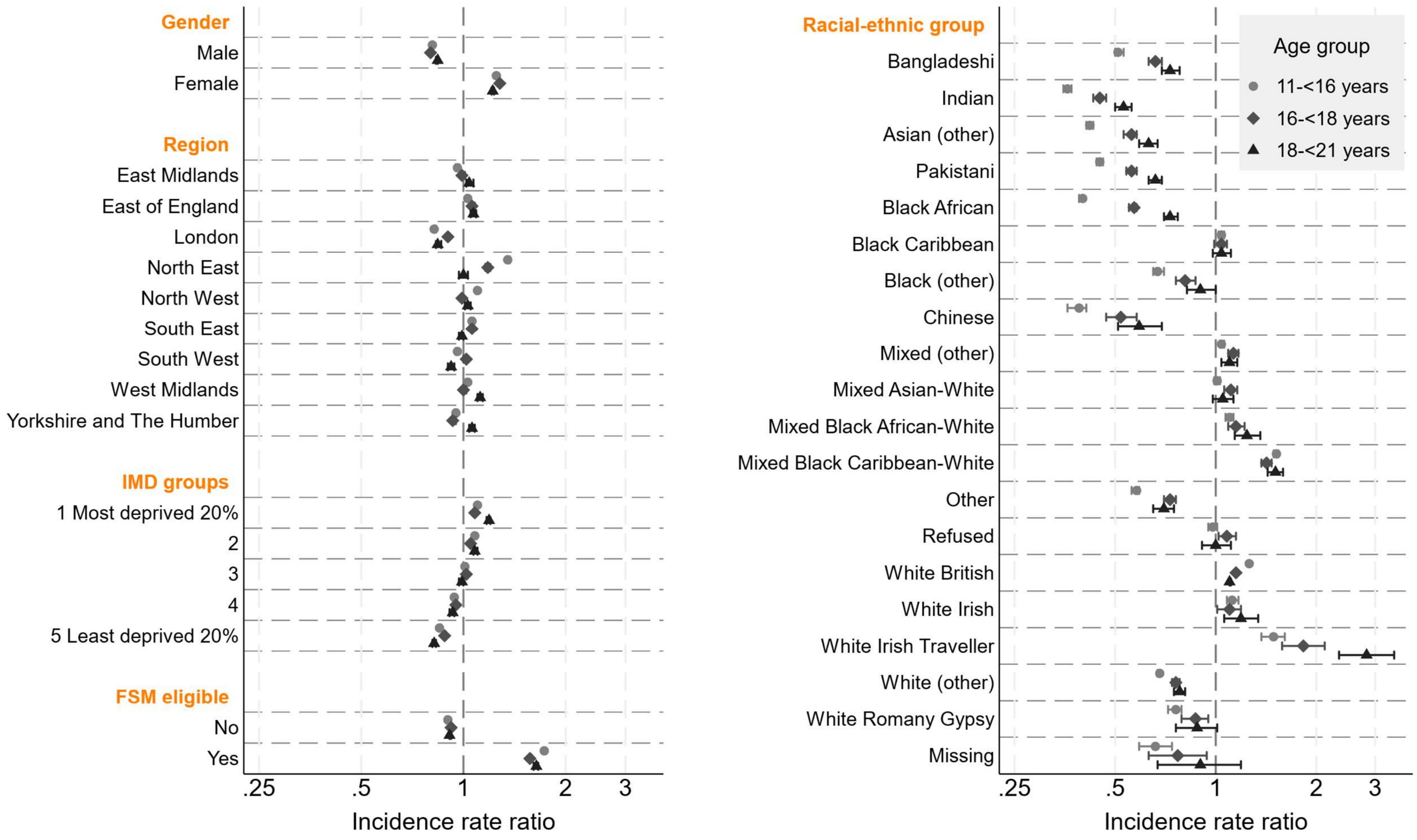


**Figure S4.** Year-adjusted incidence rate ratios (compared to the age- and social strata-specific mean rate) of first referral to mental health services with 95% confidence intervals: derived from age group-specific Poisson regression models with separate models for each group of social strata. FSM = free school meals; IMD = index of multiple deprivation.

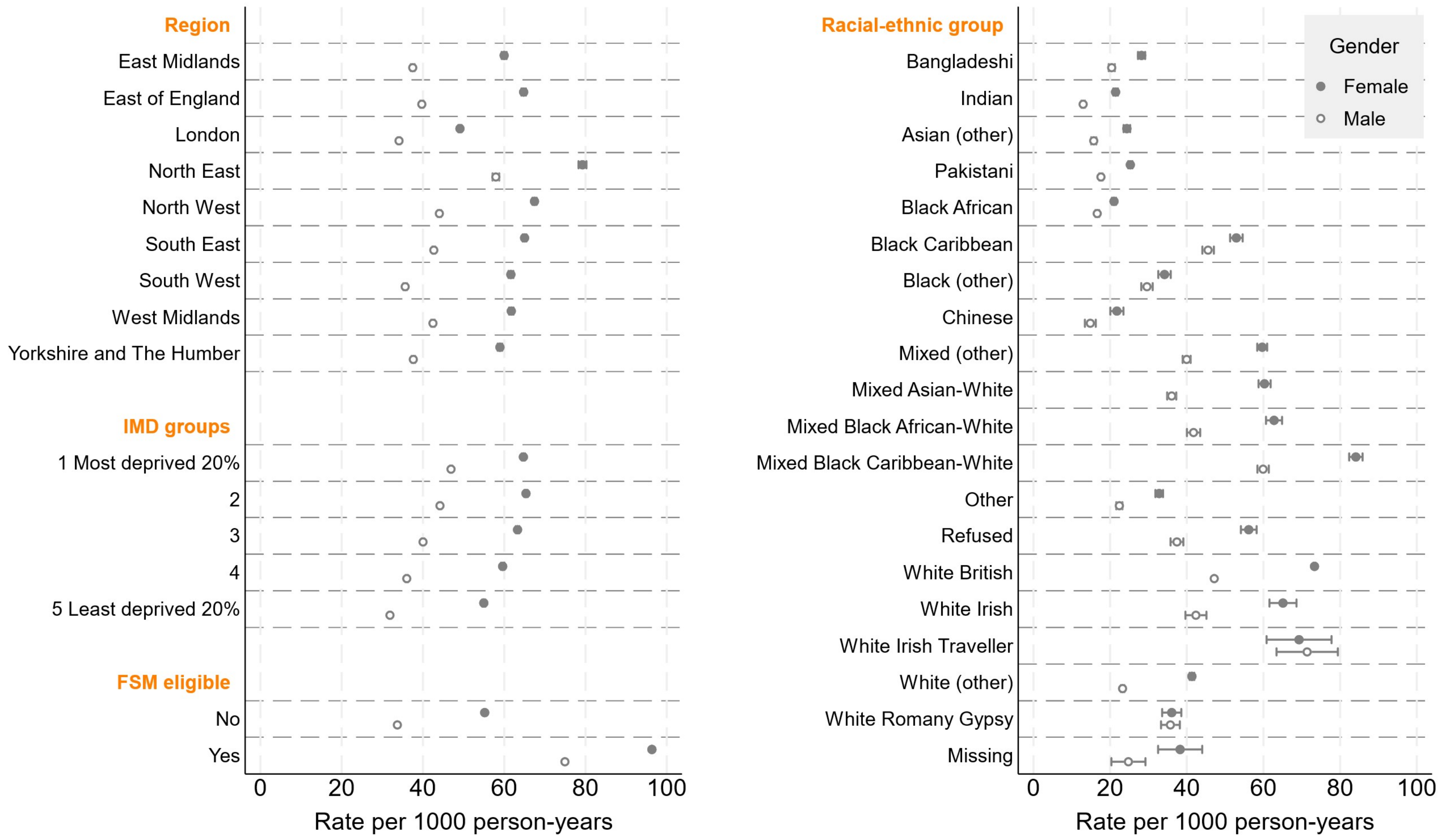
Region
East Midlands
East of England
London
North East
North West
South East
South West
West Midlands
Yorkshire and The Humber
IMD groups
1 Most deprived 20%
2
3
4
5 Least deprived 20%
FSM eligible
No
Yes
0
20
40
60
80
100
Rate per 1000 person-years
Racial-ethnic group
Bangladeshi
Indian
Asian (other)
Pakistani
Black African
Black Caribbean
Black (other)
Chinese
Mixed (other)
Mixed Asian-White
Mixed Black African-White
Mixed Black Caribbean-White
Other
Refused
White British
White Irish
White Irish Traveller
White (other)
White Romany Gypsy
Missing
Gender
Female
Male
0
20
40
60
80
100
Rate per 1000 person-years

**Figure S5.** Year-adjusted rates of first referral to mental health services per 1000 person-years: derived from separate Poisson regression regression models for each group of social strata and gender (11<16 years age group). FSM = free school meals; IMD = index of multiple deprivation.

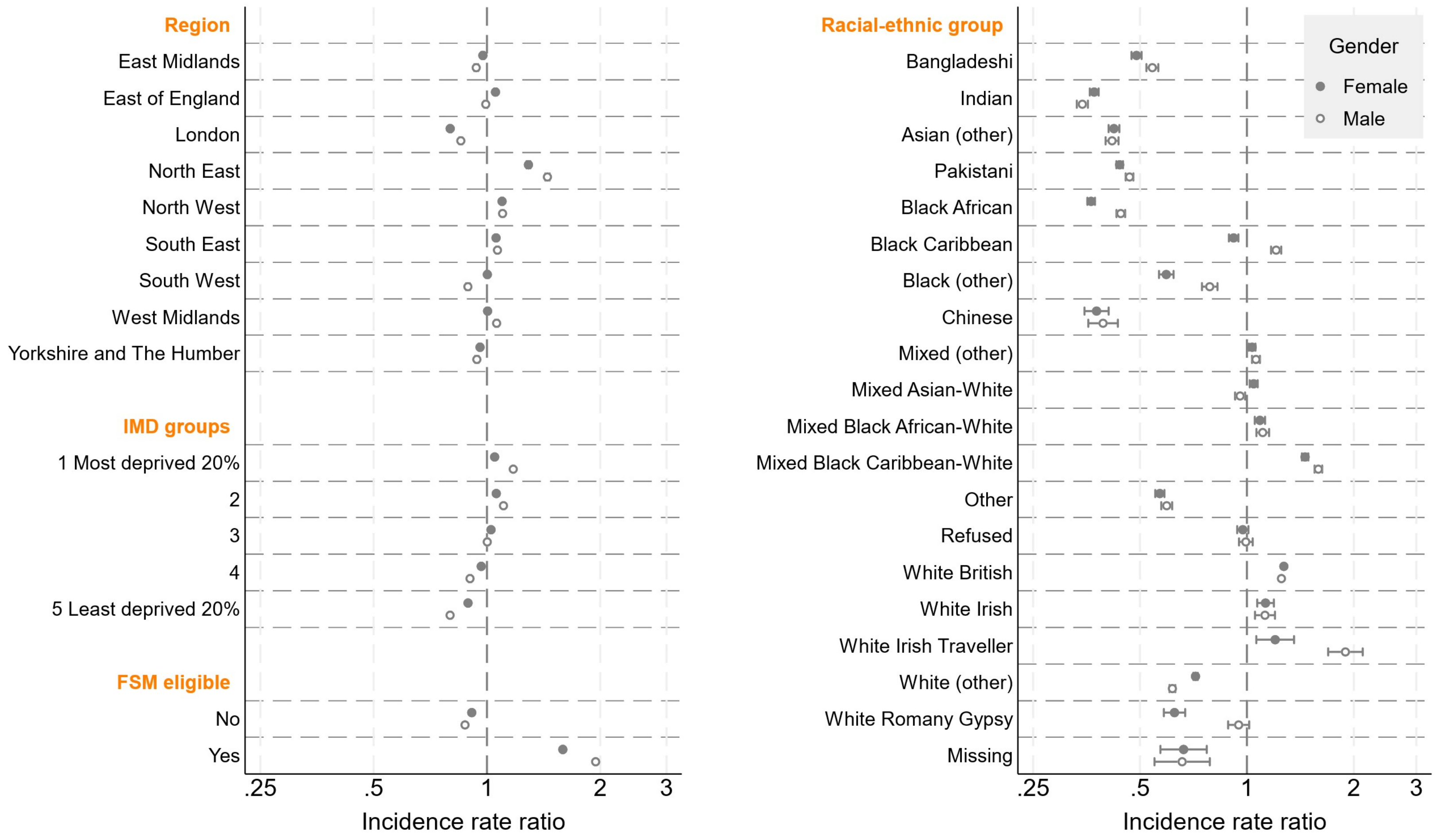
Region
East Midlands
East of England
London
North East
North West
South East
South West
West Midlands
Yorkshire and The Humber
IMD groups
1 Most deprived 20%
2
3
4
5 Least deprived 20%
FSM eligible
No
Yes
.25
.5
1
2
3
Incidence rate ratio
Racial-ethnic group
Bangladeshi
Indian
Asian (other)
Pakistani
Black African
Black Caribbean
Black (other)
Chinese
Mixed (other)
Mixed Asian-White
Mixed Black African-White
Mixed Black Caribbean-White
Other
Refused
White British
White Irish
White Irish Traveller
White (other)
White Romany Gypsy
Missing
Gender
Female
Male
.25
.5
1
2
3
Incidence rate ratio

**Figure S6.** Year-adjusted incidence rate ratios of first referral to mental health services (compared to the gender- and social strata-specific mean rate): derived from seperate Poisson regression regression models for each group of social strata and gender (11<16 years age group). FSM = free school meals; IMD = index of multiple deprivation.

Emergency department attendances

**Table S10.** Year-adjusted rates of first mental health-related emergency department attendances (95% confidence intervals): derived from age group-specific Poisson regression models with separate models for each group of social strata

| | | Age group | | |
|---|---|---|---|---|
| | | **11-<16** | **16-<18** | **18-<21** |
| | *N* people | 3,553,224 | 2,167,870 | 2,055,710 |
| | Overall | 11.2 (11.2, 11.3) | 15.9 (15.7, 16.1) | 17.4 (17.2, 17.5) |
| Gender | Male | 5.1 (5.0, 5.2) | 9.9 (9.7, 10.1) | 12.8 (12.6, 13.0) |
| | Female | 17.7 (17.5, 17.9) | 22.3 (22.0, 22.6) | 22.3 (22.1, 22.6) |
| Region | East Midlands | 12.0 (11.7, 12.3) | 17.7 (17.1, 18.3) | 18.6 (18.1, 19.2) |
| | East of England | 10.4 (10.2, 10.7) | 15.0 (14.5, 15.5) | 16.8 (16.4, 17.3) |
| | London | 9.1 (8.9, 9.3) | 13.5 (13.1, 13.9) | 14.6 (14.3, 15.0) |
| | North East | 12.4 (11.9, 12.8) | 17.7 (16.9, 18.5) | 19.3 (18.5, 20.0) |
| | North West | 13.6 (13.3, 13.9) | 16.7 (16.3, 17.2) | 18.7 (18.3, 19.2) |
| | South East | 11.2 (10.9, 11.4) | 16.3 (15.9, 16.8) | 17.3 (16.9, 17.7) |
| | South West | 10.3 (10.0, 10.6) | 16.1 (15.5, 16.6) | 17.2 (16.7, 17.7) |
| | West Midlands | 11.7 (11.4, 12.0) | 15.7 (15.2, 16.2) | 17.7 (17.2, 18.1) |
| | Yorkshire and The Humber | 11.4 (11.1, 11.7) | 16.4 (15.8, 16.9) | 18.0 (17.5, 18.5) |
| IMD groups | 1.Most deprived | 12.7 (12.5, 12.9) | 17.9 (17.5, 18.2) | 19.3 (19.0, 19.6) |
| | 2 | 12.5 (12.3, 12.7) | 17.4 (17.0, 17.8) | 18.9 (18.6, 19.3) |
| | 3 | 11.0 (10.8, 11.2) | 16.1 (15.7, 16.5) | 17.3 (16.9, 17.6) |
| | 4 | 10.1 (9.9, 10.3) | 14.3 (13.9, 14.6) | 16.1 (15.7, 16.5) |
| | 5.Least deprived | 9.2 (9.0, 9.3) | 13.1 (12.8, 13.5) | 14.7 (14.4, 15.0) |
| FSM eligible | No | 9.9 (9.8, 10.0) | 14.1 (13.9, 14.3) | 15.7 (15.6, 15.9) |

| | | | | |
|---|---|---|---|---|
| | Yes | 16.9 (16.7, 17.2) | 26.1 (25.5, 26.6) | 25.5 (25.0, 25.9) |
| Racial-ethnic group (minor) | Bangladeshi | 6.4 (5.9, 6.9) | 9.2 (8.2, 10.2) | 9.7 (8.8, 10.6) |
| | Indian | 4.2 (3.9, 4.5) | 6.9 (6.2, 7.6) | 9.1 (8.4, 9.8) |
| | Asian (other) | 5.5 (5.0, 6.0) | 8.6 (7.6, 9.6) | 11.0 (10.0, 12.0) |
| | Pakistani | 5.4 (5.1, 5.7) | 8.1 (7.5, 8.6) | 9.6 (9.0, 10.1) |
| | Black African | 5.9 (5.6, 6.3) | 10.1 (9.4, 10.8) | 13.1 (12.4, 13.9) |
| | Black Caribbean | 10.7 (9.8, 11.5) | 14.3 (12.8, 15.8) | 16.9 (15.5, 18.3) |
| | Black (other) | 8.8 (7.9, 9.8) | 12.6 (10.8, 14.4) | 16.3 (14.3, 18.2) |
| | Chinese | 4.1 (3.3, 4.9) | 8.9 (6.8, 11.1) | 8.2 (6.4, 10.0) |
| | Mixed (other) | 11.4 (10.8, 12.0) | 17.9 (16.6, 19.3) | 19.2 (17.9, 20.5) |
| | Mixed Asian & White | 11.4 (10.7, 12.2) | 16.5 (15.0, 18.1) | 19.2 (17.6, 20.8) |
| | Mixed Black African & White | 14.0 (12.9, 15.1) | 19.5 (17.2, 21.7) | 22.7 (20.3, 25.1) |
| | Mixed Black Caribbean & White | 16.3 (15.4, 17.2) | 23.9 (22.1, 25.6) | 23.3 (21.8, 24.8) |
| | Other | 7.2 (6.7, 7.7) | 10.5 (9.5, 11.6) | 11.7 (10.7, 12.8) |
| | Refused | 10.6 (9.7, 11.6) | 16.9 (14.7, 19.1) | 17.9 (15.7, 20.1) |
| | White British | 12.8 (12.7, 13.0) | 17.6 (17.4, 17.8) | 18.8 (18.6, 19.0) |
| | White Irish | 13.0 (11.1, 14.8) | 17.3 (14.2, 20.5) | 16.8 (14.1, 19.5) |
| | White Irish Traveller | 11.5 (7.7, 15.2) | 23.5 (14.8, 32.2) | 29.3 (21.0, 37.5) |
| | White (other) | 8.4 (8.1, 8.7) | 12.0 (11.4, 12.7) | 13.5 (12.8, 14.1) |
| | White Romany Gypsy | 7.9 (6.6, 9.3) | 14.3 (11.5, 17.2) | 12.8 (10.3, 15.3) |
| | Missing | 8.2 (5.1, 11.2) | 5.2 (1.3, 9.1) | 18.6 (12.5, 24.6) |

FSM = free school meals; IMD = index of multiple deprivation

**Table S11.** Year-adjusted incidence rate ratios of first mental health-related emergency department attendance (95% confidence intervals), compared to the age- and social strata-specific mean rate: derived from age group-specific Poisson regression models with separate models for each group of social strata

| | | Age group | | |
|---|---|---|---|---|
| | | **11-<16** | **16-<18** | **18-<21** |
| Gender | Male | 0.55 (0.54, 0.55) | 0.66 (0.65, 0.66) | 0.75 (0.74, 0.75) |
| | Female | 1.89 (1.87, 1.91) | 1.56 (1.54, 1.59) | 1.37 (1.35, 1.38) |
| Region | East Midlands | 1.08 (1.05, 1.10) | 1.13 (1.09, 1.17) | 1.08 (1.05, 1.11) |
| | East of England | 0.93 (0.91, 0.96) | 0.94 (0.91, 0.97) | 0.97 (0.94, 1.00) |
| | London | 0.81 (0.80, 0.83) | 0.84 (0.81, 0.86) | 0.83 (0.81, 0.85) |
| | North East | 1.11 (1.07, 1.14) | 1.13 (1.08, 1.19) | 1.12 (1.08, 1.17) |
| | North West | 1.22 (1.20, 1.24) | 1.06 (1.03, 1.09) | 1.09 (1.06, 1.11) |
| | South East | 1.00 (0.98, 1.02) | 1.03 (1.01, 1.06) | 1.00 (0.98, 1.02) |
| | South West | 0.92 (0.90, 0.94) | 1.01 (0.98, 1.05) | 0.99 (0.96, 1.02) |
| | West Midlands | 1.05 (1.02, 1.07) | 0.99 (0.96, 1.02) | 1.02 (0.99, 1.05) |
| | Yorkshire and The Humber | 1.02 (1.00, 1.05) | 1.03 (1.00, 1.07) | 1.04 (1.01, 1.07) |
| IMD groups | 1.Most deprived | 1.14 (1.12, 1.15) | 1.14 (1.12, 1.16) | 1.12 (1.11, 1.14) |
| | 2 | 1.12 (1.10, 1.14) | 1.11 (1.08, 1.13) | 1.10 (1.08, 1.12) |
| | 3 | 0.99 (0.97, 1.01) | 1.02 (0.99, 1.04) | 1.00 (0.98, 1.02) |
| | 4 | 0.91 (0.89, 0.92) | 0.89 (0.87, 0.92) | 0.93 (0.91, 0.95) |
| | 5.Least deprived | 0.82 (0.80, 0.83) | 0.82 (0.80, 0.84) | 0.84 (0.82, 0.86) |
| FSM eligible | No | 0.90 (0.90, 0.91) | 0.90 (0.90, 0.91) | 0.92 (0.91, 0.92) |
| | Yes | 1.54 (1.52, 1.56) | 1.76 (1.71, 1.80) | 1.54 (1.51, 1.57) |
| Racial-ethnic group (minor) | Bangladeshi | 0.59 (0.55, 0.64) | 0.58 (0.52, 0.64) | 0.55 (0.50, 0.61) |
| | Indian | 0.39 (0.36, 0.42) | 0.43 (0.39, 0.47) | 0.52 (0.48, 0.56) |
| | Asian (other) | 0.51 (0.47, 0.56) | 0.54 (0.48, 0.60) | 0.63 (0.57, 0.69) |
| | Pakistani | 0.50 (0.48, 0.53) | 0.50 (0.47, 0.54) | 0.54 (0.51, 0.58) |
| | Black African | 0.55 (0.52, 0.58) | 0.63 (0.59, 0.68) | 0.75 (0.71, 0.80) |

| | | | | |
|---|---|---|---|---|
| | Black Caribbean | 0.99 (0.91, 1.08) | 0.92 (0.82, 1.03) | 0.99 (0.91, 1.08) |
| | Black (other) | 0.82 (0.74, 0.91) | 0.80 (0.69, 0.94) | 0.95 (0.84, 1.08) |
| | Chinese | 0.38 (0.31, 0.47) | 0.56 (0.43, 0.72) | 0.46 (0.37, 0.58) |
| | Mixed (other) | 1.06 (1.00, 1.12) | 1.17 (1.08, 1.27) | 1.14 (1.06, 1.23) |
| | Mixed Asian & White | 1.06 (1.00, 1.14) | 1.07 (0.97, 1.19) | 1.14 (1.04, 1.25) |
| | Mixed Black African & White | 1.30 (1.21, 1.41) | 1.28 (1.12, 1.45) | 1.36 (1.21, 1.53) |
| | Mixed Black Caribbean & White | 1.52 (1.44, 1.61) | 1.62 (1.49, 1.75) | 1.40 (1.31, 1.51) |
| | Other | 0.67 (0.62, 0.72) | 0.67 (0.60, 0.74) | 0.67 (0.61, 0.74) |
| | Refused | 0.99 (0.90, 1.08) | 1.10 (0.95, 1.26) | 1.05 (0.92, 1.20) |
| | White British | 1.20 (1.19, 1.20) | 1.15 (1.14, 1.16) | 1.11 (1.11, 1.12) |
| | White Irish | 1.21 (1.05, 1.39) | 1.13 (0.93, 1.38) | 0.99 (0.83, 1.18) |
| | White Irish Traveller | 1.07 (0.77, 1.49) | 1.61 (1.06, 2.44) | 1.80 (1.31, 2.46) |
| | White (other) | 0.78 (0.75, 0.81) | 0.77 (0.72, 0.81) | 0.78 (0.74, 0.82) |
| | White Romany Gypsy | 0.74 (0.62, 0.87) | 0.93 (0.75, 1.15) | 0.73 (0.60, 0.90) |
| | Missing | 0.76 (0.52, 1.10) | 0.32 (0.15, 0.69) | 1.09 (0.77, 1.55) |

FSM = free school meals; IMD = index of multiple deprivation

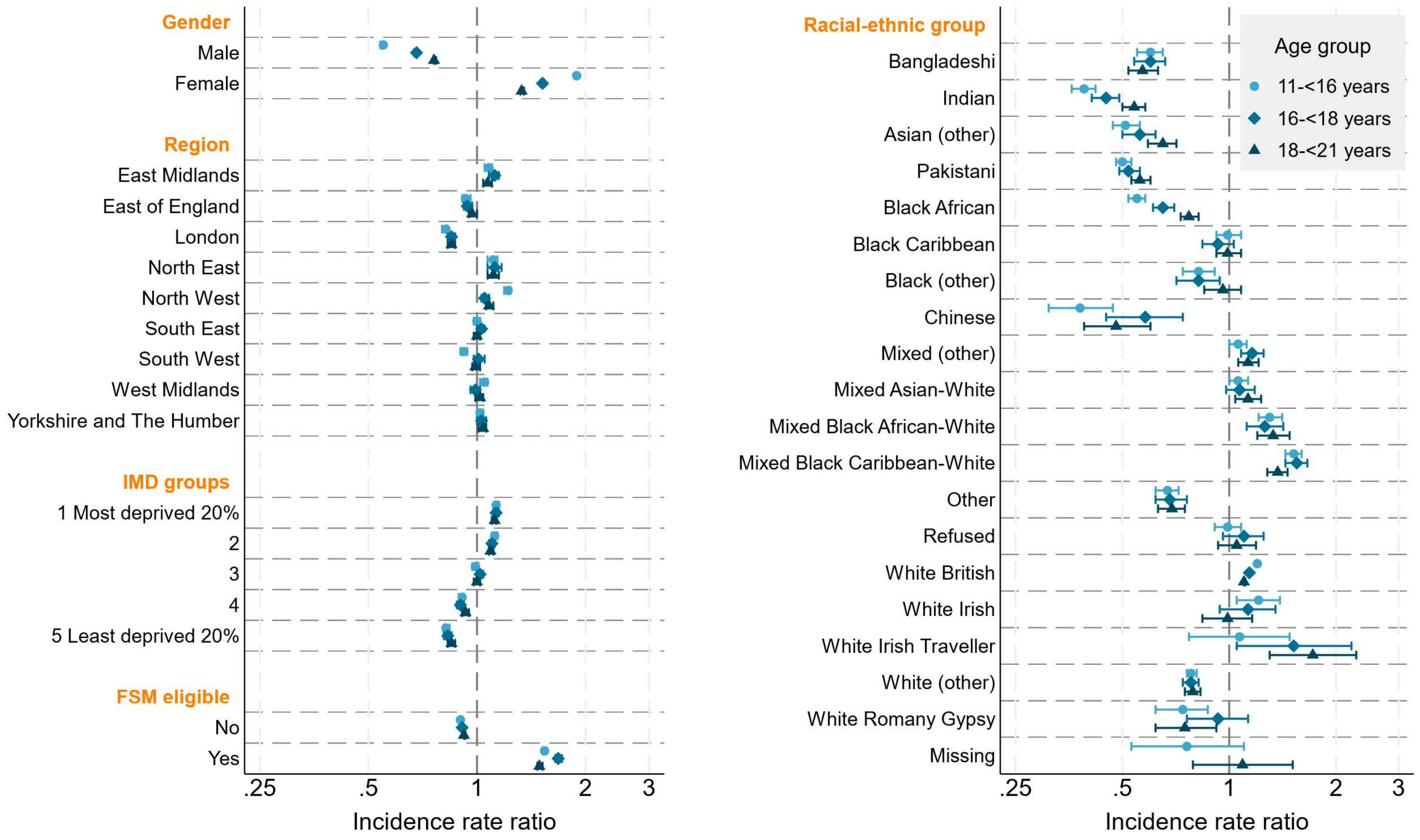


**Figure S7.** Year-adjusted incidence rate ratios (compared to the age- and social strata-specific mean rate) of first mental health-related emergency department attendance with 95% confidence intervals: derived from age group-specific Poisson regression models with separate models for each group of social strata. FSM = free school meals; IMD = index of multiple deprivation

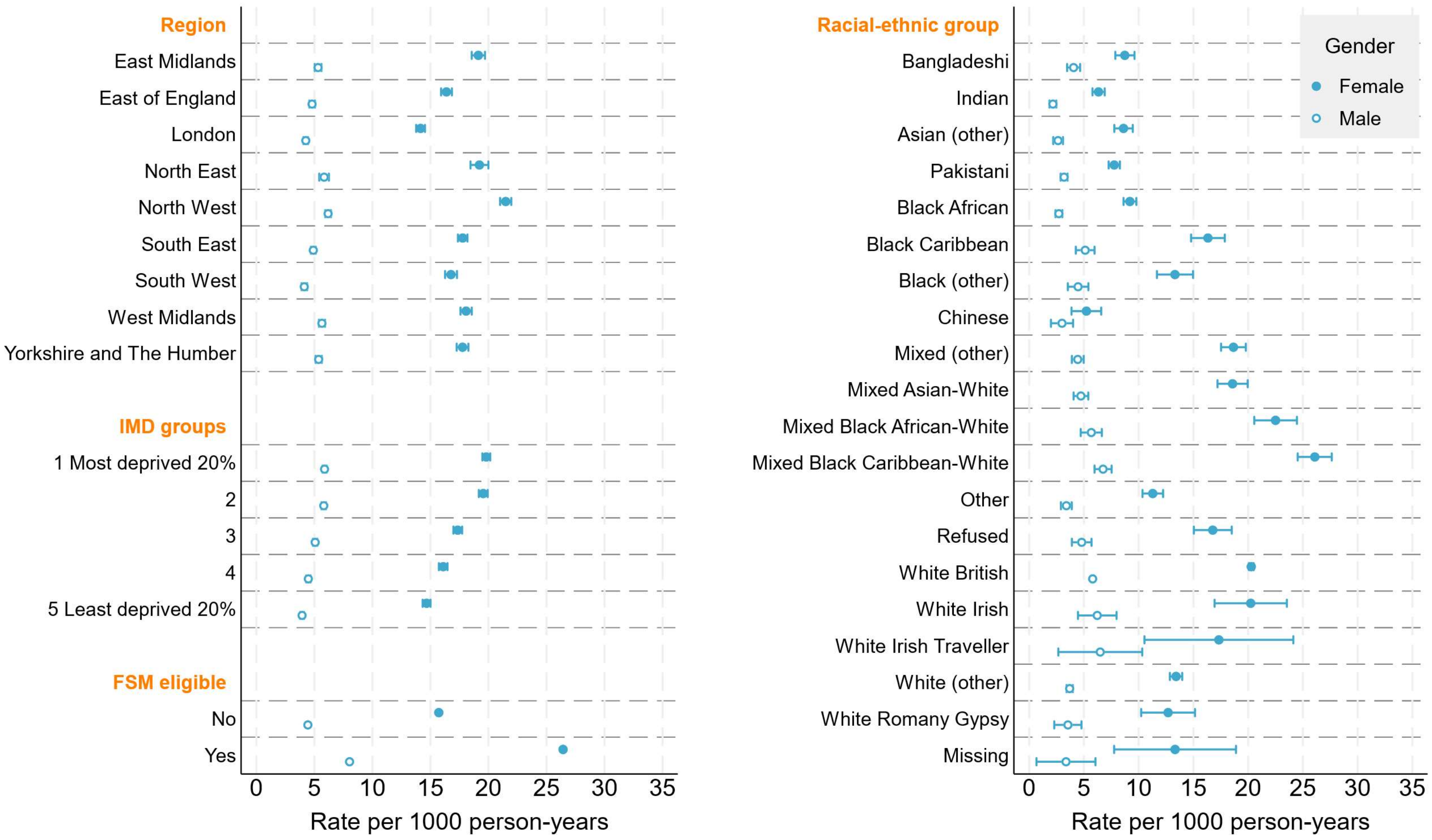

Region
East Midlands
East of England
London
North East
North West
South East
South West
West Midlands
Yorkshire and The Humber
IMD groups
1 Most deprived 20%
2
3
4
5 Least deprived 20%
FSM eligible
No
Yes
0 5 10 15 20 25 30 35
Rate per 1000 person-years
Racial-ethnic group
Bangladeshi
Indian
Asian (other)
Pakistani
Black African
Black Caribbean
Black (other)
Chinese
Mixed (other)
Mixed Asian-White
Mixed Black African-White
Mixed Black Caribbean-White
Other
Refused
White British
White Irish
White Irish Traveller
White (other)
White Romany Gypsy
Missing
Gender
Female
Male
0 5 10 15 20 25 30 35
Rate per 1000 person-years

**Figure S8.** Year-adjusted rates of first mental health-related emergency department attendance per 1000 person-years: derived from seperate Poisson regression regression models for each group of social strata and gender (11<16 years age group). FSM = free school meals; IMD = index of multiple deprivation

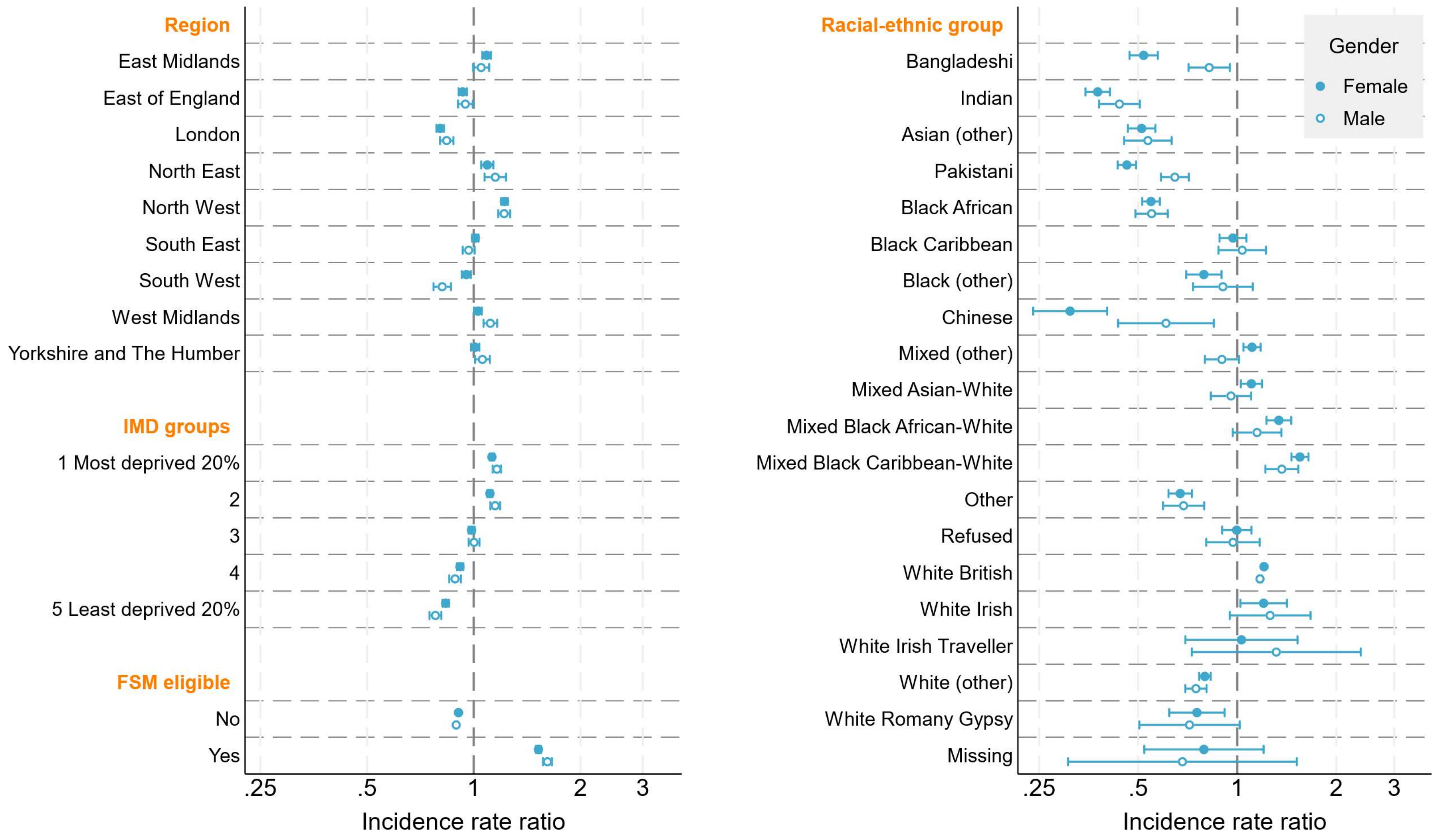

Region
East Midlands
East of England
London
North East
North West
South East
South West
West Midlands
Yorkshire and The Humber
IMD groups
1 Most deprived 20%
2
3
4
5 Least deprived 20%
FSM eligible
No
Yes
.25
.5
1
2
3
Incidence rate ratio
Racial-ethnic group
Bangladeshi
Indian
Asian (other)
Pakistani
Black African
Black Caribbean
Black (other)
Chinese
Mixed (other)
Mixed Asian-White
Mixed Black African-White
Mixed Black Caribbean-White
Other
Refused
White British
White Irish
White Irish Traveller
White (other)
White Romany Gypsy
Missing
Gender
Female
Male
.25
.5
1
2
3
Incidence rate ratio

**Figure S9.** Year-adjusted incidence rate ratios of referrals to mental health services (compared to the gender- and social strata-specific mean rate): derived from seperate Poisson regression regression models for each group of social strata and gender (11<16 years group). FSM = free school meals; IMD = index of multiple deprivation

Mental health-related emergency hospital admissions

**Table S12.** Year-adjusted rates of first mental health-related emergency hospital admission (95% confidence intervals): derived from age group-specific Poisson regression models with separate models for each group of social strata

| | | Age group | | |
|---|---|---|---|---|
| | | **11-<16** | **16-<18** | **18-<21** |
| | *N* people | 5,628,357 | 2,769,042 | 1,710,950 |
| | Overall | 3.8 (3.8, 3.9) | 4.9 (4.8, 5.0) | 3.9 (3.8, 3.9) |
| Gender | Male | 1.4 (1.4, 1.4) | 2.7 (2.6, 2.8) | 2.7 (2.6, 2.8) |
| | Female | 6.4 (6.3, 6.4) | 7.3 (7.2, 7.4) | 5.2 (5.0, 5.3) |
| Region | East Midlands | 3.3 (3.2, 3.4) | 4.6 (4.4, 4.8) | 4.2 (4.0, 4.4) |
| | East of England | 3.3 (3.2, 3.3) | 3.9 (3.7, 4.1) | 3.5 (3.3, 3.7) |
| | London | 2.4 (2.4, 2.5) | 3.4 (3.3, 3.5) | 2.7 (2.6, 2.9) |
| | North East | 4.6 (4.5, 4.8) | 5.9 (5.6, 6.2) | 5.1 (4.8, 5.5) |
| | North West | 5.2 (5.1, 5.3) | 4.7 (4.5, 4.8) | 4.4 (4.2, 4.5) |
| | South East | 3.8 (3.7, 3.8) | 6.3 (6.1, 6.5) | 4.5 (4.3, 4.7) |
| | South West | 5.0 (4.9, 5.1) | 7.6 (7.3, 7.8) | 4.6 (4.4, 4.8) |
| | West Midlands | 4.2 (4.1, 4.2) | 4.4 (4.2, 4.6) | 3.4 (3.2, 3.6) |
| | Yorkshire and The Humber | 3.4 (3.3, 3.4) | 4.2 (4.0, 4.4) | 3.3 (3.1, 3.4) |
| IMD groups | 1.Most deprived | 4.2 (4.2, 4.3) | 5.2 (5.0, 5.3) | 4.4 (4.2, 4.5) |
| | 2 | 4.1 (4.0, 4.1) | 5.2 (5.0, 5.3) | 4.2 (4.1, 4.4) |
| | 3 | 3.9 (3.8, 3.9) | 5.1 (4.9, 5.2) | 3.7 (3.5, 3.8) |
| | 4 | 3.6 (3.5, 3.6) | 4.6 (4.5, 4.7) | 3.6 (3.4, 3.7) |

| | | | | |
|---|---|---|---|---|
| | 5.Least deprived | 3.2 (3.2, 3.3) | 4.4 (4.3, 4.5) | 3.4 (3.2, 3.5) |
| FSM eligible | No | 3.3 (3.3, 3.4) | 4.4 (4.3, 4.4) | 3.5 (3.4, 3.5) |
| | Yes | 6.3 (6.2, 6.4) | 7.7 (7.5, 7.9) | 5.9 (5.7, 6.0) |
| Racial-ethnic group (minor) | Bangladeshi | 2.0 (1.8, 2.1) | 2.5 (2.1, 2.8) | 2.5 (2.1, 2.9) |
| | Indian | 1.2 (1.1, 1.3) | 1.9 (1.7, 2.2) | 1.7 (1.4, 1.9) |
| | Asian (other) | 1.5 (1.4, 1.7) | 2.6 (2.2, 2.9) | 2.7 (2.3, 3.2) |
| | Pakistani | 1.5 (1.5, 1.6) | 2.3 (2.1, 2.5) | 2.1 (1.9, 2.4) |
| | Black African | 1.4 (1.3, 1.5) | 2.4 (2.1, 2.6) | 2.6 (2.3, 2.9) |
| | Black Caribbean | 2.9 (2.7, 3.1) | 3.2 (2.8, 3.7) | 3.2 (2.7, 3.7) |
| | Black (other) | 2.3 (2.0, 2.5) | 3.4 (2.7, 4.0) | 3.9 (3.1, 4.7) |
| | Chinese | 1.3 (1.1, 1.6) | 2.3 (1.6, 3.0) | 1.9 (1.1, 2.6) |
| | Mixed (other) | 4.0 (3.8, 4.2) | 5.4 (4.9, 5.9) | 4.2 (3.6, 4.7) |
| | Mixed Asian & White | 4.1 (3.9, 4.4) | 5.5 (4.9, 6.1) | 4.5 (3.9, 5.2) |
| | Mixed Black African & White | 4.2 (3.9, 4.6) | 5.4 (4.6, 6.2) | 4.4 (3.4, 5.3) |
| | Mixed Black Caribbean & White | 5.3 (5.1, 5.6) | 6.6 (6.0, 7.2) | 4.7 (4.1, 5.3) |
| | Other | 2.0 (1.9, 2.2) | 2.5 (2.2, 2.9) | 2.8 (2.4, 3.3) |
| | Refused | 3.8 (3.5, 4.1) | 5.6 (4.7, 6.5) | 3.7 (2.8, 4.5) |
| | White British | 4.4 (4.4, 4.5) | 5.5 (5.4, 5.6) | 4.2 (4.2, 4.3) |
| | White Irish | 4.1 (3.6, 4.6) | 5.6 (4.4, 6.7) | 3.5 (2.4, 4.5) |
| | White Irish Traveller | 5.0 (3.7, 6.2) | 4.7 (2.2, 7.2) | 7.4 (3.8, 11.0) |
| | White (other) | 2.7 (2.6, 2.8) | 3.5 (3.2, 3.7) | 2.7 (2.4, 2.9) |
| | White Romany Gypsy | 2.5 (2.1, 2.9) | 5.0 (3.9, 6.1) | 3.2 (2.1, 4.3) |
| | Missing | 2.7 (1.8, 3.6) | 5.5 (3.0, 8.0) | 2.6 (0.7, 4.5) |

FSM = free school meals; IMD = index of multiple deprivation

**Table S13.** Year-adjusted incidence rate ratios of first mental health-related emergency hospital admission (95% confidence intervals), compared to the age- and social strata-specific mean rate: derived from age group-specific Poisson regression models with separate models for each group of social strata

| | | Age group | | |
|---|---|---|---|---|
| | | 11-<16 | 16-<18 | 18-<21 |
| | *N* people | 5,628,357 | 2,769,042 | 1,710,950 |
| Gender | Male | 0.48 (0.47, 0.48) | 0.62 (0.61, 0.63) | 0.74 (0.73, 0.75) |
| | Female | 2.18 (2.16, 2.20) | 1.68 (1.66, 1.71) | 1.40 (1.37, 1.42) |
| Region | East Midlands | 0.89 (0.87, 0.91) | 0.96 (0.93, 1.01) | 1.11 (1.05, 1.17) |
| | East of England | 0.88 (0.86, 0.89) | 0.82 (0.79, 0.85) | 0.92 (0.87, 0.96) |
| | London | 0.66 (0.64, 0.67) | 0.72 (0.69, 0.74) | 0.72 (0.69, 0.75) |
| | North East | 1.24 (1.21, 1.28) | 1.23 (1.17, 1.30) | 1.35 (1.26, 1.43) |
| | North West | 1.39 (1.37, 1.41) | 0.98 (0.95, 1.01) | 1.14 (1.10, 1.19) |
| | South East | 1.01 (1.00, 1.03) | 1.33 (1.29, 1.36) | 1.18 (1.14, 1.22) |
| | South West | 1.33 (1.31, 1.36) | 1.59 (1.54, 1.64) | 1.21 (1.16, 1.27) |
| | West Midlands | 1.12 (1.10, 1.14) | 0.93 (0.89, 0.96) | 0.89 (0.85, 0.94) |
| | Yorkshire and The Humber | 0.90 (0.89, 0.92) | 0.88 (0.84, 0.91) | 0.85 (0.81, 0.90) |
| IMD groups | 1.Most deprived | 1.11 (1.10, 1.13) | 1.06 (1.03, 1.08) | 1.13 (1.10, 1.16) |
| | 2 | 1.07 (1.06, 1.09) | 1.05 (1.03, 1.08) | 1.10 (1.07, 1.14) |
| | 3 | 1.02 (1.00, 1.03) | 1.04 (1.01, 1.07) | 0.96 (0.92, 0.99) |
| | 4 | 0.93 (0.92, 0.95) | 0.94 (0.91, 0.97) | 0.93 (0.89, 0.96) |
| | 5.Least deprived | 0.84 (0.83, 0.86) | 0.90 (0.88, 0.93) | 0.87 (0.84, 0.90) |
| FSM eligible | No | 0.90 (0.89, 0.90) | 0.91 (0.91, 0.92) | 0.92 (0.91, 0.92) |
| | Yes | 1.69 (1.67, 1.71) | 1.61 (1.57, 1.64) | 1.54 (1.50, 1.59) |
| Racial-ethnic group (minor) | Bangladeshi | 0.55 (0.51, 0.59) | 0.52 (0.46, 0.60) | 0.66 (0.56, 0.77) |
| | Indian | 0.34 (0.32, 0.37) | 0.41 (0.37, 0.46) | 0.44 (0.38, 0.51) |
| | Asian (other) | 0.43 (0.39, 0.46) | 0.55 (0.48, 0.63) | 0.72 (0.62, 0.84) |
| | Pakistani | 0.43 (0.41, 0.45) | 0.49 (0.45, 0.53) | 0.56 (0.51, 0.63) |
| | Black African | 0.40 (0.38, 0.42) | 0.50 (0.46, 0.55) | 0.68 (0.61, 0.76) |
| | Black Caribbean | 0.81 (0.75, 0.88) | 0.68 (0.60, 0.79) | 0.85 (0.72, 0.99) |
| | Black (other) | 0.63 (0.57, 0.71) | 0.72 (0.60, 0.87) | 1.03 (0.83, 1.28) |

| | | | | |
|---|---|---|---|---|
| | Chinese | 0.37 (0.31, 0.45) | 0.49 (0.36, 0.67) | 0.50 (0.33, 0.74) |
| | Mixed (other) | 1.11 (1.05, 1.16) | 1.15 (1.05, 1.26) | 1.11 (0.97, 1.25) |
| | Mixed Asian & White | 1.15 (1.08, 1.21) | 1.16 (1.04, 1.30) | 1.20 (1.03, 1.39) |
| | Mixed Black African & White | 1.18 (1.09, 1.27) | 1.16 (0.99, 1.35) | 1.16 (0.94, 1.43) |
| | Mixed Black Caribbean & White | 1.48 (1.41, 1.55) | 1.40 (1.28, 1.53) | 1.23 (1.09, 1.40) |
| | Other | 0.56 (0.52, 0.60) | 0.54 (0.47, 0.62) | 0.75 (0.64, 0.88) |
| | Refused | 1.06 (0.97, 1.15) | 1.20 (1.02, 1.40) | 0.97 (0.76, 1.23) |
| | White British | 1.23 (1.22, 1.24) | 1.18 (1.17, 1.19) | 1.12 (1.11, 1.14) |
| | White Irish | 1.13 (1.00, 1.28) | 1.18 (0.96, 1.46) | 0.91 (0.67, 1.24) |
| | White Irish Traveller | 1.38 (1.07, 1.78) | 1.01 (0.60, 1.70) | 1.95 (1.20, 3.18) |
| | White (other) | 0.75 (0.73, 0.78) | 0.74 (0.69, 0.79) | 0.71 (0.65, 0.78) |
| | White Romany Gypsy | 0.70 (0.60, 0.82) | 1.06 (0.85, 1.33) | 0.85 (0.60, 1.20) |
| | Missing | 0.76 (0.55, 1.05) | 1.17 (0.75, 1.84) | 0.69 (0.33, 1.44) |

FSM = free school meals; IMD = index of multiple deprivation

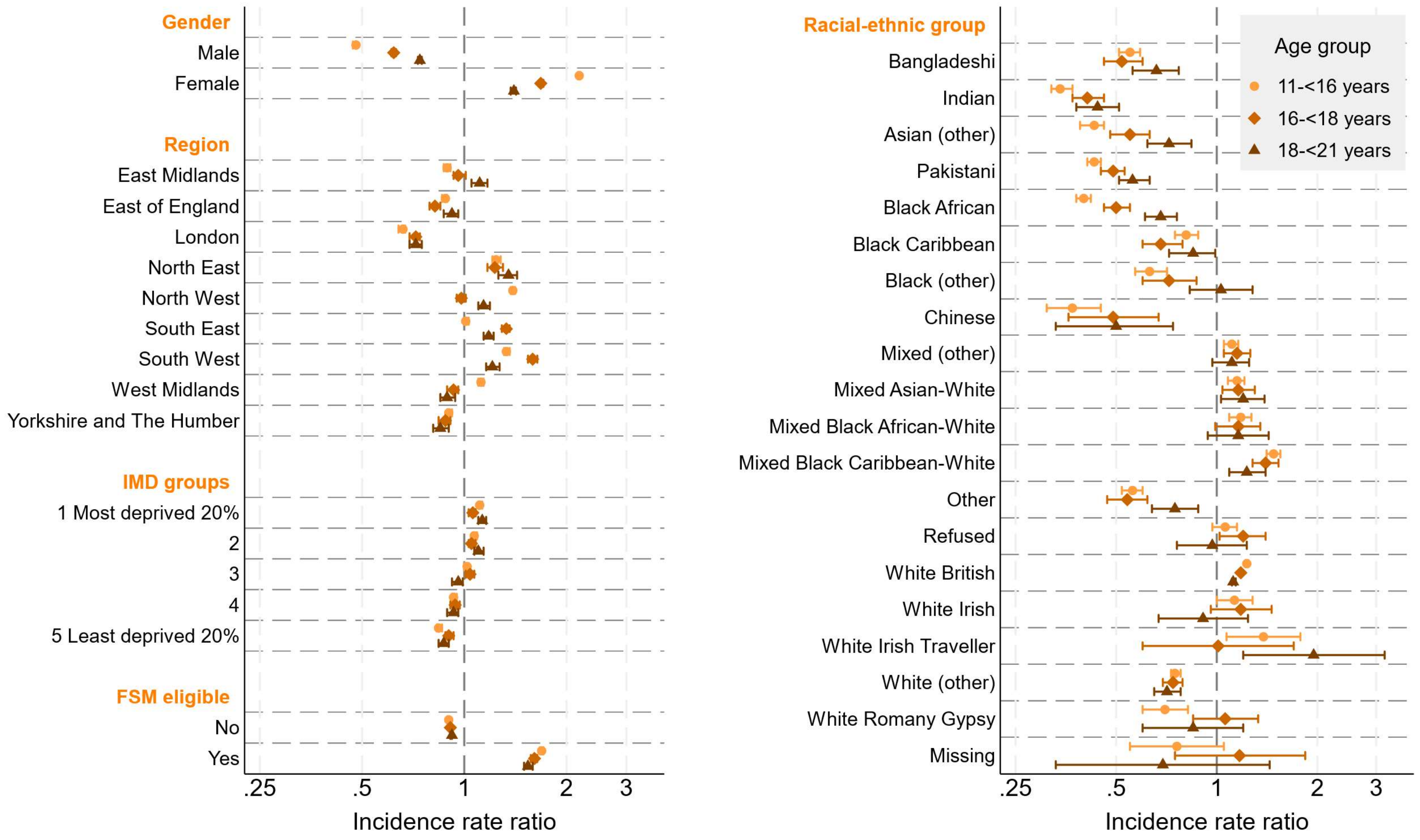


**Figure S10.** Year-adjusted incidence rate ratios (compared to the age- and social strata-specific mean rate) of first mental health-related emergency hospital admission with 95% confidence intervals: derived from age group-specific Poisson regression models with separate models for each group of social strata. FSM = free school meals; IMD = index of multiple deprivation

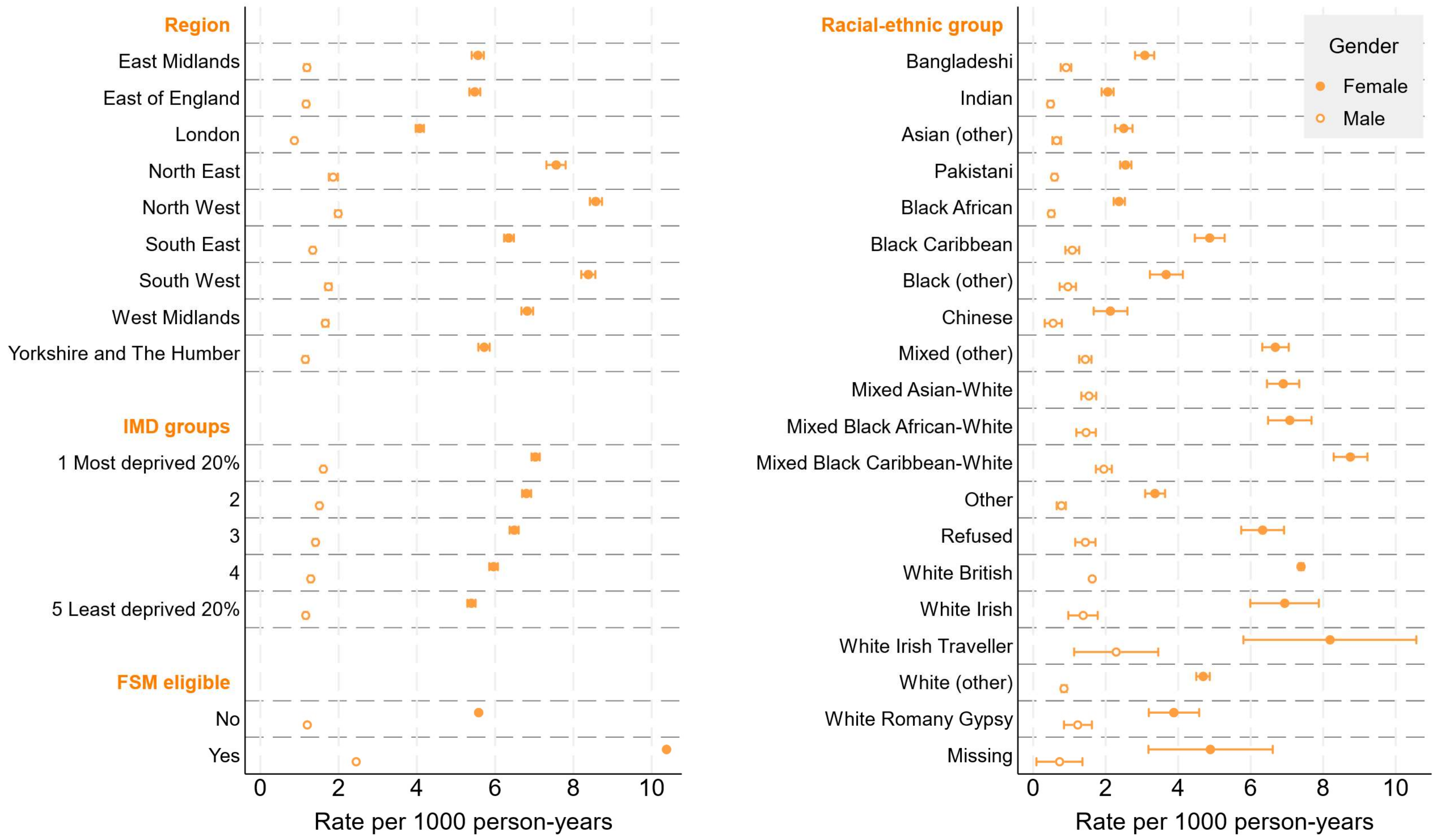

Region
East Midlands
East of England
London
North East
North West
South East
South West
West Midlands
Yorkshire and The Humber
IMD groups
1 Most deprived 20%
2
3
4
5 Least deprived 20%
FSM eligible
No
Yes
0
2
4
6
8
10
Rate per 1000 person-years
Racial-ethnic group
Bangladeshi
Indian
Asian (other)
Pakistani
Black African
Black Caribbean
Black (other)
Chinese
Mixed (other)
Mixed Asian-White
Mixed Black African-White
Mixed Black Caribbean-White
Other
Refused
White British
White Irish
White Irish Traveller
White (other)
White Romany Gypsy
Missing
0
2
4
6
8
10
Rate per 1000 person-years
Gender
Female
Male

**Figure S11.** Year-adjusted rates of first mental health-related emergency hospital admission per 1000 person-years: derived from seperate Poisson regression regression models for each group of social strata and gender (11<16 years group). FSM = free school meals; IMD = index of multiple deprivation

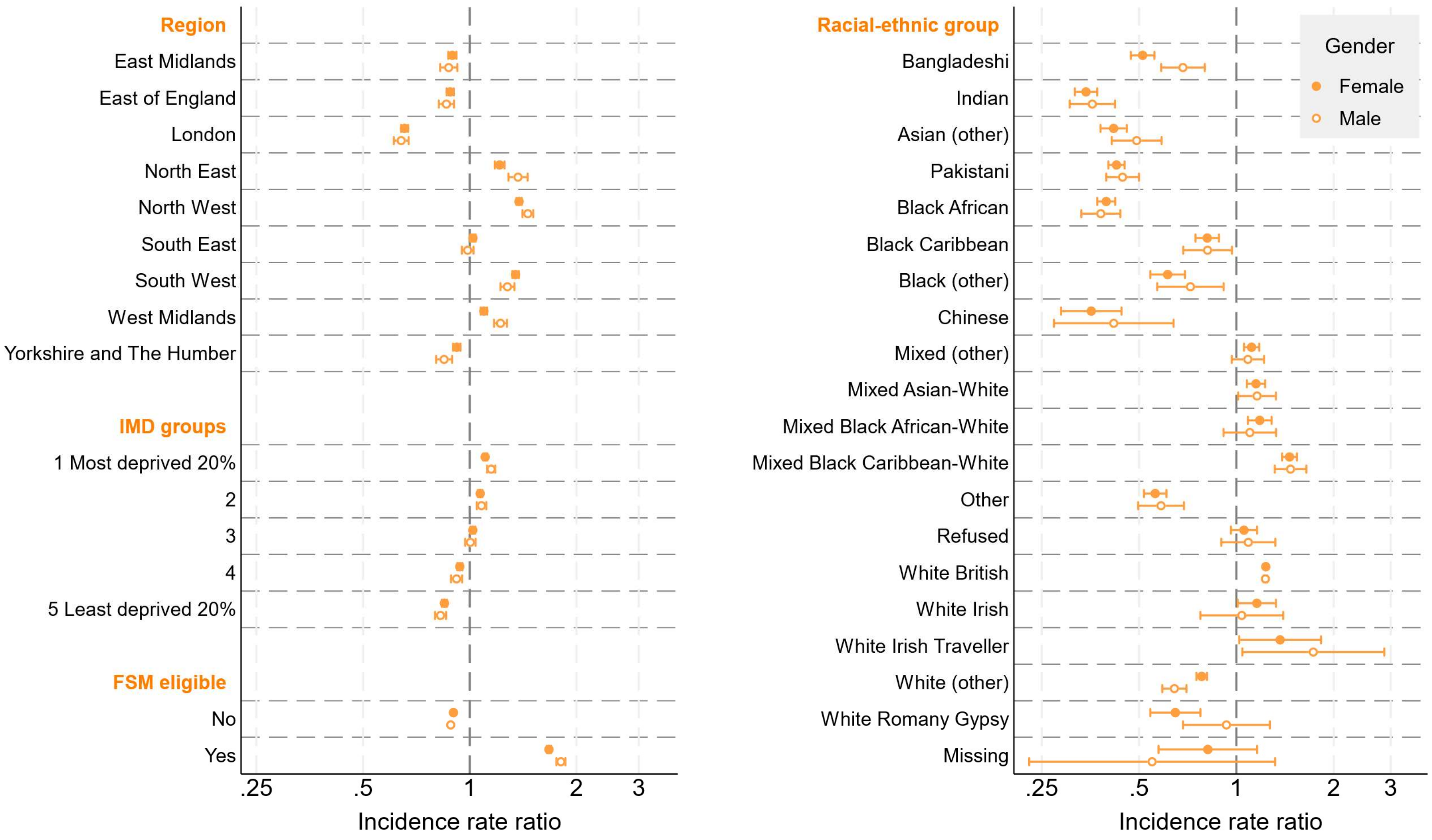

Region
East Midlands
East of England
London
North East
North West
South East
South West
West Midlands
Yorkshire and The Humber
IMD groups
1 Most deprived 20%
2
3
4
5 Least deprived 20%
FSM eligible
No
Yes
.25
.5
1
2
3
Incidence rate ratio
Racial-ethnic group
Bangladeshi
Indian
Asian (other)
Pakistani
Black African
Black Caribbean
Black (other)
Chinese
Mixed (other)
Mixed Asian-White
Mixed Black African-White
Mixed Black Caribbean-White
Other
Refused
White British
White Irish
White Irish Traveller
White (other)
White Romany Gypsy
Missing
Gender
Female
Male

**Figure S12.** Year-adjusted incidence rate ratios of first mental health-related emergency hospital admission (compared to the gender- and social strata-specific mean rate): derived from seperate Poisson regression regression models for each group of social strata and gender (11-<16 years group). FSM = free school meals; IMD = index of multiple deprivation

Mental health-related emergency hospital admissions (including potentially psychosomatic symptoms)

**Table S14.** Year-adjusted rates of first mental health-related emergency hospital admission (including potentially psychosomatic symptoms; 95% confidence intervals): derived from age group-specific Poisson regression models with separate models for each group of social strata

| | | Age group | | |
|---|---|---|---|---|
| | | **11-<16** | **16-<18** | **18-<21** |
| | *N* people | 5,628,357 | 2,769,042 | 1,710,950 |
| | Overall | 9.5 (9.5, 9.6) | 10.6 (10.5, 10.7) | 11.2 (11.1, 11.3) |
| Gender | Male | 6.3 (6.3, 6.4) | 6.6 (6.5, 6.7) | 7.5 (7.4, 7.7) |
| | Female | 12.9 (12.9, 13.0) | 15.0 (14.8, 15.1) | 15.4 (15.2, 15.5) |
| Region | East Midlands | 8.1 (7.9, 8.2) | 9.5 (9.3, 9.8) | 11.6 (11.2, 12.0) |
| | East of England | 8.7 (8.6, 8.8) | 9.7 (9.5, 10.0) | 10.5 (10.1, 10.8) |
| | London | 7.1 (7.0, 7.2) | 8.0 (7.8, 8.2) | 7.9 (7.6, 8.1) |
| | North East | 11.6 (11.4, 11.8) | 12.6 (12.1, 13.0) | 15.1 (14.5, 15.6) |
| | North West | 12.2 (12.1, 12.3) | 11.3 (11.1, 11.6) | 12.9 (12.6, 13.3) |
| | South East | 8.9 (8.8, 9.0) | 11.7 (11.4, 11.9) | 11.9 (11.6, 12.2) |
| | South West | 11.1 (11.0, 11.3) | 13.6 (13.2, 13.9) | 11.9 (11.6, 12.3) |
| | West Midlands | 10.9 (10.7, 11.0) | 10.8 (10.5, 11.0) | 12.4 (12.0, 12.7) |
| | Yorkshire and The Humber | 8.8 (8.7, 8.9) | 9.6 (9.4, 9.9) | 9.5 (9.2, 9.8) |
| IMD groups | 1.Most deprived | 10.5 (10.4, 10.6) | 11.6 (11.4, 11.8) | 12.8 (12.6, 13.1) |
| | 2 | 10.0 (9.9, 10.1) | 11.1 (10.9, 11.3) | 11.8 (11.5, 12.0) |
| | 3 | 9.7 (9.6, 9.8) | 10.8 (10.6, 11.0) | 10.9 (10.6, 11.1) |
| | 4 | 9.0 (8.9, 9.1) | 9.8 (9.6, 10.0) | 10.3 (10.1, 10.6) |
| | 5.Least deprived | 8.2 (8.1, 8.3) | 9.3 (9.1, 9.5) | 9.8 (9.6, 10.1) |
| FSM eligible | No | 8.8 (8.7, 8.8) | 9.7 (9.6, 9.8) | 10.4 (10.3, 10.5) |
| | Yes | 13.4 (13.3, 13.5) | 15.1 (14.9, 15.4) | 15.3 (15.0, 15.6) |
| Racial-ethnic group (minor) | Bangladeshi | 7.1 (6.8, 7.4) | 7.3 (6.8, 7.9) | 7.1 (6.4, 7.7) |
| | Indian | 5.9 (5.7, 6.1) | 6.3 (5.9, 6.8) | 6.5 (6.0, 7.0) |

| | | | | |
|---|---|---|---|---|
| | Asian (other) | 5.7 (5.5, 6.0) | 6.9 (6.3, 7.4) | 7.5 (6.8, 8.2) |
| | Pakistani | 8.6 (8.4, 8.8) | 8.6 (8.2, 9.0) | 9.7 (9.2, 10.2) |
| | Black African | 5.8 (5.6, 5.9) | 6.6 (6.2, 7.0) | 8.2 (7.7, 8.7) |
| | Black Caribbean | 7.7 (7.4, 8.1) | 7.8 (7.1, 8.5) | 8.3 (7.5, 9.2) |
| | Black (other) | 6.9 (6.5, 7.4) | 7.6 (6.6, 8.6) | 10.0 (8.7, 11.4) |
| | Chinese | 3.7 (3.3, 4.1) | 4.9 (3.8, 5.9) | 4.6 (3.4, 5.8) |
| | Mixed (other) | 9.0 (8.7, 9.3) | 10.0 (9.3, 10.6) | 10.5 (9.6, 11.3) |
| | Mixed Asian & White | 9.2 (8.8, 9.6) | 10.4 (9.5, 11.2) | 10.5 (9.5, 11.5) |
| | Mixed Black African & White | 9.3 (8.8, 9.8) | 9.8 (8.7, 10.9) | 11.1 (9.6, 12.5) |
| | Mixed Black Caribbean & White | 11.0 (10.6, 11.4) | 12.4 (11.6, 13.2) | 11.7 (10.7, 12.6) |
| | Other | 6.8 (6.5, 7.0) | 7.5 (6.9, 8.1) | 8.5 (7.7, 9.3) |
| | Refused | 8.8 (8.3, 9.3) | 10.3 (9.1, 11.5) | 9.5 (8.1, 10.9) |
| | White British | 10.4 (10.3, 10.4) | 11.5 (11.4, 11.7) | 12.1 (12.0, 12.2) |
| | White Irish | 9.5 (8.7, 10.3) | 10.4 (8.8, 12.0) | 10.5 (8.6, 12.4) |
| | White Irish Traveller | 12.6 (10.6, 14.7) | 10.2 (6.5, 13.9) | 18.1 (12.2, 23.9) |
| | White (other) | 7.4 (7.2, 7.6) | 7.8 (7.4, 8.2) | 8.7 (8.3, 9.2) |
| | White Romany Gypsy | 8.4 (7.7, 9.2) | 11.6 (9.9, 13.4) | 12.4 (10.2, 14.7) |
| | Missing | 7.0 (5.5, 8.4) | 10.1 (6.7, 13.5) | 7.8 (4.4, 11.2) |

FSM = free school meals; IMD = index of multiple deprivation

**Table S15.** Year-adjusted incidence rate ratios of first mental health-related emergency hospital admission including potentially psychosomatic symptoms (95% confidence intervals), compared to the age- and social strata-specific mean rate: derived from age group-specific Poisson regression models with separate models for each group of social strata

| | | Age group | |
|---|---|---|---|
| | **11-<16** | **16-<18** | **18-<21** |
| *N* people | 5,628,357 | 2,769,042 | 1,710,950 |

| | | | | |
|---|---|---|---|---|
| Gender | Male | 0.71 (0.70, 0.71) | 0.68 (0.67, 0.68) | 0.72 (0.71, 0.72) |
| | Female | 1.44 (1.44, 1.45) | 1.54 (1.52, 1.55) | 1.46 (1.44, 1.47) |
| Region | East Midlands | 0.86 (0.85, 0.87) | 0.91 (0.89, 0.94) | 1.05 (1.02, 1.08) |
| | East of England | 0.92 (0.91, 0.94) | 0.93 (0.91, 0.95) | 0.95 (0.92, 0.97) |
| | London | 0.75 (0.75, 0.76) | 0.76 (0.74, 0.78) | 0.71 (0.69, 0.73) |
| | North East | 1.24 (1.22, 1.26) | 1.20 (1.16, 1.24) | 1.36 (1.31, 1.42) |
| | North West | 1.30 (1.29, 1.31) | 1.08 (1.06, 1.11) | 1.17 (1.14, 1.20) |
| | South East | 0.95 (0.94, 0.96) | 1.12 (1.10, 1.14) | 1.08 (1.05, 1.10) |
| | South West | 1.18 (1.17, 1.20) | 1.30 (1.27, 1.33) | 1.08 (1.05, 1.11) |
| | West Midlands | 1.16 (1.14, 1.17) | 1.03 (1.00, 1.05) | 1.12 (1.09, 1.15) |
| | Yorkshire and The Humber | 0.94 (0.93, 0.95) | 0.92 (0.89, 0.94) | 0.86 (0.83, 0.88) |
| IMD groups | 1.Most deprived | 1.11 (1.10, 1.11) | 1.10 (1.09, 1.12) | 1.15 (1.13, 1.17) |
| | 2 | 1.05 (1.04, 1.06) | 1.05 (1.04, 1.07) | 1.05 (1.03, 1.07) |
| | 3 | 1.02 (1.01, 1.03) | 1.02 (1.00, 1.04) | 0.97 (0.95, 0.99) |
| | 4 | 0.94 (0.93, 0.95) | 0.93 (0.91, 0.94) | 0.93 (0.91, 0.95) |
| | 5.Least deprived | 0.86 (0.85, 0.87) | 0.88 (0.87, 0.90) | 0.88 (0.86, 0.90) |
| FSM eligible | No | 0.93 (0.93, 0.93) | 0.93 (0.93, 0.93) | 0.94 (0.93, 0.94) |
| | Yes | 1.42 (1.41, 1.43) | 1.45 (1.43, 1.47) | 1.38 (1.35, 1.41) |
| Racial-ethnic group (minor) | Bangladeshi | 0.76 (0.73, 0.79) | 0.70 (0.65, 0.76) | 0.64 (0.58, 0.70) |
| | Indian | 0.62 (0.60, 0.65) | 0.61 (0.57, 0.65) | 0.59 (0.54, 0.64) |
| | Asian (other) | 0.61 (0.58, 0.64) | 0.66 (0.61, 0.71) | 0.68 (0.62, 0.75) |
| | Pakistani | 0.92 (0.90, 0.94) | 0.83 (0.79, 0.87) | 0.88 (0.84, 0.93) |
| | Black African | 0.61 (0.60, 0.63) | 0.63 (0.60, 0.67) | 0.74 (0.70, 0.79) |
| | Black Caribbean | 0.83 (0.79, 0.86) | 0.75 (0.69, 0.82) | 0.75 (0.68, 0.83) |
| | Black (other) | 0.74 (0.69, 0.79) | 0.73 (0.64, 0.83) | 0.91 (0.79, 1.04) |
| | Chinese | 0.40 (0.35, 0.44) | 0.47 (0.38, 0.58) | 0.42 (0.32, 0.54) |
| | Mixed (other) | 0.96 (0.93, 0.99) | 0.96 (0.89, 1.02) | 0.95 (0.87, 1.03) |
| | Mixed Asian & White | 0.98 (0.94, 1.02) | 1.00 (0.92, 1.08) | 0.95 (0.86, 1.05) |
| | Mixed Black African & White | 0.99 (0.94, 1.05) | 0.94 (0.84, 1.06) | 1.00 (0.87, 1.14) |

| | | | | |
|---|---|---|---|---|
| | Mixed Black Caribbean & White | 1.17 (1.13, 1.21) | 1.19 (1.12, 1.27) | 1.05 (0.97, 1.14) |
| | Other | 0.72 (0.69, 0.75) | 0.72 (0.66, 0.78) | 0.76 (0.70, 0.84) |
| | Refused | 0.94 (0.89, 1.00) | 0.99 (0.88, 1.11) | 0.86 (0.74, 1.00) |
| | White British | 1.11 (1.10, 1.11) | 1.11 (1.10, 1.11) | 1.09 (1.09, 1.10) |
| | White Irish | 1.02 (0.94, 1.10) | 1.00 (0.86, 1.16) | 0.95 (0.79, 1.13) |
| | White Irish Traveller | 1.35 (1.15, 1.58) | 0.98 (0.68, 1.40) | 1.63 (1.18, 2.25) |
| | White (other) | 0.79 (0.77, 0.81) | 0.75 (0.72, 0.78) | 0.79 (0.75, 0.83) |
| | White Romany Gypsy | 0.90 (0.83, 0.98) | 1.12 (0.96, 1.30) | 1.12 (0.94, 1.34) |
| | Missing | 0.74 (0.60, 0.91) | 0.97 (0.69, 1.36) | 0.70 (0.45, 1.09) |

FSM = free school meals; IMD = index of multiple deprivation

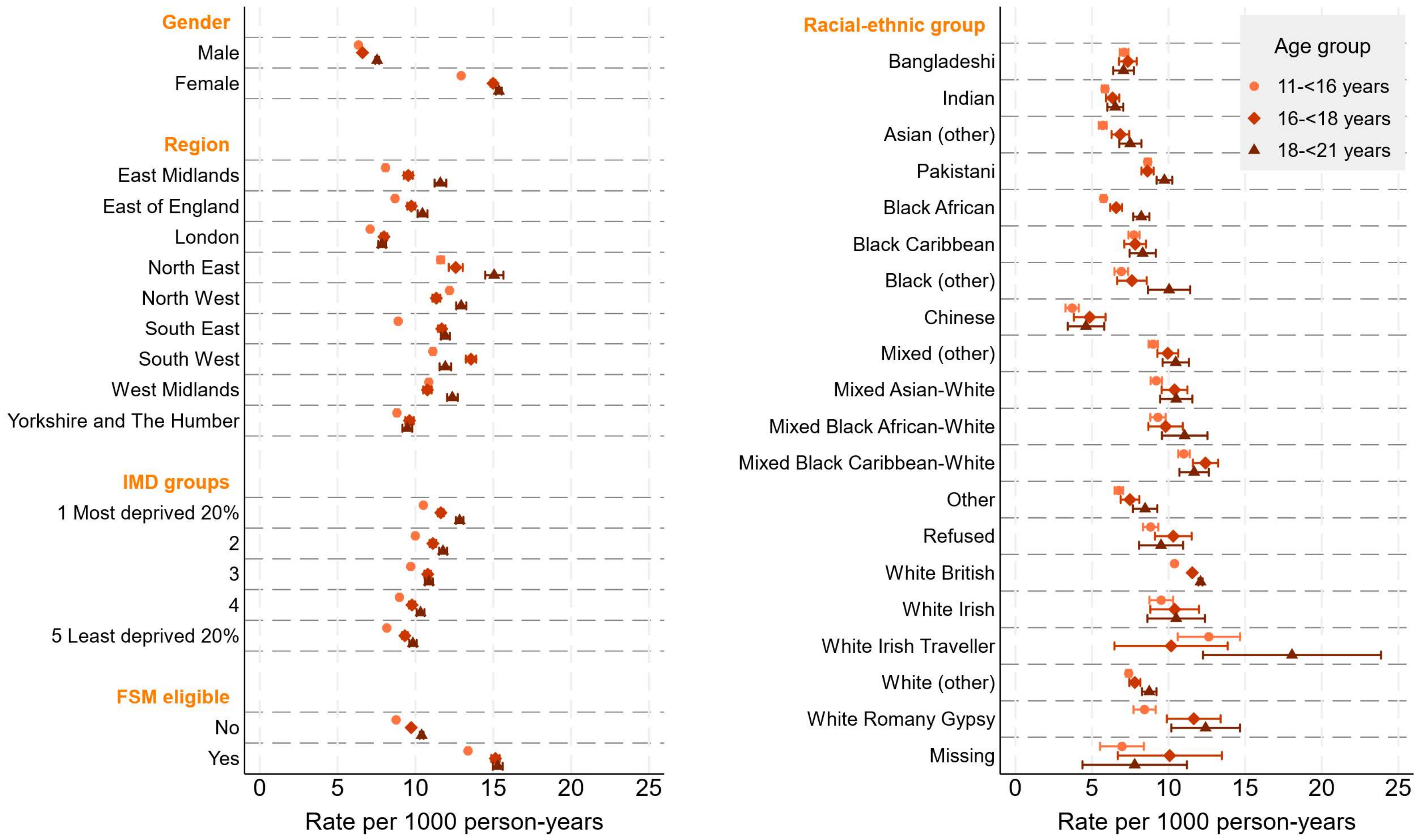


**Figure S13.** Year-adjusted rates of first mental health-related emergency hospital admission (including potentially psychosomatic symptoms; per 1000 person-years) with 95% confidence intervals: derived from age group-specific Poisson regression models with separate models for each group of social strata. FSM = free school meals; IMD = index of multiple deprivation

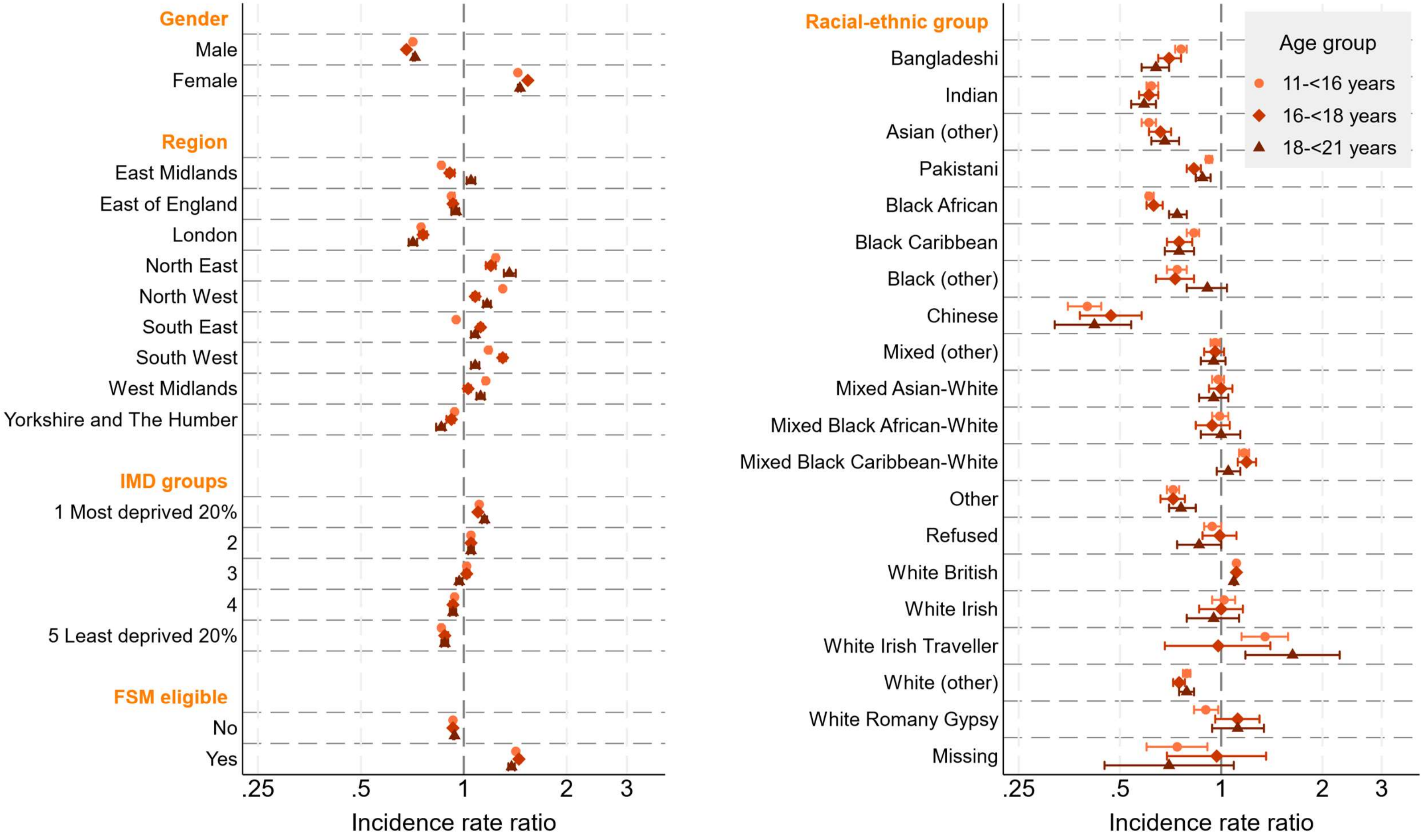
Gender
Male
Female
Region
East Midlands
East of England
London
North East
North West
South East
South West
West Midlands
Yorkshire and The Humber
IMD groups
1 Most deprived 20%
2
3
4
5 Least deprived 20%
FSM eligible
No
Yes
.25
.5
1
2
3
Incidence rate ratio
Racial-ethnic group
Bangladeshi
Indian
Asian (other)
Pakistani
Black African
Black Caribbean
Black (other)
Chinese
Mixed (other)
Mixed Asian-White
Mixed Black African-White
Mixed Black Caribbean-White
Other
Refused
White British
White Irish
White Irish Traveller
White (other)
White Romany Gypsy
Missing
.25
.5
1
2
3
Incidence rate ratio
Age group
11-<16 years
16-<18 years
18-<21 years

**Figure S14.** Year-adjusted incidence rate ratios (compared to the age- and social strata-specific mean rate) of first mental health-related emergency hospital admission including potentially psychosomatic symptoms with 95% confidence intervals: derived from age group-specific Poisson regression models with separate models for each group of social strata. FSM = free school meals; IMD = index of multiple deprivation

Bradford cohort results

In we repeated all main analyses on a sub-cohort of pupils residing in the Local Authority of Bradford (defined by the Middle Layer Super Output Area of their home address) in Year 7 between 2012/13 and 2021/22. For these analyses, the follow-up for all outcomes was restricted to the first chronological event of: outcome event, death, age **16 years** or end of study (31st August 2022). This means that the results pertain to adolescents aged 11-<16 years only. We restricted follow-up time compared to the national cohort due to low numbers of events in older ages groups for some events.

**Table S16.** Year-adjusted rates of first referral to NHS-funded mental health services, mental health-related ED attendance and emergency hospital admission per 1000 person-years (95% confidence intervals) among pupils residing in Bradford: derived from Poisson regression models with separate models for each group of social strata (11-<16 years old)

| | | **Referrals to MHS** | **ED attendances** | **Hospital admissions** | **Hospital admissions (+PS)** |
|---|---|---|---|---|---|
| | *N* people | 71,445 | 45,430 | 71,480 | 71,480 |
| | Overall | 40.0 (39.0, 40.9) | 6.8 (6.2, 7.4) | 3.4 (3.2, 3.6) | 11.0 (10.6, 11.4) |
| Gender | Male | 31.8 (30.7, 33.0) | 3.1 (2.5, 3.7) | 1.1 (0.9, 1.3) | 8.0 (7.6, 8.5) |
| | Female | 48.6 (47.2, 50.1) | 10.7 (9.6, 11.8) | 5.8 (5.4, 6.3) | 14.2 (13.5, 14.8) |
| IMD groups | 1. Most deprived | 38.1 (36.9, 39.3) | 5.4 (4.6, 6.1) | 3.0 (2.7, 3.3) | 10.6 (10.1, 11.1) |
| | 2 | 44.4 (42.0, 46.9) | 8.1 (6.5, 9.8) | 3.7 (3.1, 4.3) | 12.0 (11.0, 13.1) |
| | 3 | 42.1 (39.6, 44.6) | 7.7 (6.0, 9.5) | 3.8 (3.2, 4.5) | 11.7 (10.6, 12.8) |
| | 4 | 45.5 (41.9, 49.0) | 10.8 (7.9, 13.7) | 4.1 (3.2, 5.0) | 11.8 (10.3, 13.4) |
| | 5. Least deprived | 34.4 (31.1, 37.6) | 8.9 (6.2, 11.7) | 4.0 (3.1, 5.0) | 9.5 (8.0, 10.9) |
| FSM eligible | No | 34.6 (33.6, 35.5) | 6.3 (5.6, 7.0) | 3.0 (2.8, 3.3) | 10.3 (9.9, 10.8) |
| | Yes | 61.1 (58.5, 63.6) | 8.3 (6.9, 9.7) | 4.7 (4.1, 5.3) | 13.6 (12.6, 14.6) |
| Racial-ethnic group | Bangladeshi | 15.7 (12.5, 18.8) | 2.4 (0.3, 4.5) | 0.8 (0.2, 1.5) | 6.3 (4.6, 8.0) |
| | Indian | 16.5 (12.4, 20.6) | | 1.1 (0.2, 2.0) | 8.7 (6.2, 11.3) |
| | Asian (other) | 26.4 (19.3, 33.5) | | 1.3 (0.0, 2.6) | 7.7 (4.6, 10.8) |
| | Pakistani | 24.4 (23.2, 25.6) | 3.6 (2.9, 4.4) | 1.9 (1.6, 2.2) | 10.9 (10.2, 11.6) |
| | Black African | 17.0 (11.6, 22.4) | | 2.0 (0.4, 3.7) | 4.7 (2.1, 7.2) |

| | | | | |
|---|---|---|---|---|
| Black Caribbean | 41.7 (24.1, 59.3) | | | 10.6 (3.3, 17.8) |
| Black (other) | 18.1 (2.2, 34.0) | | | |
| Chinese | | | | |
| Mixed (other) | 35.0 (27.9, 42.1) | 6.1 (1.6, 10.6) | 2.2 (0.7, 3.7) | 9.4 (6.1, 12.6) |
| Mixed Asian & White | 66.4 (58.0, 74.8) | 14.8 (8.7, 20.9) | 4.3 (2.5, 6.0) | 11.3 (8.4, 14.2) |
| Mixed Black African & White | 42.9 (27.2, 58.5) | | | 11.5 (4.4, 18.7) |
| Mixed Black Caribbean & White | 59.8 (49.7, 69.9) | 12.7 (5.6, 19.8) | 7.6 (4.6, 10.5) | 13.6 (9.6, 17.6) |
| Other | 15.8 (10.8, 20.7) | | 1.6 (0.2, 2.9) | 5.9 (3.2, 8.6) |
| Refused | 36.6 (25.5, 47.7) | 6.8 (0.1, 13.5) | 4.0 (0.8, 7.2) | 11.0 (5.4, 16.5) |
| White British | 57.8 (56.2, 59.5) | 10.0 (8.8, 11.1) | 4.9 (4.5, 5.3) | 12.2 (11.5, 12.8) |
| White Irish | 65.3 (34.1, 96.5) | | | 14.7 (1.7, 27.7) |
| White Irish Traveller | | | | |
| White (other) | 24.1 (20.3, 27.9) | 5.0 (2.4, 7.6) | 3.6 (2.4, 4.8) | 7.8 (5.9, 9.7) |
| White Romany Gypsy | 19.4 (14.3, 24.6) | 4.4 (0.1, 8.8) | 2.4 (0.8, 3.9) | 9.6 (6.4, 12.7) |
| Missing | | | | |

ED = emergency department; FSM = free school meals; IMD = index of multiple deprivation; MHS = mental health services; PS = potentially psychosomatic symptoms; rates with confidence intervals crossing 0 are not shown due to unreliability of estimates

**Table S17.** Year-adjusted incidence rate ratios of first referral to NHS-funded mental health services, mental health-related ED attendance and emergency hospital admission (95% confidence intervals), compared to the social strata-specific mean rate, among pupils residing in Bradford: derived from Poisson regression models with separate models for each group of social strata (11-<16 years old)

| | | **Referrals to MHS** | **ED attendances** | **Hospital admissions** | **Hospital admissions (+PS)** |
|---|---|---|---|---|---|
| | *N* people | 71,445 | 45,430 | 71,480 | 71,480 |
| Gender | Male | 0.81 (0.80, 0.83) | 0.55 (0.49, 0.61) | 0.45 (0.41, 0.48) | 0.76 (0.73, 0.79) |
| | Female | 1.24 (1.21, 1.27) | 1.89 (1.69, 2.10) | 2.36 (2.16, 2.59) | 1.34 (1.29, 1.39) |
| IMD groups | 1. Most deprived | 0.96 (0.94, 0.98) | 0.82 (0.75, 0.88) | 0.90 (0.85, 0.95) | 0.97 (0.93, 1.00) |
| | 2 | 1.12 (1.06, 1.17) | 1.24 (1.02, 1.50) | 1.10 (0.95, 1.27) | 1.10 (1.01, 1.19) |
| | 3 | 1.06 (1.00, 1.12) | 1.18 (0.95, 1.46) | 1.14 (0.98, 1.33) | 1.06 (0.97, 1.16) |
| | 4 | 1.14 (1.06, 1.23) | 1.65 (1.27, 2.15) | 1.22 (0.99, 1.51) | 1.08 (0.95, 1.22) |
| | 5. Least deprived | 0.86 (0.79, 0.95) | 1.36 (1.00, 1.84) | 1.19 (0.95, 1.50) | 0.86 (0.74, 1.00) |
| FSM eligible | No | 0.89 (0.88, 0.90) | 0.94 (0.90, 0.98) | 0.91 (0.88, 0.94) | 0.94 (0.93, 0.96) |
| | Yes | 1.57 (1.51, 1.63) | 1.23 (1.05, 1.43) | 1.40 (1.25, 1.57) | 1.24 (1.16, 1.33) |
| Racial-ethnic group | Bangladeshi | 0.44 (0.36, 0.53) | 0.42 (0.18, 0.98) | 0.29 (0.14, 0.60) | 0.59 (0.45, 0.77) |
| | Indian | 0.46 (0.36, 0.59) | | 0.39 (0.18, 0.86) | 0.82 (0.62, 1.10) |
| | Asian (other) | 0.74 (0.56, 0.96) | | 0.46 (0.17, 1.21) | 0.73 (0.48, 1.09) |
| | Pakistani | 0.68 (0.65, 0.70) | 0.63 (0.54, 0.74) | 0.65 (0.58, 0.73) | 1.03 (0.98, 1.08) |
| | Black African | 0.47 (0.35, 0.65) | | 0.71 (0.32, 1.56) | 0.44 (0.26, 0.76) |
| | Black Caribbean | 1.16 (0.76, 1.77) | | | 1.00 (0.50, 1.97) |
| | Black (other) | 0.50 (0.21, 1.21) | | | |
| | Chinese | | | | |
| | Mixed (other) | 0.97 (0.80, 1.19) | 1.07 (0.51, 2.23) | 0.76 (0.38, 1.52) | 0.88 (0.63, 1.24) |
| | Mixed Asian & White | 1.85 (1.63, 2.10) | 2.59 (1.71, 3.92) | 1.47 (0.98, 2.21) | 1.06 (0.82, 1.37) |
| | Mixed Black African & White | 1.19 (0.83, 1.72) | | | 1.09 (0.59, 2.02) |
| | Mixed Black Caribbean & White | 1.67 (1.41, 1.97) | 2.21 (1.26, 3.90) | 2.62 (1.77, 3.88) | 1.28 (0.95, 1.72) |

| | | | | |
|---|---|---|---|---|
| Other | 0.44 (0.32, 0.60) | | 0.54 (0.23, 1.28) | 0.56 (0.35, 0.88) |
| Refused | 1.02 (0.75, 1.38) | 1.19 (0.45, 3.17) | 1.38 (0.62, 3.08) | 1.03 (0.63, 1.71) |
| White British | 1.61 (1.57, 1.66) | 1.74 (1.55, 1.95) | 1.68 (1.56, 1.82) | 1.15 (1.10, 1.20) |
| White Irish | 1.82 (1.13, 2.93) | | | 1.39 (0.57, 3.35) |
| White Irish Traveller | | | | |
| White (other) | 0.67 (0.58, 0.78) | 0.87 (0.52, 1.46) | 1.24 (0.88, 1.75) | 0.73 (0.58, 0.93) |
| White Romany Gypsy | 0.54 (0.42, 0.71) | 0.78 (0.29, 2.05) | 0.82 (0.43, 1.56) | 0.90 (0.65, 1.25) |
| Missing | | | | |

ED = emergency department; FSM = free school meals; IMD = index of multiple deprivation; MHS = mental health services; PS = potentially psychosomatic symptoms; IRRs based on estimated rates with confidence intervals less than 0 are not shown

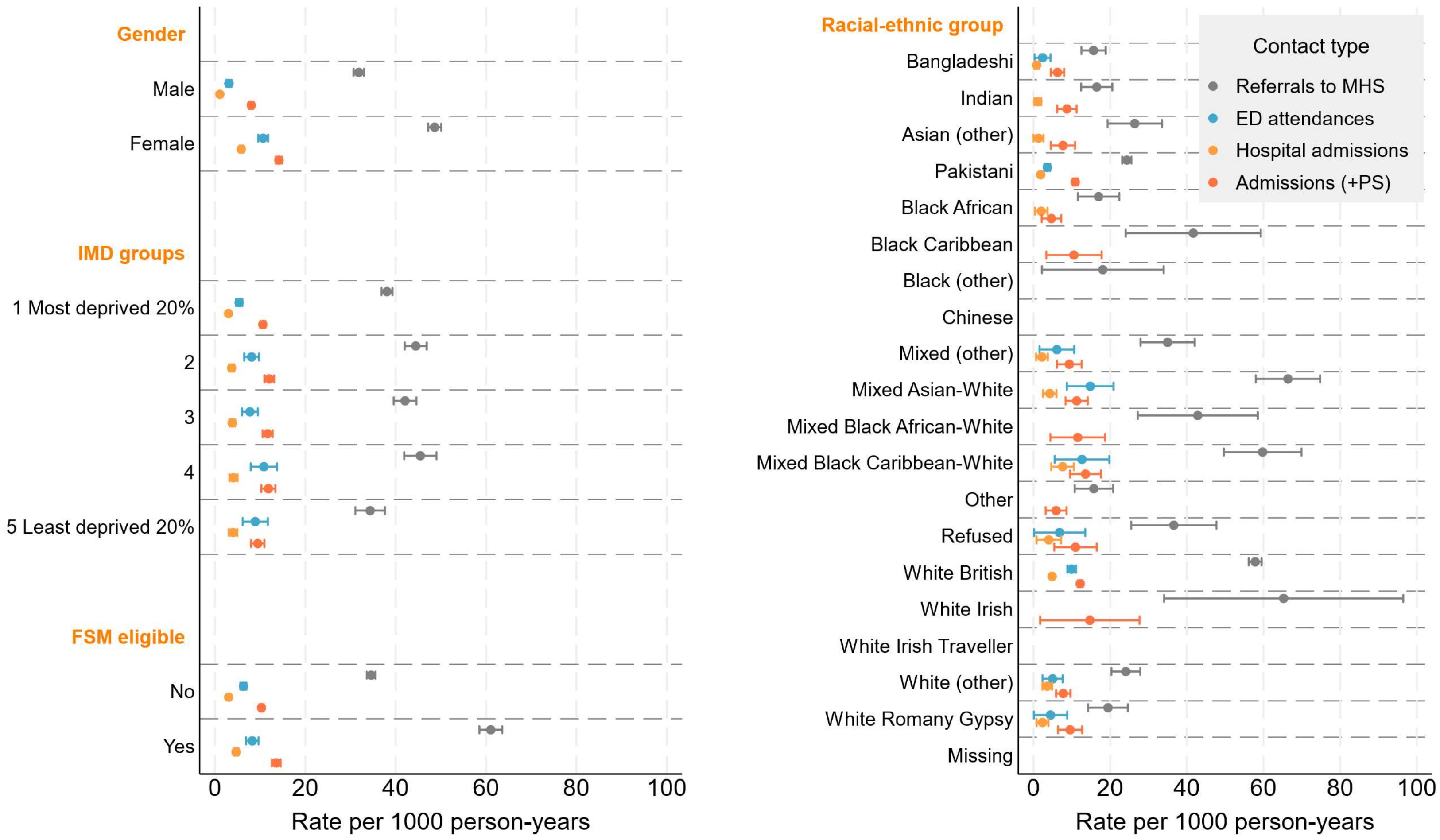


**Figure S16.** Year-adjusted incidence rate ratios (compared to the social strata- and service-specific mean rate) of first referrals to mental health services, mental health-related emergency department attendances and hospital admissions among pupils residing in Bradford: derived from service-specific Poisson regression models with separate models for each social strata (11-<16 years old group only). Estimated rates with confidence intervals less than 0 are not shown. ED = emergency department; FSM = free school meals; IMD = index of multiple deprivation; MHS = mental health services; PS = potentially psychosomatic symptoms

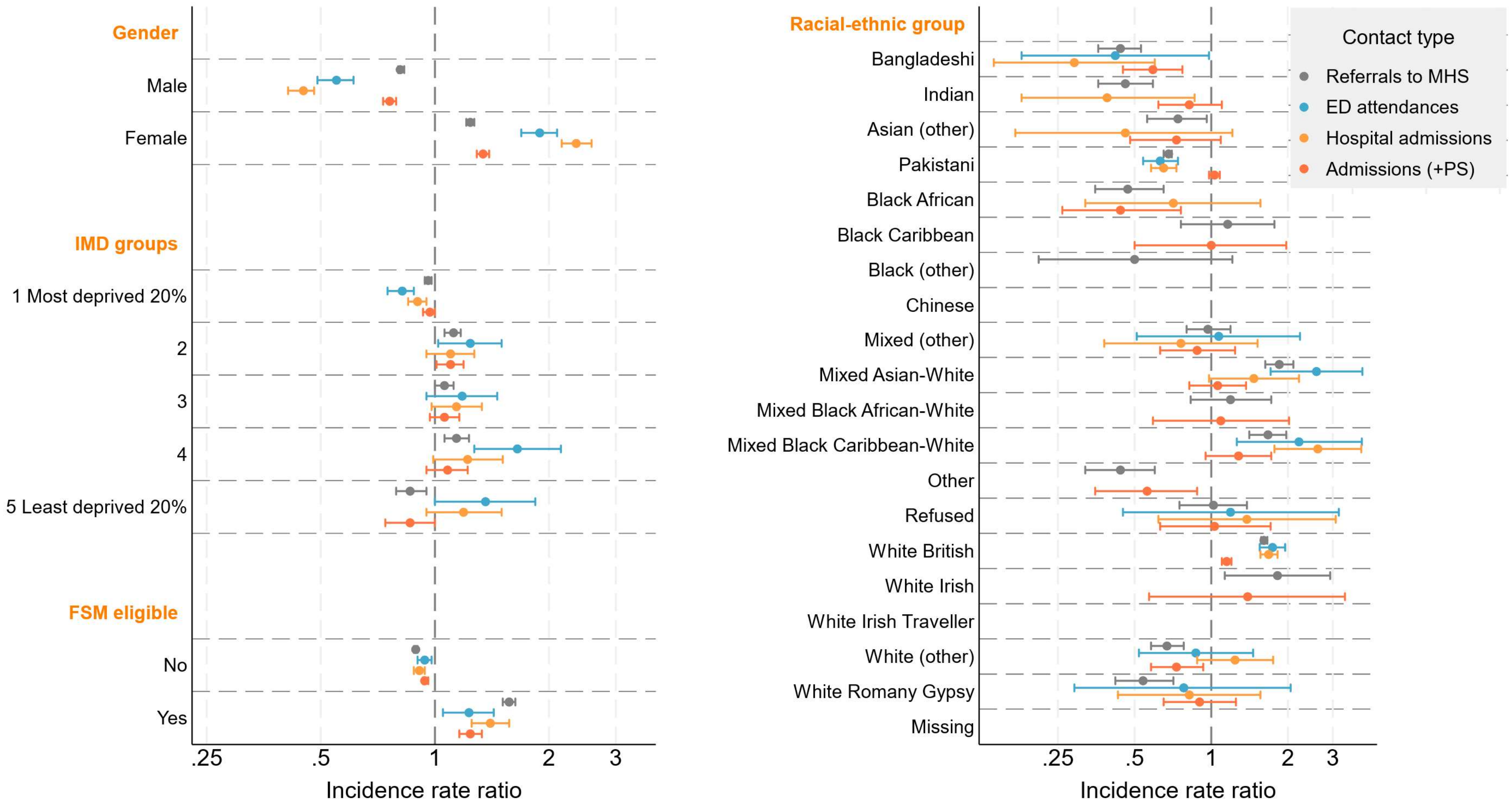

Gender
Male
Female
IMD groups
1 Most deprived 20%
2
3
4
5 Least deprived 20%
FSM eligible
No
Yes
.25
.5
1
2
3
Incidence rate ratio
Racial-ethnic group
Bangladeshi
Indian
Asian (other)
Pakistani
Black African
Black Caribbean
Black (other)
Chinese
Mixed (other)
Mixed Asian-White
Mixed Black African-White
Mixed Black Caribbean-White
Other
Refused
White British
White Irish
White Irish Traveller
White (other)
White Romany Gypsy
Missing
.25
.5
1
2
3
Incidence rate ratio
Contact type
Referrals to MHS
ED attendances
Hospital admissions
Admissions (+PS)

**Figure S17.** Year-adjusted incidence rate ratios (compared to the social strata- and service-specific mean rate) of first referral to mental health services, mental health-related emergency department attendances and hospital admissions (per 1000 person-years) with 95% confidence intervals among pupils residing in Bradford: derived from Poisson regression models with separate models for each social strata and service (11-<16 years old group only). IRRs based on estimated rates with confidence intervals less than 0 are not shown. ED = emergency department; FSM = free school meals; IMD = index of multiple deprivation; MHS = mental health services; PS = potentially psychosomatic symptoms
